# Genetic effects on the timing of parturition and links to fetal birth weight

**DOI:** 10.1101/2022.05.04.22274624

**Authors:** Pol Sole-Navais, Christopher Flatley, Valgerdur Steinthorsdottir, Marc Vaudel, Julius Juodakis, Jing Chen, Triin Laisk, Abigail L. LaBella, David Westergaard, Jonas Bacelis, Ben Brumpton, Line Skotte, Maria C. Borges, Øyvind Helgeland, Anubha Mahajan, Matthias Wielscher, Frederick Lin, Catherine Briggs, Carol A. Wang, Gunn-Helen Moen, Robin N. Beaumont, Jonathan P. Bradfield, Abin Abraham, Gudmar Thorleifsson, Maiken E. Gabrielsen, Sisse R. Ostrowski, Dominika Modzelewska, Ellen A. Nohr, Elina Hypponen, Amit Srivastava, Octavious Talbot, Catherine Allard, Scott M. Williams, Ramkumar Menon, Beverley M. Shields, Gardar Sveinbjornsson, Huan Xu, Mads Melbye, Lowe Jr William, Luigi Bouchard, Emily Oken, Ole B. Pedersen, Daniel F. Gudbjartsson, Christian Erikstrup, Erik Sørensen, Early Growth Genetics Consortium, Estonian Biobank Research Team, Danish Blood Donor Study Genomic Consortium, Rolv T. Lie, Kari Teramo, Mikko Hallman, Thorhildur Juliusdottir, Hakon Hakonarson, Henrik Ullum, Andrew T. Hattersley, Line Sletner, Mario Merialdi, Sheryl Rifas-Shiman, Thora Steingrimsdottir, Denise Scholtens, Christine Power, Jane West, Mette Nyegaard, John A. Capra, Anne H. Skogholt, Per Magnus, Ole A. Andreassen, Unnur Thorsteinsdottir, Struan F.A. Grant, Elisabeth Qvigstad, Craig E. Pennell, Marie-France Hivert, Geoffrey M. Hayes, Marjo-Riitta Jarvelin, Mark I. McCarthy, Deborah A. Lawlor, Henriette S. Nielsen, Reedik Mägi, Antonis Rokas, Kristian Hveem, Kari Stefansson, Bjarke Feenstra, Pål Njolstad, Louis J. Muglia, Rachel M. Freathy, Stefan Johanson, Ge Zhang, Bo Jacobsson

## Abstract

The timing of parturition is crucial for neonatal survival and infant health. Yet, its genetic basis remains largely unresolved. We present a maternal genome-wide meta-analysis of gestational duration (n = 195,555), identifying 22 associated loci (24 independent variants) and an enrichment in genes differentially expressed during labor. A meta-analysis of preterm delivery (cases = 18,797, controls = 260,246) revealed 6 associated loci, and large genetic similarities with gestational duration. Analysis of the parental transmitted and non-transmitted alleles (n = 136,833) shows that 15 of the gestational duration genetic variants act through the maternal genome, while seven act both through the maternal and fetal, and two act only via the fetal genome. Finally, the maternal effects on gestational duration show signs of antagonistic pleiotropy with the fetal effects on birth weight: maternal alleles that increase gestational duration have negative fetal effects on birth weight.

---

In humans, similar to mammals broadly, the timing of delivery is crucial for neonatal survival and health. Preterm delivery is the world-leading direct cause of death in neonates and children under five years of age^1^. While the rate of neonatal mortality has substantially decreased in recent years, the reduction attributable to preterm delivery is one of the lowest among the major causes of mortality ^2^. This partly reflects the relatively poor knowledge of the processes governing the timing of delivery in humans. Parturition may be initiated by a diversity of biological and mechanical pathways. Some of these are part of the physiological timing process, while others may override pregnancy maintenance with fail-safe mechanisms (e.g., in the case of uterine infection) ^3^. The diversity of the mechanisms has led to the conceptualization of preterm delivery as a syndrome ^4^, with various pathophysiological processes contributing to its etiology. Both maternal and fetal genomes are involved in these mechanisms. Yet, genetic studies have identified only a handful of loci associated with the timing of parturition ^5,6^.

Gestational duration is the major determinant of birth weight (i.e., the longer the gestation, the heavier the newborn). At the same time, uterine load is one of the known triggers of parturition ^7^, evidenced by half of twin pregnancies delivering preterm ^8^. Both the maternal and fetal genomes contribute to birth weight as well, as revealed in recent genome-wide association studies (GWAS) ^9,10^, and over evolutionary time may have even conflicted on gestational duration and birth weight, as proposed in the hypothesis of the genetic conflicts of pregnancy ^11^. This hypothesis suggests that the maternal genome favors slightly shorter gestations and lower birth weight while the fetal genome favors the opposite. Co-adaptation theory, instead, suggests that maternal and fetal genomes may invest resources to achieve an optimal gestational duration or birth weight that increases fitness ^12^. These known contributions, potential conflicts, and coadaptation of gestational duration and birth weight may ultimately create a complex relationship between the two.

What and how distinct are the maternal genetic effects on gestational duration and preterm delivery? What is the relationship between fetal growth and gestational duration? Is there evidence suggesting maternal-fetal co-adaptation on these traits? To address these questions, we conducted a GWAS meta-analysis of gestational duration and preterm and post-term delivery in >190,000 maternal samples with spontaneous onset of delivery. We further analyzed these results using the parental transmitted and non-transmitted alleles in >135,000 parent-offsprings.

## Results

### Genome-wide association analyses

We conducted a GWAS meta-analysis of gestational duration in 195,555 women of recent European ancestry (Supplementary Table 1), a four-fold increase in sample size compared to the largest published maternal GWAS of gestational duration ^5^. After quality control, genetic variants at 22 loci were associated with gestational duration at genome-wide significance (Fig. 1, Supplementary Table 2, and Supplementary Fig. 1). Approximate Conditional and Joint (COJO) analysis revealed two conditionally independent signals at *EBF1* and *KCNAB1* gene regions. Sixteen of the loci did not overlap with any previously reported gestational duration-associated locus ^5^. Effect sizes were relatively small, ranging from 7 (*HIVEP3/ EDN2*) to 27 (*MRPS22*) hours of gestation per allele (average duration of gestation = 282 days, 40.3 weeks). Heterogeneity in the effect estimates was limited to loci previously identified (*EBF1, WNT4, ADCY5, EEFSEC* and *AGTR2*), likely due to winner’s curse ^13^ (Supplementary Table 2 and Supplementary Fig. 2). Out-of-sample re-analysis of previously reported gestational duration-associated lead SNPs (n = 6) show that all four that were available after QC replicate at nominal significance (Supplementary Table 3). In addition, all 6 loci (± 250 kb from lead SNP) replicated at suggestive evidence.

**Fig. 1.**
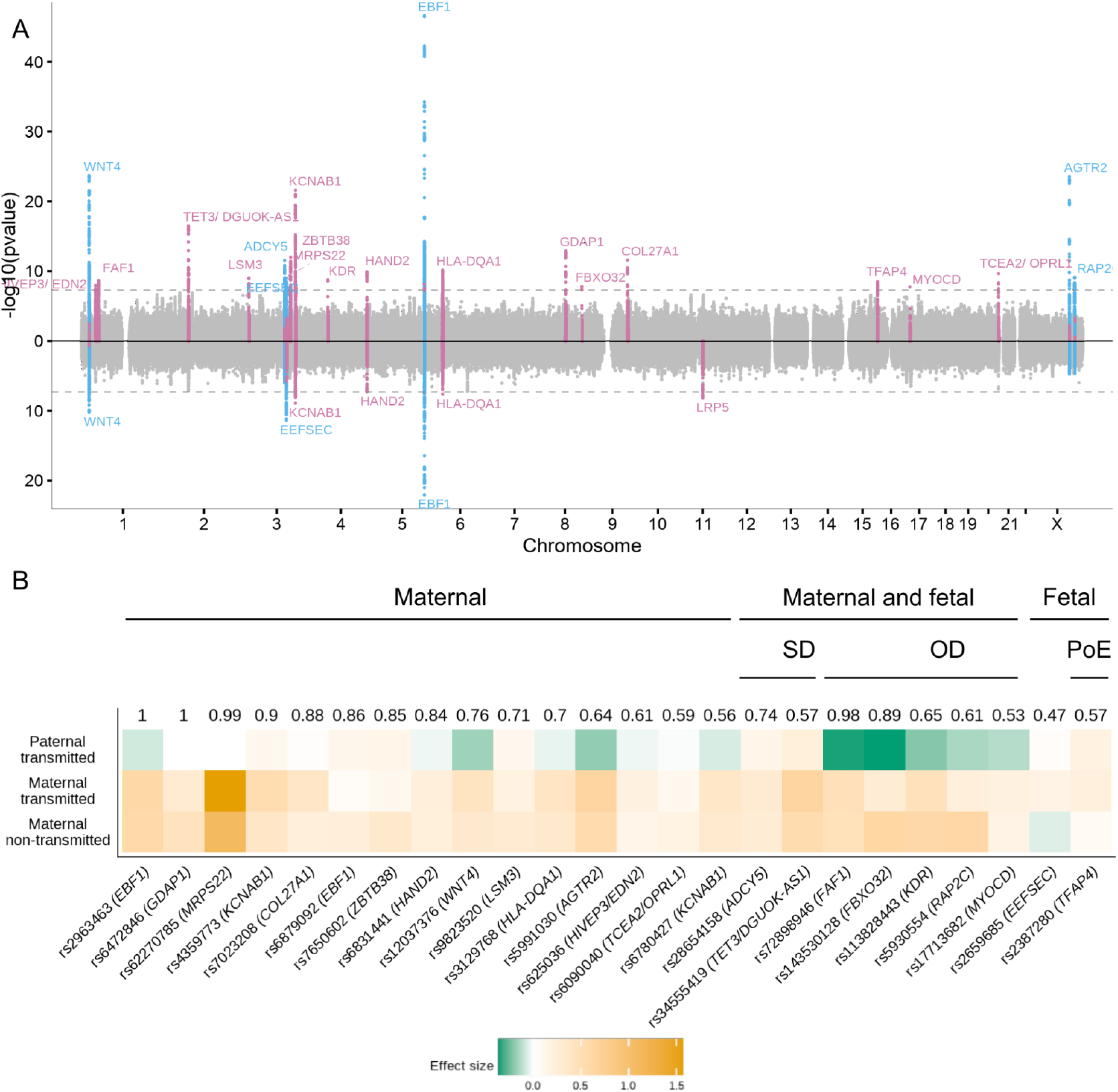
GWAS of the timing of parturition and dissection of maternal-fetal effects. (A) Miami plot illustrating the GWAS for gestational duration (top) and preterm delivery (bottom). The x-axis shows the chromosome position and the y-axis the two-sided p-value of the inverse-variance weighted meta-analysis. The dashed line represents the genome-wide significance threshold (p-value = 5×10^-8^). Each genome-wide significant locus is labeled by their nearest protein-coding gene. Blue, previously identified locus; pink, newly identified locus. (B) Clustering of the effect origin for the index SNPs for gestational duration using transmitted and non-transmitted parental alleles (n= 136,833). Numbers depicted above the heatmap are the highest probability observed for that SNP and group names define the cluster to which the highest probability refers to. The probabilities were estimated using model based clustering. Heatmap represents effect size and effect direction for the parental transmitted and non-transmitted alleles. For comparison purposes, the maternal alleles with positive effects were chosen as reference alleles. Three major groups were identified according to the highest probability: maternal only effect, fetal only effect and maternal and fetal effect. Within variants with both maternal and fetal effects two clusters were observed: same (“SD”) or opposite (“OD”) effect direction from maternal and fetal genomes. One of the fetal effects was further clustered as having a parent-of-origin effect (“PoE”), specifically, an effect from the maternal transmitted allele.

To prioritize candidate genes, we performed colocalization analysis ^14^ with cis-expression quantitative trait loci (eQTL) in induced pluripotent stem cells ^15^, endometrium ^16^, uterus, vagina and ovary ^17^ (Supplementary Table 4). eQTLs for seven protein coding (*OPRL1, ZBTB38, RGS19, TET3, COL27A1, CRISPLD1* and *ADCY5*) and four non-coding genes colocalized with gestational duration. Furthermore, colocalization analysis with blood protein QTLs ^18^ showed several trans associations: *ZBTB38* with three proteins, and *TCEA2/ OPRL1* and *WNT4* with one each. Particularly interesting are the associations with OPRL1 and POMC, which play a role in modulating nociception and pain perception; *in vitro* studies in tissues from pregnant rats and humans suggests that the administration of nociceptin inhibits uterine contractions, mediated by the *OPRL1* receptors ^19,20^.

RNA tissue-specific enrichment of top genes highlighted the endometrium and other female reproductive and smooth muscle tissues (Supplementary Fig. 3), results further supported at the genome-wide scale using stratified LD-score regression (Supplementary Fig. 4). Previous genetic studies have suggested a critical role of the decidua (endometrium) in the timing of parturition, indicating an effect early in pregnancy ^21^. Using stratified LD-score regression, we show that the heritability of gestational duration is enriched in regions harboring genes differentially expressed during labor (enrichment = 1.7, p-value = 7.1×10^-7^, Extended Data Fig. 1) ^22^, suggesting the SNPs associated with gestational duration may as well act during labor.

Stratified LD-score regression (Supplementary Fig. 5) revealed an enrichment in background selection, super enhancers, CpG content, H3K23ac and DNA methylation. Using the mosaic pipeline ^23^, we confirm that gestational duration loci have diverse evolutionary histories, including evolutionary conservation, excess population differentiation and negative selection (Supplementary Fig. 6).

We also performed a GWAS meta-analyses of preterm delivery (controls, delivery between 39 and 42 gestational weeks, n = 260,246; cases, delivery < 37 completed weeks, n = 18,797) and post-term delivery (controls, delivery between 39 and 42 gestational weeks = 115,307, cases > 42 completed weeks, n = 15,972) (Fig. 1A, Supplementary Table 2 and Supplementary Fig. 7-8). We observed a lower number of associated loci: 6 and 1 for preterm and post-term delivery, respectively. COJO analysis identified a secondary conditionally independent SNP associated with preterm delivery at the *EBF1* gene region. We identified only one locus associated with preterm delivery (rs312777, p-value = 6.6×10^-9^) that showed weak evidence of association with gestational duration (p-value = 3.9×10^-3^).

We observed a modest genetic correlation (r_g_ = -0.62; 95% CI = -0.72, -0.51) between gestational duration and preterm delivery, suggesting similarities between the two phenotypes (Supplementary Fig. 9 and Supplementary Fig. 10). Post-term delivery, instead, showed a perfect genetic correlation with gestational duration (r_g_ = 1.17; 95% CI = 0.93, 1.41), suggesting no differences in the maternal genetic effects on such traits.

### Resolving maternal-fetal effect origin

The genetic effects on pregnancy traits may be driven by two correlated genomes: the maternal and the fetal. To investigate whether the gestational duration signals originate in either or both genomes, we used phased genotype data to estimate the effects of the parental transmitted and non-transmitted alleles from 136,833 parent-offspring trios or mother-child duos (Fig. 1B, Supplementary Table 5 and Extended Data Fig. 2; the maternal samples of these duos/ trios were part of the GWAS meta-analysis). Based on pattern similarity using Gaussian mixture model–based clustering ^10^, SNPs were assigned to three large groups. Of the 24 index variants, 15 had the highest probability of a maternal effect, seven of both maternal and fetal effects (five, with opposite effect directions, and the remaining two, with the same direction). Finally, two variants were grouped as having a fetal only effect; the first, independent of the parent of origin (*TFAP4*, probability= 0.57), the second limited to the maternal transmitted allele (*EEFSEC)*. Caution should be taken when interpreting the latter considering the low probability (0.47).

The index SNP at *ADCY5* locus (rs28654158) had both maternal and fetal effects on gestational duration with the same effect direction. Interestingly, a SNP also located in the first intron of *ADCY5* harbors maternal and fetal effects on birth weight, but in opposite directions, attributed to the fetal insulin hypothesis ^9,10^. The two index SNPs for gestational duration (rs28654158) and birth weight (rs11708067) are located 50 kb apart from each other and are in low LD (R^2^ < 0.2). The birth weight SNP, also implicated in diabetes, likely acts through *ADCY5* ^24^, but it is unknown whether the gestational duration variant also acts through the same gene, although it colocalizes with *ADCY5* gene expression in the uterus (Supplementary Table 4). Despite being physically close to each other, differences between the two loci are evident in the traits they colocalize with. The gestational duration locus also affects fat-mass-related traits, while the birth weight locus affects glucose-related ones (Extended Data Fig. 3).

The only fetal index SNP identified to date in a GWAS (rs7594852; MAF = 0.49; beta = 0.37 days; 95% CI = 0.22, 0.51) ^6^ clustered as having a fetal only effect (Supplementary Table 5, probability = 1), independent of the parent of origin (beta paternal transmitted allele = -0.42, p-value = 2.7×10^-6^).

### Polygenic score of gestational duration and preterm delivery have similar prediction accuracy on preterm delivery

We built polygenic scores for gestational duration and preterm delivery using the corresponding GWAS results in the MoBa cohort (including the X chromosome) using LDpred2^25^ and estimated its effect on both traits. The polygenic score for gestational duration explains 2.2% of its variance (beta = 0.22 days per z-score; 95% CI = 0.02, 0.03; n = 3,943). The lowest decile had a mean gestational duration of 278 days (95% CI = 278, 279) while the highest decile had a mean of 283 days (95% CI = 282, 284) (Fig. 2). The polygenic score was also significantly associated with preterm delivery (Supplementary Table 6 and Supplementary Fig. 11, odds ratio = 0.994; 95% CI = 0.990, 0.997) with an area under the curve of 0.61 (95% CI = 0.55, 0.67). For comparison, a polygenic score for preterm delivery was built using the same samples as above. This polygenic score was also significantly associated with preterm delivery (Supplementary Table 6 and Supplementary Fig. 11, odds ratio = 1.005, 95% CI = 1.001, 1.009), with effect estimate similar to that obtained for the gestational duration polygenic score (after matching the direction). This reflects the genetic similarity between gestational duration and preterm delivery.

**Fig. 2.**
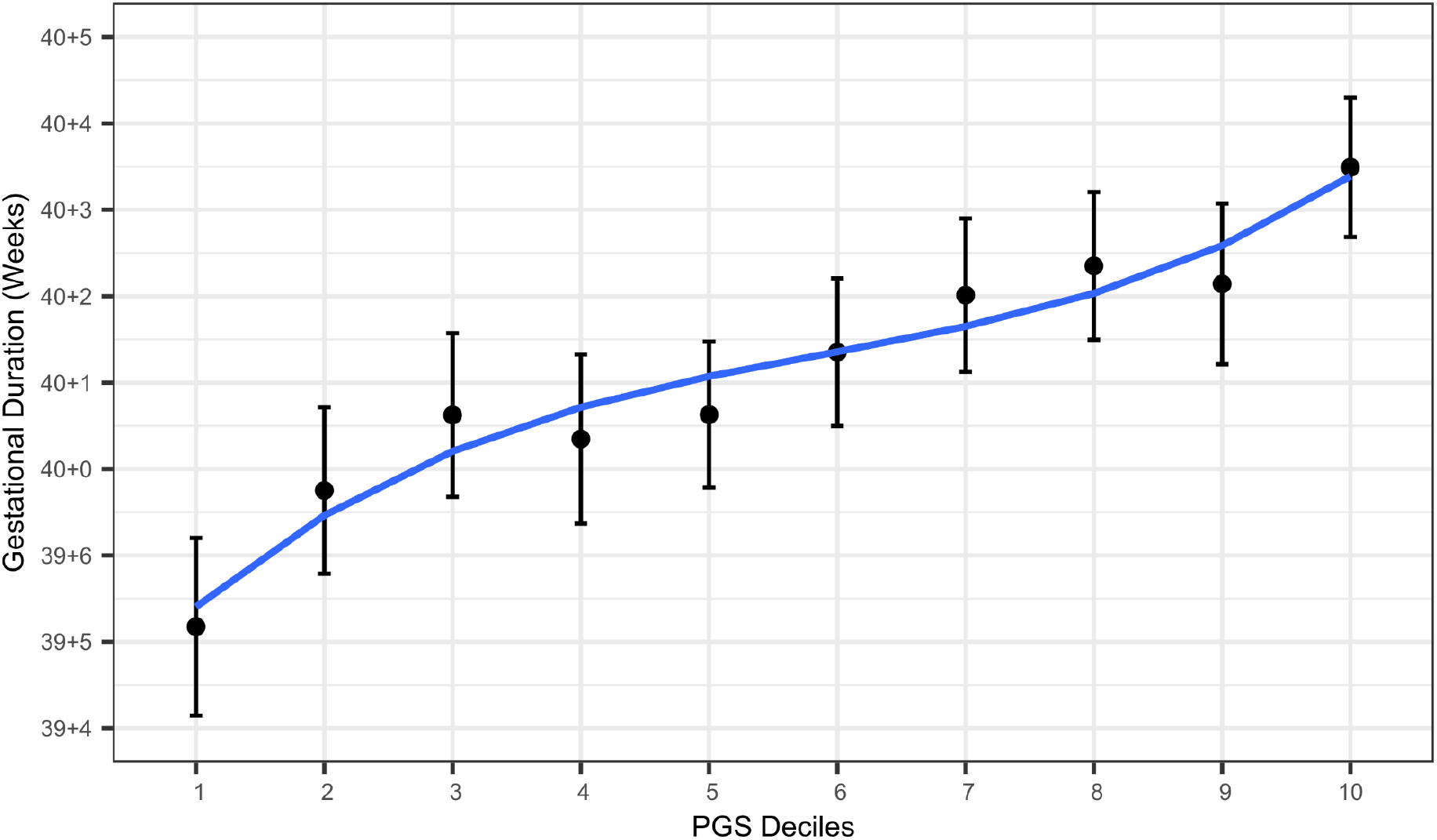
Polygenic prediction of gestational duration. Mean (95% CI) gestational duration for each decile of the gestational duration polygenic score (n = 3,943). Only spontaneous deliveries were considered.

### Strong pleiotropic effects between sex-hormones and the timing of parturition

To examine the potential shared genetic basis between the timing of parturition and other traits, we estimated the genetic correlations between 14 female reproductive traits and the maternal effects on gestational duration and preterm delivery (Fig. 3). These estimates were generally comparable, with the latter being consistently higher. Calculated bioavailable testosterone (CBAT, r_g_ = 0.40; 95% CI = 0.26, 0.54), testosterone (r_g_ = 0.35; 95% CI = 0.19, 0.51) and sex hormone binding globulin (SHBG, r_g_ = -0.16; 95% CI = -0.27, -0.06) in women were modestly genetically correlated with preterm delivery, whereas there was little genetic correlation with levels of the same hormones in men (Supplementary Table 7). We observed a positive genetic correlation between preterm delivery and the number of live births and, while this may be counter-intuitive, it is in line with a positive genetic correlation reported between miscarriage and the number of live births ^26^.

**Fig. 3.**
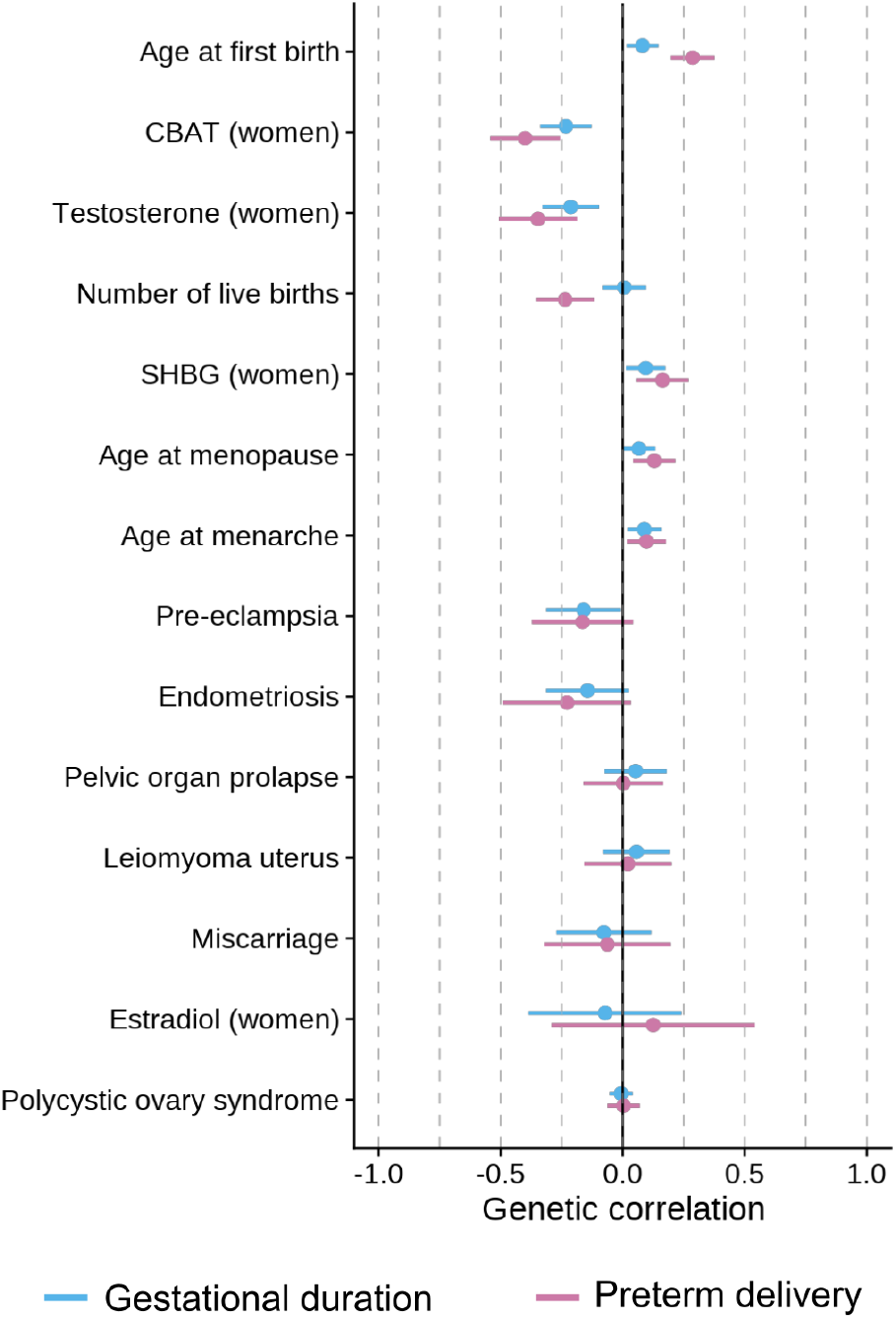
Genetic correlations between gestational duration and preterm delivery and other female reproductive traits. (A) Genetic correlations using LD-score regression. Dots are the genetic correlation estimate and error bars, the 95% CI. The direction of the genetic correlations with preterm delivery was flipped so that term deliveries were considered as cases and preterm deliveries as controls. Hence, the direction of the genetic correlations of preterm delivery matches that of gestational duration, providing a clear comparison of the 95% CI. Pink, preterm delivery; blue, gestational duration.

The genetic correlation between preterm delivery and the number of live births was twice as high in cohorts where the women’s whole reproductive history was available (r_g_ = 0.27; 95% CI = 0.11, 0.43) compared to cohorts based on a random pregnancy (r_g_ = 0.13; 95% CI = 0.00, 0.26), indicating an increased probability of preterm delivery with an increasing number of live births. We also detected a negative genetic correlation with age at first birth and age at menopause.

Genetic correlations can arise due to pleiotropy or due to a trait being causally upstream of the other. To distinguish between these situations, we used a latent causal variable (LCV) ^27^ model between sex-hormones, and preterm delivery and gestational duration (Supplementary Table 8). We observed evidence for full or nearly full genetic causality of CBAT, testosterone, and SHBG on preterm delivery (0.7 < GCP ≤ 0.8), but not on gestational duration (0.4 ≤ GCP < 0.5).

In a two-sample Mendelian randomization analysis, the concentrations of these sex hormones (Supplementary Table 9-10), including a set of variants that have consistent effects on testosterone, but no aggregate effects on SHBG ^28^, were associated with gestational duration and preterm delivery. While the MR-Egger intercept was not significantly different from 0 (Supplementary Table 10 and Extended Data Fig. 4), colocalization analyses across the genome confirmed that distinct variants underlie the associations for sex-hormones and the timing of parturition (Supplementary Fig. 12).

Using the parental transmitted and non-transmitted alleles in individual level parent-offspring data from Iceland and Norway (deCODE, MoBa and HUNT; n = 46,105 parent-offsprings, Supplementary Table 11), we observed a nominally significant association between the maternal non-transmitted alleles polygenic scores for CBAT and testosterone and gestational duration.

Testosterone and SHBG levels have a complex genetic link with the timing of parturition, likely explained by partial causality, as pointed out by the LCV analysis on gestational duration.

### Maternal, but not fetal effects on birth weight are partially mediated by gestational duration

We sought to understand the genetic relationship between gestational duration and birth weight and how the interplay between the maternal and fetal genomes affect this relationship. We used published summary statistics of birth weight (<15% of samples adjusted for gestational duration) derived from two different models ^9^: maternal only effect (adjusted by fetal effects) and fetal only effect (adjusted by maternal effects). These models were obtained using weighted linear modeling, and provide unbiased estimates for the maternal and fetal effects, respectively. The fetal effects on gestational duration were obtained from a previously published GWAS ^6^. The more recent GWAS meta-analysis of fetal growth ^10^ had >40% of samples adjusted for gestational duration, the reason why we did not use it in this section.

The maternal effects on gestational duration are strongly correlated with those on birth weight (Supplementary Fig. 13, r_g_ = 0.65; 95% CI = 0.54, 0.75). Conversely, neither the maternal (r_g_ = -0.05; -0.15, 0.04) nor the fetal (r_g_ = -0.02; 95% CI = -0.15, 0.11) effects on gestational duration were genetically correlated with the fetal only effects on birth weight. We suggest the maternal effects on birth weight are, at least partially, mediated by gestational duration, while the effects of the fetus on birth weight are not.

We then tested the extent of this mediation. Using multi-trait COJO analysis ^29^, we conditioned the genetic effects on birth weight on the maternal effects on gestational duration. After conditioning, the maternal effects on birth weight changed substantially: the SNP-heritability was reduced by 53% (p-value = 9.4×10^-7^, Supplementary Table 12), and the effect size of 87 suggestive SNPs dropped (Fig. 4A, median relative difference = -11%, Wilcoxon rank-sum test p-value = 1.3×10^-8^). Applying the same method on genome-wide significant variants classified with a “Maternal Only” effect on birth weight ^9^ provided very similar results (Supplementary Table 13 and Supplementary Fig. 14). This was further replicated using individual level data by directly adjusting for gestational duration in the linear model on birth weight (using genotypes in Icelandic data and the maternal non-transmitted alleles in MoBa, Norway, Supplementary Table 13 and Supplementary Fig. 14). In contrast, for fetal effects on birth weight, conditioning on gestational duration did not change the effect estimates or the heritability (Fig. 4A and Supplementary Table 12 for results with 108 suggestive SNPs and Supplementary Table 13 and Supplementary Fig. 14 with genome-wide significant variants classified as having a “Fetal Only” ^9^).

**Fig. 4.**
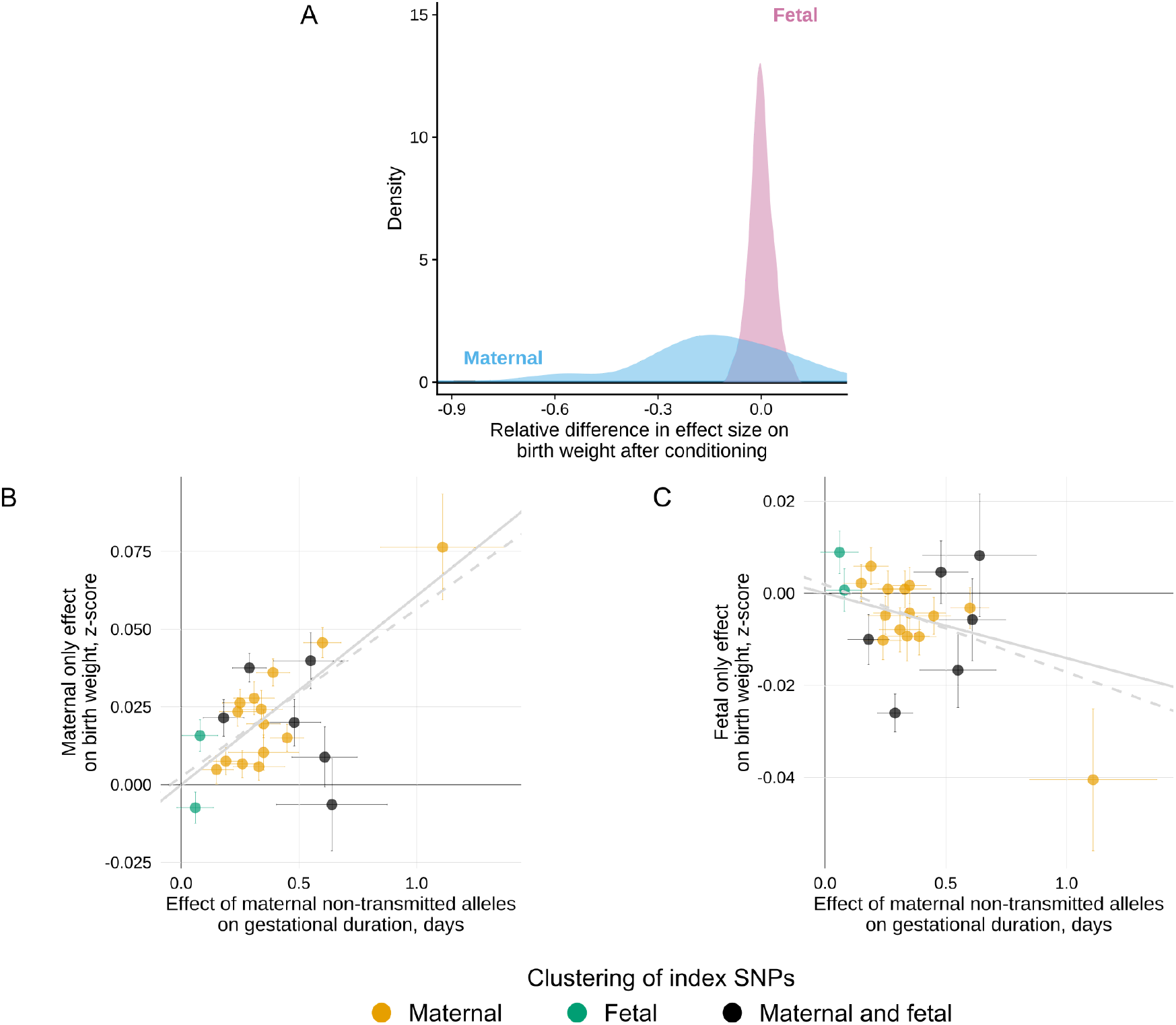
Genetic relationship between gestational duration and birth weight. (A) Distribution of the relative difference in effect size before and after conditioning the effect on birth weight by the maternal effect on gestational duration using approximate multi-trait conditional and joint analysis. In blue, relative difference in effect sizes for the maternal only effects on birth weight before and after conditioning; in pink, relative difference in effect sizes for the fetal only effects on birth weight after conditioning. After conditioning, we split the genome into approximately LD-independent regions and selected the SNPs with the lowest p-value on birth weight (p-value < 5×10^-6^) from each region (n SNPs maternal effect = 87; n SNPs fetal effect = 108). (B, C) Scatterplot for two-sample Mendelian randomization analysis for the maternal effect of gestational duration on birth weight (B, maternal or C, fetal effects). Each dot represents one of the gestational duration index SNPs. Effect sizes and standard errors from the index SNPs for gestational duration derived from the maternal non-transmitted alleles were obtained from the meta-analysis of parent-offspring data (n= 136,833). The maternal only and the fetal only effects on birth weight were extracted from a previous GWAS meta-analysis (n= 210,248 and 297,356, respectively). The x-axis shows the SNP effect of the maternal non-transmitted alleles on gestational duration, and the y-axis the effect on birth weight. Horizontal and vertical error bars represent the standard error. The gray line depicts the inverse-variance weighted method estimate, and the gray-dashed line the MR-Egger estimate. Colors represent the clustering of the SNP effects on gestational duration, performed using model based clustering.

In summary, while the maternal effects on birth weight are partially driven by gestational duration, we found no evidence for this for the fetal effects on birth weight.

### The maternal genome drives the phenotypic association between gestational duration and birth weight

It is widely accepted that longer gestations lead to heavier newborns. Here, we sought to obtain causal estimates of the effect of gestational duration on birth weight.

We used the index SNPs from our discovery GWAS and the effect estimates from the maternal non-transmitted alleles as genetic instruments in a two-sample Mendelian randomization analysis (Fig. 4B and Supplementary Fig. 15) on the maternal only effects on birth weight (derived using a weighted linear model ^9^). The maternal non-transmitted gestational duration-increasing alleles were associated with higher birth weight (beta = 0.06 z-scores per day; 95% CI = 0.05, 0.08; p-value = 1.7×10^-16^). The estimated effect (approximately 23 g per day) is concordant with the phenotypic association between gestational duration and birth weight (25 g per day in 18,452 samples from the MoBa cohort). We observed no effect from the paternal transmitted gestational duration-increasing alleles on birth weight. The LCV model confirmed a full or nearly full causal (GCP = 0.6, p-value = 0.002, Supplementary Table 8) effect of gestational duration on birth weight.

### Maternal effects on gestational duration and fetal effects on birth weight exhibit signs of antagonistic pleiotropy

First, we evaluated the impact of fetal growth on gestational duration by instrumenting fetal growth using 68 SNPs with fetal only effect on birth weight (n= 35,280 and 48,741 parent-offsprings, Supplementary Table 14) ^9^. Higher paternally transmitted birth weight score was associated with shorter duration of gestation and the estimated effect was larger when estimated using the last menstrual period (beta = -1.9 days per z-score, p-value = 4.0×10^-4^) than ultrasound. This result supports previous evidence showing faster fetal growth is associated with shorter duration of gestation ^30^. To investigate whether this was due to antagonistic pleiotropy between the fetal effects on birth weight and the maternal effects on gestational duration, we assessed the relation between birth weight-increasing alleles and maternal effects on gestational duration. The fetal birth weight-increasing alleles were not associated with maternal effects on gestational duration (Supplementary Table 15), suggesting that the results presented above are likely not due to antagonistic pleiotropic effects.

Next, we used summary statistics to investigate potential pleiotropy between the genetic effects on gestational duration and fetal birth weight. Using methods borrowed from Mendelian randomization analysis, we evaluated the association between the maternal gestational duration-increasing alleles and the fetal effects on birth weight. We observe that the alleles that increase gestational duration through a maternal effect tend to reduce birth weight through a fetal effect (Fig. 4C and Supplementary Table 15). Interestingly, this effect was not limited to the maternal transmitted alleles (beta = -0.02 z-scores per day; 95% CI = -0.03, -0.01; p-value = 3.4×10^-4^), but was also observed for the maternal non-transmitted gestational duration increasing alleles (beta = -0.01 z-scores per day; 95% CI = -0.02, -0.01; p-value = 6.2×10^-3^). The paternal transmitted gestational duration increasing alleles were not associated with fetal only effects on birth weight (Supplementary Table 15).

## Discussion

The timing of parturition is crucial for neonatal survival and health. Yet, discovery of maternal and fetal genetic effects lags behind that of other pregnancy traits such as birth weight ^9^ and fetal growth ^10^. In this GWAS meta-analysis of parturition timing, we identified 17 loci not previously reported, one of which was more strongly linked to preterm delivery than to gestational duration. The results support large similarities in the maternal genetic effects on gestational duration and preterm delivery. By including parent-offspring data with a similar sample size to that of the discovery GWAS, we were able to discern maternal from fetal effects with high certainty in most index SNPs. Finally, the results show a complex genetic relationship between the maternal and fetal genomes on gestational duration and birth weight.

Our understanding of the molecular signals governing the timing of parturition in humans has not advanced significantly. Previous genomic evidence suggests a critical role of the decidua ^21^, denoting an effect on the timing of parturition as early as implantation. We report that the SNP-heritability of gestational duration is enriched in genes differentially expressed during labor in the myometrium. We suggest the maternal effects on the duration of gestation may as well act during labor, for instance, by inhibiting uterine contractions. Genetic studies of gestational duration may prove useful in the discovery of drug targets as tocolytic agents or for labor induction. At the same time, the genetic effects on gestational duration and preterm delivery are largely similar; this is opposed to the heterogeneity observed at the phenotypic and transcriptomic levels ^31,32^. As an example, while the polygenic score of gestational duration is still inadequate for clinical use, it had a similar effect on preterm delivery as a polygenic score of preterm delivery itself.

Gestational duration is the major determinant of birth weight. While the maternal genome affects offspring birth weight through many different causal pathways (e.g., maternal glucose levels ^9,10^), the effects are partly mediated by gestational duration. This has implications on the interpretation of GWAS of birth weight and downstream analyses, such as Mendelian randomization. In contrast with this, the fetal genetic effects on birth weight are not mediated by gestational duration, suggesting the fetal genome mainly acts on birth weight by modulating fetal growth. Interestingly, the maternal gestational duration increasing alleles have negative fetal effects on birth weight, likely reflecting antagonistic pleiotropy. The opposite was not true; fetal birth weight increasing alleles were not associated with maternal effects on gestational duration. We speculate that the fetal effects on birth weight have likely coadapted to increase the fitness of the fetus in pregnancies genetically predisposed to a shorter duration. It has been suggested that both gestational duration and birth weight are under balancing selection, with intermediate values of these traits having highest fitness ^3,33^. As exemplified here, this could lead to antagonistic pleiotropy favoring the coadaptation of maternal and fetal effects to attain optimal gestational duration and birth weight ^12^.

The presented results have several limitations. First, we analyzed data from participants of European ancestry. Over 70% of the samples were obtained from Nordic countries, with genotype data linked to the Medical Birth Registers; in these countries, the preterm delivery rate is one of the lowest in the world ^1^. Studying diverse ancestries would propel the identification of novel loci associated with gestational duration and aid in fine-mapping efforts, as has been previously shown for other traits ^34^. Second, to understand the relationship between gestational duration and birth weight, we used summary statistics from a previously published birth weight GWAS that was partially adjusted for gestational duration (<15% of samples) and excluded preterm deliveries. This is likely to affect our analyses by reducing their power. Third, we assumed a causal association between gestational duration and birth weight. While this is known to be true to some extent (i.e., longer gestations are linked to heavier newborns), pleiotropy between gestational duration and birth weight could be very well at play. And, fourth, phenotypic heterogeneity between cohorts (e.g., gestational duration estimation method) may have hindered the identification of additional signals.

In conclusion, the present results provide evidence of large genetic similarities between gestational duration and preterm delivery and further our understanding of the complex relationship between gestational duration and birth weight, likely shaped by strong evolutionary forces. Particularly, we showed that the maternal effects on birth weight are largely driven by gestational duration and that the maternal and fetal genomes have antagonistic pleiotropic effects on gestational duration and birth weight.

## Online Methods

### Phenotype definition

In this study we included pregnancies with a singleton live birth and a spontaneous onset of delivery: medically initiated deliveries (either by induction or planned cesarean section) were excluded or part of controls for preterm delivery. Gestational duration in days was estimated using either the last menstrual period date or ultrasound. We excluded pregnancies lasting <140 days (20 completed weeks) or >310 days (44 completed weeks), as well as women with health complications prior to or during pregnancy and congenital fetal malformations. Spontaneous preterm delivery was defined as a spontaneous delivery <259 days (37 completed gestational weeks) or by using the ICD-10 O60 code, and controls as a delivery occurring between 273 and 294 days (39 and 42 gestational weeks). Post-term delivery was defined as a delivery occurring >294 days (42 completed weeks) or ICD-10 O48 code, and controls as a spontaneous delivery between 273 and 294 days (39 and 42 gestational weeks). Given the perfect genetic correlation between gestational duration and post-term delivery GWAS, and the small power of the latter, all downstream analyses are focused on gestational duration and preterm delivery.

### Study cohorts and individual-level GWAS

This study consists of cohorts participating in the Early Growth Genetics (EGG) Consortium and the Norwegian Mother, Father and Child Cohort study (MoBa) ^35^, deCODE genetics ^10^, Trøndelag Health Study (HUNT) ^36^, Danish Blood Donor Study (DBDS) ^37^, the Estonian Genome Center of the University of Tartu (EGCUT) ^38^ and summary statistics from FinnGen ^39^ and a previous GWAS of gestational duration and preterm delivery performed using 23andMe, Inc, data ^5^. A total of 18 different cohorts (Supplementary Table 1) provided GWAS data under an additive model for meta-analysis for the maternal genome, resulting in 195,555 samples for gestational duration, 276,218 samples for preterm delivery (n cases= 18,797) and 115,307 samples for post-term delivery (n cases= 15,972) of recent European ancestries (indicated by principal component analysis). For binary outcomes (preterm and post-term deliveries), only cohorts with an effective sample size > 100 were included. Detailed description of the cohorts included can be found in the Supplementary text. All study participants provided a signed informed consent, and all research studies were approved by the relevant institutional ethics review boards (Supplementary Text).

Each individual cohort applied specific quality control procedures, data imputation and analysis independently following the consortium recommendations. Unless more stringent, samples were excluded if genotype call rate <95%, autosomal mean heterozygosity >3 standard deviations from the cohort mean, sex mismatch or major recent ancestry was other than European (HapMap central european). Genetic variants were excluded if genotype call rate <98%, Hardy-Weinberg equilibrium p-value <1×10^-6^ or minor allele frequency (MAF) <1%. Reference panels for imputation were either 1000 Genomes Project (1KG) ^40^, Haplotype Reference Consortium (HRC) ^41^, 10KUK, or a combination of one of the mentioned reference panels and own whole-genome sequencing data (deCODE, HUNT, DBDS, and FinnGen). Each individual cohort performed a GWAS using an additive linear regression model adjusted for, at least, genetic principal components or relationship matrix on autosomal chromosomes and chromosome X. Summary statistics for each individual cohort were stored centrally and underwent quality control procedures before meta-analysis. Genetic variant ids were converted to ‘CHR:POS:REF:EFF’ (positions were mapped to the Genome Reference Consortium Human Build 37, hg19), where EFF was the alphabetically higher allele - effect sizes were aligned accordingly. Alleles for insertion/deletions were coded as ‘I/D’, respectively. Only sequence variants from the Haplotype Reference Consortium panel or 1000 Genomes Project were included in the meta-analysis and genetic variants with a MAF > 0.05%, minor allele count > 6, an imputation INFO score > 0.4, MAF +-20% compared to HRC or 1KG, and a reported p-value with a less than 10% difference with a calculated p-value (from the z-score) in the -log_10_ scale were included.

### Meta-analysis of genome-wide association studies

After quality control, individual-cohort GWAS summary statistics were pooled using fixed effects inverse-variance weighted meta-analysis with METAL ^42^ without genomic control correction. We also performed an analysis of heterogeneity of effects (Supplementary Table 2, *I*^2^ statistic). After meta-analysis, we removed genetic variants reported in less than half the number of available samples for each phenotype, resulting in 9-10 million genetic variants. For example, the variant observed in the largest number of samples for gestational duration was available in 195,555 individuals, only variants reported in at least 97,778 were kept. Genomic inflation factors were low for all three phenotypes (Supplementary Table 16, gestational duration λ = 1.14, preterm delivery λ = 1.08 and post-term delivery λ = 1.05). LD-score regression intercepts were substantially lower than genomic inflation factors, suggesting that the inflation in test statistics was mostly due to polygenicity (Supplementary Table 16). Test statistics were not further adjusted for genomic control for any of the phenotypes. If not otherwise stated, all analyses presented in this manuscript are two-sided tests.

Initially, we naively defined independent loci based on physical distance, where SNPs within 250 kb from the index SNP were considered to be at the same locus. Novel loci were defined as loci not overlapping previously reported gestational duration loci in the largest GWAS performed to date ^5^. Finally, we used conditional analysis to resolve independent loci (see below).

### Conditional analysis

We looked for conditionally independent associations within each locus using approximate Conditional and Joint (COJO) analysis ^43^ implemented in Genome-wide Complex Trait Analysis (GCTA) software ^44^. We ran a stepwise model selection (-cojo-slct) to identify conditionally independent genetic variants at p-value< 5×10^-8^ for each of the genome-wide significant loci (using a radius of 1.5 Mb from the index SNP). Overlapping loci were merged into a single locus (only two loci overlapped, at 3q23). LD between genetic variants was estimated from 19,092 maternal samples from the Norwegian Mother, Father and Child Cohort, after excluding variants with imputation INFO score< 0.4. We converted the reference panel from BGEN files to hard-called PLINK binary format (.bed). As per default in COJO, genetic variants >10 Mb apart were assumed to be in complete linkage equilibrium.

### Gene prioritization

To prioritize genes at the gestational duration loci identified, we set the baseline as the nearest protein coding gene to the index SNP at each independent locus. While naive, this approach has been consistently shown to outperform other single metrics for locus-to-gene mapping ^45,46^. Next, we performed colocalization analysis for cis-eQTLs in 1,367 human iPSC lines from the i2QTL resource (± 250 kb from gene start and stop position) ^15^, endometrium (± 250 kb from gene start and stop position) ^16^ and uterus, vagina and ovary from GTEx (± 1 Mb around transcription start site) ^17^. Unfortunately, none of the variants we identified were in LD (R^2^ > 0.6) with missense variants. To complement the prioritization of genes, we queried each of the index SNPs for blood protein QTLs ^18^(both in cis and trans). For all index SNPs that were protein QTLs (p-value < 5×10^-6^), we performed colocalization analyses (± 1.5 Mb around the index SNP). We excluded the *HLA* region due to its large pleiotropic effects.

### Colocalization

We utilized genetic colocalization to identify pleiotropic effects between gestational duration and expression and protein quantitative trait locus (see Gene prioritization) and with other female and reproductive traits. To this end, we applied COLOC ^14^, which evaluates, in a Bayesian statistical framework, whether a single locus from two different phenotypes best fits a model where the associations are due to a single shared variant or distinct variants in close LD. For each tested locus, this information is summarized in the posterior probability of five hypotheses. Given phenotypes A and B, in a specific locus:

- No association for any of the two phenotypes
- Association with phenotype A but not with phenotype B
- Association with phenotype B but not with phenotype A
- Association with both phenotypes, association driven by SNPs in low LD
- Association with both phenotypes, association driven by SNPs in close LD

The last two hypotheses explicitly model an association with both phenotypes at the same locus, but in the former, the leading SNPs are not shared (i.e., lead variants for the two phenotypes are in low LD) and in the other the leading SNPs are shared (i.e., lead variants for the two phenotypes are in close LD), despite not knowing which one is the causal one.

Prior probabilities for each for the non-null hypotheses were set as suggested by Wallace (prior probabilities that a random SNP in the loci is associated with phenotype A, phenotype B, or both phenotypes, 1×10^-4^, 1×10^-4^, and 5×10^-6^, respectively), which are considered more conservative than the ones set by default ^47^.

Strong evidence of colocalization was defined as a posterior probability of colocalization >0.9.

### Enrichment analysis

We tested for enrichment based on top loci and genome-wide using partitioned LD-score regression. To test for over-representation in tissue-specific RNA expression (Human Protein Atlas, RNA consensus tissue gene data) ^48^ a Wilcoxon rank-sum test was performed on normalized RNA for genes within our set (above-mentioned) and all other genes. Significance for this test was set at Bonferroni correction for the number of tissues (p-value<0.05 / 61), and suggestive evidence at p-value< 0.1/ 61. At the genome-wide level, we performed partitioned heritability using LD-score regression to test for enrichment in 97 different annotations ^49,50^ and tissue-specific RNA expression using 205 different tissues/ cell types ^51^, using pre-computed partitioned LD-scores for subjects of recent European ancestry (baseline-LD model v2.2). We further explored the possibility that the genetic effects on gestational duration were acting late in pregnancy. To this end, we applied stratified LD-score regression to investigate whether SNP-heritability was enriched in regions harboring genes differentially expressed during labor in single-cells from myometrium^22^. We calculated LD scores (European individuals from phase 3 of the 1000 Genomes project) for sets of genes differentially expressed at labor (± 100 kb) for each cell type separately and for the overall set of genes differentially expressed in the myometrium. In the manuscript, we report exclusively the analysis performed on the overall set of genes, given that the -log10(p-value) for enrichment was a linear function of the number of differentially expressed genes, which ranged from 2 to >3000 (Supplementary Fig. 16). In this context, it is unwise to compare the enrichment in the different cell types. Stratified LD-score regression was run together with the baseline model mentioned above.

### Genetic correlations

We estimated genetic correlations by performing LD-score regression ^52^ locally using pre-computed LD-scores from 1000 Genomes Project samples of recent European ancestry. The MHC region (chr6:28477797-33448354) was removed prior to running LD-score regression.

### Resolving effect origin

To classify the identified index SNPs for gestational duration as having maternal, fetal or maternal and fetal origin, we performed an association analysis using the parental transmitted and non-transmitted alleles on gestational duration. We used phased genotype data (i.e. estimated haplotypes) in parent-offsprings or mother-child duos to infer the parent-of-origin of the genotyped/ imputed alleles as previously described ^30^. Once the transmitted allele was identified, the non-transmitted maternal allele was extracted. Briefly, parental origin of each allele was inferred using genotypes of relatives, reference cohort data, or distributions of genotypes within the cohort and LD measurements. Different methods were used for phasing in each of the cohorts providing data for this analysis (see Supplementary Table 17). In Icelandic data, parental alleles were inferred by combining long-range phasing, genealogy and a maximum likelihood estimation ^10,53,54^. All other cohorts were phased using SHAPEIT2 ^55^, except HUNT which was phased using Eagle v2.3 ^56^. The two methods use a hidden Markov model in combination with information from genetic relatives to refine phase calls and assign parents to haplotypes. Reliability of all these methods is considered to be high, particularly when large amounts of identical-by-descent segments are present (i.e., when parent-offspring data is available).

For each index SNP we fit the following linear regression model:

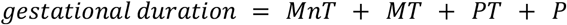

where *MnT* and *MT* refer to the maternal non-transmitted and transmitted alleles respectively, and PT refers to the paternal transmitted alleles. The latter is interpreted as a fetal only genetic effect, while the effect of the maternal non-transmitted allele is a maternal only genetic effect. We first estimated the effects of the index SNPs in each birth cohort separately; effect sizes were then combined through fixed-effect meta-analysis, totalling a sample size of 136,833 (104,962 parent-offspring trios from Iceland with at least one genotyped individual, 17,024 parent-offspring trios from the MoBa cohort, 5,122 parent-offspring trios from the HUNT cohort, and 9,725 mother-child duos from the Avon Longitudinal Study of Parents and Children (ALSPAC), Finnish birth data set (FIN), the Danish National Birth Cohort (DNBC), the Genomic and Proteomic Network for Preterm Birth Research (GPN) and the Hyperglycemia and Adverse Pregnancy Outcome (HAPO)). The analysis of the Icelandic data was done on 104,962 parent-offspring trios with at least one genotyped individual. This includes 18,165 fully genotyped trios, 5,208 with only child and mother and 1,875 with only child and father genotyped, 40,182 with both parents genotyped but not the child, and 1,627, 24,965 and 12,868 with only child, mother or father genotyped, respectively.

To classify the identified genetic variants into classes with similar patterns of effect we used model based clustering ^10^. Variants were clustered based on estimated effects of the transmitted and non-transmitted parental alleles into five clusters. Two clusters assume fetal effect only, one with effect independent of parent-of-origin and one where the effect is limited to the maternally transmitted allele; a cluster with maternal effect only; and two clusters with both maternal and fetal effects, either in opposite or same direction.

### Locus pleiotropy at 3q21

After identifying locus pleiotropy between the maternal effect on gestational duration and the fetal only effect on birth weight at the *ADCY5* gene region, we set out to investigate differences between the two top SNPs in their colocalization with other traits. Phenome-wide colocalization for the two regions (defined as 1.5Mb around the index SNP) was performed using summary statistics from FinnGen (data freeze 5) and Pan UK Biobank data ^57^ (in subjects of recent European ancestry). We included all phenotypes available from FinnGen (n traits= 2,803), while for Pan UK Biobank, we reduced it to summary statistics with an estimated heritability >0.01 and that were labeled as biomarkers, continuous trait or ICD-10 codes (n traits = 832). Given the exploratory nature of this analysis, despite using 3,635 phenotypes, we used a lenient posterior probability of colocalization (>= 0.75).

### Female reproductive traits

We obtained summary statistics for several female reproductive traits from different sources (minimum sample size 10,000). We included summary statistics from the following traits: miscarriage ^26^, gestational duration (fetal genome) ^6^, age at first birth, age at menarche ^58^, age at menopause ^59^, number of live births ^58^, testosterone ^60^, CBAT ^60^, SHBG ^60^, oestradiol (women)^58^, pelvic organ prolapse (FinnGen), polycystic ovary syndrome (^61^ and FinnGen), endometriosis ^58^, leiomyoma uterus (FinnGen) and pre-eclampsia ^62^. For polycystic ovary syndrome, we meta-analyzed summary statistics from the largest published GWAS ^61^ and FinnGen. We estimated genetic correlations between gestational duration and preterm delivery and these traits, and latent causal variable analysis between sex hormones (testosterone, CBAT and SHBG) and gestational duration and preterm delivery. We further explored causality using two-sample Mendelian randomization and inspected whether the effects originated in the maternal or the fetal genome (see below “Mendelian randomization”). Finally, when one trait is causally upstream of the other, it is expected that the two traits would share a causal variant at some of the trait-associated loci. To test for this at the genome-wide scale, we performed colocalization analysis between sex-hormones and gestational duration and preterm delivery using approximately LD independent regions^63^.

To obtain GWAS estimates for preterm delivery independent of the number of live births, we split the cohorts into two groups and then meta-analyzed per strata: on one side, cohorts based on a random pregnancy per mother (the probability of having at least one preterm delivery is not affected by the number of previous or subsequent deliveries) and cohorts with whole reproductive history of a woman (i.e., cohorts using life-time ICD codes or with data on > 1 pregnancy for the same mother).

### Latent causal variable analysis

We used latent causal variable analysis to distinguish (partial) causation from genetic correlation ^27^. For this, we used the traits that were genetically correlated with gestational duration or preterm delivery (birth weight or sex-hormones). In the LCV model, a latent variable mediates the genetic correlation between two phenotypes. The genetic causality proportion, which quantifies the proportion of the genetic correlation that is due to causality, is then estimated using mixed fourth moments. Whenever GCP p-values were significant, we defined GCP>= 0.6 between two traits as evidence of full or nearly full genetic causality, and GCP< 0.6 as evidence of limited partial causal implication.

### Gestational duration and preterm delivery polygenic score analysis

To obtain an independent sample for training and validation of a polygenic score, the meta-analyses for gestational duration were rerun, excluding the MoBa cohort. These new meta-analysis results were used as the base datasets to calculate the polygenic scores. After applying the same exclusion criteria to it as used for the study samples in the meta-analysis, and removing duplicated samples and those with a kinship of greater than 0.125, the MoBa cohort was randomly split, using 80% (n=15,768) as the training cohort and the remaining 20% (n=3,942) as the validation cohort.

### QC of Training Genotypes Dataset

From the training cohort, genotypes were excluded if they had a minor allele frequency of less than 0.01, info scores < 0.7, a Hardy–Weinberg equilibrium less than 1.0×10^-6^ and genotype call rates less than 0.01.

### Polygenic Score Calculation

LDpred2 was used for the calculation of the polygenic scores ^25^. As we wanted to include the X chromosome in the polygenic score, we mapped the genotypes to the genetic map taken from Bolt-LMM ^64^. Polygenic scores were calculated for a range of models using a grid of hyperparameter values; proportion of causal SNPs from 10^-5^ to 1 for 21 values, and proportions of heritability of 0.7, 1, and 1.4, calculated from a constrained LD-score regression. LDpred2 also uses a third hyperparameter that allows for sparse effect size estimates (i.e. some effects are exactly 0). This resulted in a total of 126 combinations of hyperparameter values for the range of grid models ^25^. The variance explained was used to decide which of the grid models was the most appropriate polygenic score.

We found the polygenic score (with ten principal components and adjusted for genotyped batch) that utilised the hyperparameters of proportion of causal SNPs of 0.0032, 0.7 of the heritability, and did not allow for sparse effect size estimates, was the most appropriate for the training cohort. This model accessed weighted betas from 1,123,366 variants to explain 2.3% of the variability in the testing sample. We then extracted the weighted betas for each variant from this model to be used in the polygenic score validation.

### Polygenic Score Validation

The remaining MoBa cohort was used for validation of the polygenic score. The SNPs included from the polygenic score model that explained the most variance in the training data were extracted from the genotyped data of the validation cohort. These SNPs were then used with the weighted betas from the training model to calculate a polygenic score for each individual in the validation cohort. These polygenic scores were converted to z-scores to enable comparison of polygenic scores between those derived from the gestational duration linear model and the preterm delivery logistic models.

To test the performance of the polygenic score, a linear regression was conducted for gestational duration by the polygenic score. A second model was used that adjusted for 5 principal components and genotyped batch. R^2^ was calculated for the models to quantify variance explained.

The utility of the polygenic score for the prediction of preterm delivery was also assessed. Gestational duration was dichotomized into preterm delivery (less than 37 weeks) or full term (greater than or equal to 39 weeks and less than 41 weeks). Two models were analyzed, one assessing just the polygenic score and a second adjusting for 5 principal components and genotype batch. Receiver operating characteristic, area under the curve were calculated for each model and used as assessment of diagnostic accuracy.

### Multi-trait conditional analysis

GCTA was used to perform bi-directional multi-trait COJO (mtCOJO) ^29^ analysis using summary statistics. The gestational duration GWAS was conditioned on the birth weight GWAS and vice-versa, using birth weight summary statistics from the largest GWAS meta-analysis of birth weight ^9^. We did not condition on the fetal effects on gestational duration due to a lack of power in the fetal GWAS ^6^. We obtained birth weight summary statistics from four different GWAS within the EGG Consortium: using the maternal genome - offspring birth weight, the fetal genome - own birth weight and using a weighted linear model to adjust the GWAS of offspring birth weight by the fetal genome, and the GWAS of own birth weight by the maternal genome.

To select variants with suggestive association with maternal or fetal effects on birth weight, we first split the genome into approximately LD-independent regions, and from each region, selected the variant with the lowest p-value (p-value < 5×10^-6^). For each of these suggestive variants (87 maternal and 108 fetal), we calculated the relative difference in effect size before and after conditioning as follows,

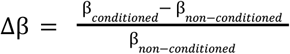

A negative Δβ suggests that adjusting for gestational duration reduces the effect size.

To test for robustness of results, we repeated the same analysis for genome-wide significant variants classified as having a maternal only effect on birth weight (n = 31) or a fetal only effect on birth weight (n = 62) ^9^. For such variants, we applied the same method as above, multi-trait COJO analysis, and a linear regression model with and without adjusting for gestational duration in individual level data from Iceland (genetic dosage, n mothers = 32,511 and n fetuses = 16,387) and Norway (MoBa, parental transmitted and non-transmitted alleles, n = 21,060 parent-offsprings). For individual level data, we fitted the following two linear models:

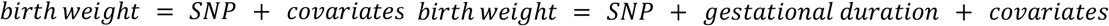

Where SNP is the genotype dosage in Icelandic data for the maternal genome or the fetal genome, and the maternal non-transmitted allele and paternal transmitted allele for maternal and fetal SNPs, respectively. Covariates included the first six principal components and batch for MoBa data. By employing different data sets, and different genetic information (dosage vs parental transmitted / non-transmitted alleles), allowed us to provide further evidence and robustness in our findings.

For all these analyses, we estimated statistical significance of differences between the effect estimates from the two models applying a paired Wilcoxon rank-sum test.

We estimated the heritability of the birth weight GWAS before and after conditioning for gestational duration using LD-score regression^65^. This method estimates polygenic heritability as the variance explained by common genetic variants (autosomal MAF>= 0.01). To test the significance of differences between heritability estimates before and after conditioning, we calculated a z-scored as follows ^66^,

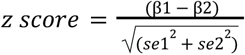

where β1 is the non-conditioned estimate, β2 the conditioned estimate, and *se*1 and *se*2 the standard errors for the non conditioned and conditioned estimates, respectively.

### Mendelian randomization

We performed Mendelian randomization to study the effects of gestational duration (maternal) on birth weight (maternal) and the effects of fetal growth (fetal effect on birth weight) and sex hormones on gestational duration.

To study the effect of gestational duration on birth weight, we employed two-sample Mendelian randomization. The 24 index SNPs (22 autosomal SNPs) from the present gestational duration meta-analysis and the effect sizes from the parental transmitted and non-transmitted alleles analysis were used to instrument gestational duration; birth weight was instrumented using summary statistics from a previous GWAS of offspring’s birth weight with minimal adjustment by gestational duration (< 15% of samples) ^9^. While there is a recent publication on fetal growth ^10^, this analysis was largely adjusted for gestational duration (> 40% of samples). To avoid confounding due to the correlation between the maternal and fetal genomes, we used summary statistics derived using a weighted linear model ^9^. This allowed us to obtain quasi-unbiased estimates for the fetal effects on birth weight (adjusting for the maternal effect):

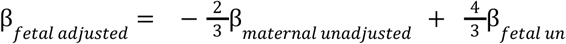

and for the maternal effects on birth weight (adjusting for the fetal effect):

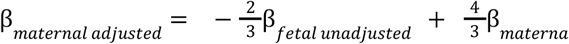

To obtain a causal estimate, we performed an inverse-variance weighted analysis with standard errors calculated using the delta method ^67^. We assessed the impact of horizontal pleiotropy on the causal estimate with MR-Egger regression. The intercept was used to determine whether the average pleiotropic effect is not statistically different from zero (p-value > 0.100). In such cases, the inverse-variance weighted method estimate is a consistent estimate of the causal effect ^68^. Whenever the MR-Egger intercept is significantly different from 0, we report the estimate from the MR-Egger analysis. Both inverse-variance weighted method and MR-Egger regression were performed on R using the MendelianRandomization package ^69^.

We assessed the effect of sex hormones (testosterone, SHBG and CBAT) on gestational duration using two-sample Mendelian randomization and instrumenting the hormones using a polygenic score for the parental transmitted and non-transmitted alleles. For each sex hormone, we obtained a list of independent SNPs genome-wide associated with these traits (**Supplementary Table 9**) by performing GWAS clumping (R^2^ > 0.001) using the following PLINK command:

plink --bfile <1000 Genomes> --clump {GWAS summary statistics} --clump-r2 0.001--clump-kb 1000 --clump-p1 5e-8 --clump-p2 1e-5

We also used a set of SNPs associated with testosterone, but with no aggregated effects on SHBG, as clustered in ^28^. Such variants were used as instrumental variables in the two-sample Mendelian randomization analysis and to construct the polygenic score for the parental transmitted and non-transmitted alleles. The current meta-analysis results were employed as outcome for the two-sample Mendelian randomization analysis (inverse-variance weighted and MR-Egger). We subsequently constructed the polygenic score for the maternal transmitted and non-transmitted alleles and the paternal transmitted alleles in 46,105 parent-offsprings from Iceland and Norway. We estimated the effects of the maternal non-transmitted (MnT_PGS_) and transmitted (MT_PGS_) and paternal transmitted (PT_PGS_) alleles polygenic score using the following linear model:

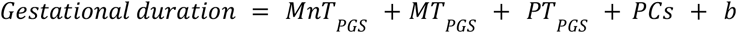

Again, effects from each of the three data sets (Iceland, MoBa and HUNT) were combined using fixed-effect inverse-variance weighted meta-analysis.

To understand the impact of fetal growth on gestational duration, we used individual genetic data from 35,280 (ultrasound-gestational duration) and 48,741 (last menstrual period-gestational duration) parent-offsprings from Iceland, the MoBa cohort and HUNT. To instrument fetal growth, we used 68 SNPs with fetal only effect on birth weight as classified in Warrington et al. using Structural Equation Modeling ^9^. Based on these 68 SNPs, we constructed a fetal growth polygenic score for the parental transmitted and non-transmitted alleles and regressed these on gestational duration (estimated by ultrasound or last menstrual period, separately). We estimated the effects of the maternal non-transmitted (MnT_PGS_) and transmitted (MT_PGS_) and paternal transmitted (PT_PGS_) alleles polygenic score using the following linear model:

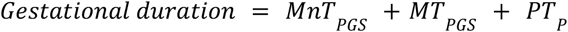

Effect estimates from each of the three data sets (Iceland, MoBa and HUNT) were pooled using fixed-effects inverse-variance weighted meta-analysis.

### Testing for maternal-fetal pleiotropic effects on gestational duration and birth weight

We further investigated what are the fetal effects on birth weight for the maternal gestational duration increasing alleles, and the maternal effects on gestational duration for the fetal birth weight increasing alleles. To study this, we borrowed the inverse-variance weighted analysis from Mendelian randomization, but using the effects of two distinct genomes, the maternal and fetal. We caution that this should not be interpreted under a causal framework.

To understand what the maternal gestational duration-raising alleles do to birth weight when present in the fetus, we used the effect sizes and standard errors of the parental transmitted and non-transmitted alleles for the 22 autosomal index SNPs on gestational duration and assessed its effects on the same SNPs with a fetal only effect on birth weight. To understand what the fetal birth weight-raising alleles do to gestational duration when present in the mother, we used the effect sizes and standard errors of 68 autosomal SNPs associated with fetal effects on birth weight and the effect sizes and standard errors from the current maternal GWAS of gestational duration.

### Evolutionary analysis

To examine the evolutionary history of three regions identified in the GWAS meta-analysis, we ran the significant variants through the MOSAIc pipeline ^23^. This pipeline is designed to detect enrichment in evolutionary signals using a variety of sequence-based metrics of selection. The sequence based evolutionary measures used in this method include: 1) Beta Score which detects balanced polymorphisms to infer balancing selection ^70^, 2) ARGWEAVE uses ancestral recombination graphs to infer the evolutionary origin of regions ^71^, 3) GERP uses sequence conservation to infer positive and negative selection ^72^, 4) LINSIGHT uses sequence conservation to infer positive and negative selection ^73^, 5) phastCONS100 uses sequence conservation to infer positive and negative selection, 6) PhyloP uses substitution rate to infer positive and negative selection, 7) iES uses haplotype homozygosity to infer positive selection, 8) XPEHH uses haplotype homozygosity to detect population-specific positive selection, and 9) Fst uses population differentiation to infer local adaptation.

Variants from the GWAS that passed a significance threshold (p-value <1×10^-8^) were clumped into regions using PLINK such that the clumps of variants had an R^2^ >0.9 and were within 500 kb. We then obtained 5,000 control variants matched on variant count, LD structure and minor allele frequency. The evolutionary metrics were obtained for all variants, and the maximum value was extracted for analysis. Finally, the evolutionary metrics were also obtained for the control variants and further used to create a background distribution. Then a z-score and p-value were produced for each experimental genomic region compared to its unique background distribution.

### Variant annotation

Variants were annotated using Ensembl’s Variant Effect Predictor (hg19) command line tool ^74^. Physical coordinates of protein coding genes were obtained from the UCSC Table Browser ^75^, and were matched to the index SNPs using bedtools v2.29.2 ^76^.

## Supporting information

Supplementary Test

## Data Availability

Cohorts should be contacted individually for access to raw genotype data, as each cohort has different data access policies. Summary statistics from the meta-analysis excluding 23andMe and the summary statistics of the top 10,000 SNPs for each phenotype will be made available at the EGG website (https://egg-consortium.org/). Access to the full set, including 23andMe results, can be obtained after approval from 23andMe is presented to the corresponding author or by completion of a Data Transfer Agreement (https://research.23andme.com/dataset-access/), which exists to protect the privacy of 23andMe participants. Access to the Danish National Birth Cohort (phs000103.v1.p1), Hyperglycemia and Adverse Pregnancy Outcome (phs000096.v4.p1), and Genomic and Proteomic Network (phs000714.v1.p1) individual-level phenotype and genetic data can be obtained through dbGaP Authorized Access portal (https://dbgap.ncbi.nlm.nih.gov/dbgap/aa/wga.cgi?page=login). The informed consent under which the data or samples were collected is the basis for determining the appropriateness of sharing data through unrestricted-access databases or NIH-designated controlled-access data repositories. The summary statistics used in this publication other than the one generated are available at the following links: fetal GWAS of gestational duration (https://egg-consortium.org/), fetal and maternal GWAS of gestational duration (https://egg-consortium.org/), miscarriage (http://www.geenivaramu.ee/tools/misc_sumstats.zip), age at first birth, oestradiol (women), endometriosis, number of live births and age at menarche (http://www.nealelab.is), age at menopause (https://www.reprogen.org), testosterone (women) 49, SHBG, testosterone and CBAT (https://doi.org/10.6084/m9.figshare.c.5304500.v1), pelvic organ prolapse and leiomyome of the uterus (https://www.finngen.fi/fi), polycystic ovary syndrome (https://www.repository.cam.ac.uk/handle/1810/283491 and https://www.finngen.fi/fi) and pre-eclampsia (European Genome-phenome Archive, https://ega-archive.org, EGAD00010001984). Pan-UK Biobank data is available at https://pan.ukbb.broadinstitute.org/.
For pre-computed LD scores for European populations (https://data.broadinstitute.org/alkesgroup/LDSCORE/eur_w_ld_chr.tar.bz2). g:Profiler, https://biit.cs.ut.ee/gprofiler/gost.

## Data availability

Cohorts should be contacted individually for access to raw genotype data, as each cohort has different data access policies. Summary statistics from the meta-analysis excluding 23andMe and the summary statistics of the top 10,000 SNPs for each phenotype will be made available at the EGG website (https://egg-consortium.org/). Access to the full set, including 23andMe results, can be obtained after approval from 23andMe is presented to the corresponding author or by completion of a Data Transfer Agreement (https://research.23andme.com/dataset-access/), which exists to protect the privacy of 23andMe participants. Access to the Danish National Birth Cohort (phs000103.v1.p1), Hyperglycemia and Adverse Pregnancy Outcome (phs000096.v4.p1), and Genomic and Proteomic Network (phs000714.v1.p1) individual-level phenotype and genetic data can be obtained through dbGaP Authorized Access portal (https://dbgap.ncbi.nlm.nih.gov/dbgap/aa/wga.cgi?page=login). The informed consent under which the data or samples were collected is the basis for determining the appropriateness of sharing data through unrestricted-access databases or NIH-designated controlled-access data repositories. The summary statistics used in this publication other than the one generated are available at the following links: fetal GWAS of gestational duration (https://egg-consortium.org/), fetal and maternal GWAS of gestational duration (https://egg-consortium.org/), miscarriage (http://www.geenivaramu.ee/tools/misc_sumstats.zip), age at first birth, oestradiol (women), endometriosis, number of live births and age at menarche (http://www.nealelab.is), age at menopause (https://www.reprogen.org), testosterone (women) ^60^, SHBG, testosterone and CBAT (https://doi.org/10.6084/m9.figshare.c.5304500.v1), pelvic organ prolapse and leiomyome of the uterus (https://www.finngen.fi/fi), polycystic ovary syndrome (https://www.repository.cam.ac.uk/handle/1810/283491 and https://www.finngen.fi/fi) and pre-eclampsia (European Genome-phenome Archive, https://ega-archive.org, EGAD00010001984). Pan-UK Biobank data is available at https://pan.ukbb.broadinstitute.org/. For pre-computed LD scores for European populations (https://data.broadinstitute.org/alkesgroup/LDSCORE/eur_w_ld_chr.tar.bz2), and for multi-tissue gene expression pre-computed stratified LD scores (https://alkesgroup.broadinstitute.org/LDSCORE/LDSC_SEG_ldscores/Multi_tissue_gene_expr_1000Gv3_ldscores.tgz). eQTL data from GTEx is available at https://gtexportal.org/home/ and from endometrium at http://reproductivegenomics.com.au/shiny/endo_eqtl_rna/. Protein QTL data was obtained from https://www.omicscience.org/apps/pgwas/.

## Code availability

Code for this project has been structured using a Snakemake workflow ^77^ and is available at (https://github.com/PerinatalLab/metaGWAS).

## Acknowledgements

Full acknowledgements and funding statements are detailed in the Supplementary Text.

## Consortia

Consortia members are listed in the Supplementary Text.

## Contributions

The core working group comprised: P.S-N., C.F., V.S., M.V., J.C., P.N., L.J.M., R.M.F., S.J., G.Z., B.J. Performed analyses in their respective cohorts: P.S-N., C.F., M.V., J.C., T.L., A.L.L., A.A, D.W., B.B., L.S., M.C.B., A.M., M.W., F.L., C.B., C.A.W., G.M., R.N.B., J.P.B., G.T., O.T., G.S., H.X., D.F.G., S.R., D.S., B.F. Individual cohort designers and principal investigators included: V.S., J.B., J.J., A.M., M.E.G., EA.N., E.H., S.M.W., R.M., M.M., L.W., L.B., E.O., R.T.L., K.T., M.H., T.J., H.H., B.M.S., L.S., M.M., D.S., H.U., C.P., M.N., J.A.C., A.H.S., P.M., O.A.A., UT., S.F.G., E.Q., C.E.P., M.H., G.M.H., M.I.McC., D.A.L., H.S.N., R.M., K.H., K.S., B.F., L.J.M., R.M.F., S.J., G.Z., B.J. Sample collection, phenotyping and/ or genotyping was performed by: V.S., J.B., J.J., O.H., C.A.W., G.T., M.E.G., S.R.O., D.M., E.A.N., E.H., A.S., C.A., A.T.H., M.M., L.W., L.B., E.O., O.B.P., C.E., E.S., K.T., H.U., M.H., T.J., H.H., B.M.S., L.S., T.S., C.P., J.W., A.H.S., P.M., O.A.A., U.T., S.F.G., E.Q., C.E.P., M.H., G.M.H., M.J., M.I.McC., D.A.L., R.M., K.H., B.F., P.N.,L.J.M., G.Z., B.J.

