## Supplementary Test for "Genetic effects on the timing of parturition and links to fetal birth weight"

Steffen Andersen[81], Karina Banasik[8], Søren Brunak[8], Kristoffer Burgdorf[28], Maria Didriksen[28], Khoa Manh Dinh[43], Christian Erikstrup[43], Daniel Gudbjartsson[2], Thomas F. Hansen[82], Henrik Hjalgrim[12,83], Gregor Jemec[84], Poul Jennum[85], Pär I. Johansson[28], Margit A. Larsen[28], Susan Mikkelsen[43], Kasper R. Nielsen[86], Mette Nyegaard[61], Sisse R. Ostrowski[28], Ole B. Pedersen[41], Kari Stefansson[2], Hreinn Stefansson[2], Susanne Sækmose[41], Erik Sørensen[28], Unnur Thorsteinsdottir[2], Mie T. Brun[87], Henrik Ullum[88], Thomas Werge[89]

[81]Department of Finance, Copenhagen Business School, Copenhagen, Denmark
[82]Danish Headache Center, Department of Neurology, Copenhagen University Hospital, Rigshospitalet â “ Glostrup, Glostrup, Denmark
[83]Centre for Cancer Research, Danish Cancer Society, Copenhagen, Denmark
[84]Department of Dermatology, Zealand University hospital - Roskilde, Roskilde, Denmark
[85]Department of clinical neurophysiology, University of Copenhagen, Copenhagen, Denmark
[86]Department of Clinical Immunology, Aalborg University Hospital, Aalborg, Denmark
[87]Department of Clinical Immunology, Odense University Hospital, Odense, Denmark
[88]Statens Serum Institute, Copenhagen, Denmark
[89]Institute of Biological Psychiatry Mental Health Centre, Sct. Hans, Copenhagen University Hospital â “ Roskilde, Roskilde, Denmark

#### Estonian Biobank Research Team

Andres Metspalu[6], Lili Milani[6], Tõnu Esko[6], Mari Nelis[6], Georgi Hudjashov[6]

#### Early Growth Genetics

Emma Ahlqvist[90], Tarunveer S. Ahluwalia[91,92,93], Jonas Bacelis[94], Robin N. Beaumont[95], Thomas Bond[96], Dorret I. Boomsma[97], Jonathan P. Bradfield[98,26], Lachlan Coin[99], Cyrus Cooper[100], Diana L. Cousminer[101], John A. Curtin[102], Adnan Custovic[103], Felix R. Day[104],

Maneka De Silva[70], Paul Elliott[96], João Fadista[12], Bjarke Feenstra[105], Romy Gaillard[106], Frank Geller[105], Niels Grarup[107], Leif Groop[108,90], Monica Guxens[109,110,111], Dexter Hadley[112], Johannes Hebebrand[113], Ã˜yvind Helgeland[15,114,115], Tine B. Henrisksen[116], Anke Hinney[113], Joel N. Hirschhorn[117], Marie-France Hivert[20,118,119], Berthold Hocher[120,121], John W. Holloway[122], Momoko Horikoshi[123], Jouke-Jan Hottenga[124,125,126], Stefan Johansson[114,127], Heidi J. Kalkwarf[128], Sailesh Kotecha[129], Zoltán Kutalik[130,131,132], Joan M. Lappe[133], Alexandra M. Lewin[134], Cecilia M. Lindgren[16,135], Reedik Mägi[136], Per Magnus[137], Niina S. McCarthy[138], Camilla S. Morgen[139], Louis J. Muglia[140,141,142,143], Ronny Myhre[144], Ioanna Ntalla[145], Sharon E. Oberfield[146], Emily Oken[20], Niina Pitkänen[147], Beate St Pourcain[148,149], Rashmi B. Prasad[90,150], Rebecca M. Reynolds[151], Rebecca C. Richmond[149,14], Alina Rodriguez[152,153], Rany Salem[154,155,156,157], Theresia M. Schnurr[107], John A. Shepherd[158], Angela Simpson[102], Line Skotte[12], Eric A. Steegers[159], Jordi Sunyer[109,110,111,160], Elisabeth Thiering[161,162], Jessica Tyrrell[95,163], Marc Vaudel[3,15], Carol A. Wang[164], Nicole M. Warrington[165,166,167], William J. Watkins[129], H-Erich Wichmann[168,169,170], Babette S. Zemel[171], Ge Zhang[172,141,142], Andrew T. Hattersley[173], Bo Korner[1,15], Deborah A Lawlor[174,175], David Evans[99,166,174], Ellen A. Nohr[176], Eleftheria Zeggini[177,178], Elina Hypponen[179,32], Struan F. Grant[180,181,182,183,184], Janine Felix[185,186], Jeff Murray[187], James F. Wilson[188], Jun Liu[189], Linda S. Adair[190], Karen L. Mohlke[191], Ronald C. Ma[192,193,194], Sylvain Sebert[195], Tanja Vrijkotte[196], Thorkild I. Sørensen[197], Torben Hansen[198], Vincent Jaddoe[185,186], Mariona Bustamante[109,110,111], Martine Vrijheid[109,110,111], Pol Sole-Navais[1], Izzuddin Aris[20], Sara E. Stinson[198], Jens-Christian Holm[199], Justiina Ronkainen[70], Christopher Flatley[94], William Schierding[200], Alice E. Hughes[201], Joseph T. Glessner[202,203], Triinu Peters[113], Xiaoping Wu[105], Jeffrey J. Beck[204,205], Yee-Ming Chan[206,207], Jia Zhu[206], Roseann E. Peterson[208], Brandon Lim[95]

[90]Department of Clinical Sciences, Diabetes and Endocrinology, Lund University Diabetes Centre, Malmö, Sweden

[91]COPSAC, Copenhagen Prospective Studies on Asthma in Childhood, Herlev and Gentofte Hospital, University of Copenhagen, Copenhagen, Denmark

[92]Steno Diabetes Center Copenhagen, Herlev, Denmark

[93]The Bioinformatics Center, Department of Biology, University of Copenhagen, Copenhagen, Denmark

[94]Department of Obstetrics and Gynecology, Sahlgrenska University Hospital, Gothenburg, Sweden

[95]Institute of Biomedical and Clinical Science, University of Exeter Medical School, University of Exeter, Royal Devon and Exeter Hospital, Exeter, UK

[96]Department of Epidemiology and Biostatistics, School of Public Health, Imperial College London, London, UK

[97]Netherlands Twin Register, Department of Biological Psychology, VU Universiteit, Amsterdam, the Netherlands

[98]Center for Applied Genomics, Children's Hospital of Philadelphia, Philadelphia, United States

[99]Institute for Molecular Bioscience, University of Queensland, Brisbane, Australia

[100]MRC Lifecourse Epidemiology Unit, Faculty of Medicine, University of Southampton, Southampton, UK

[101]Genomic Sciences, Glaxosmithkline, Collegeville, PA, USA

[102]Division of Infection Immunity and Respiratory Medicine, School of Biological Sciences, The University of Manchester, Manchester Academic Health Science Centre, and Manchester University NHS Foundation Trust, Manchester, UK

[103]Department of Paediatrics, Imperial College, London, UK

[104]MRC Epidemiology Unit, University of Cambridge School of Clinical Medicine, Cambridge, UK
[105]Department of Epidemiology Research, Statens Serum Institute, Copenhagen, Denmark
[106]Department of Epidemiology, Erasmus MC, University Medical Center Rotterdam, Rotterdam, the Netherlands
[107]Novo Nordisk Foundation Center for Basic Metabolic Research, Section of Metabolic Genetics, Faculty of Health and Medical Sciences, University of Copenhagen, Copenhagen, Denmark
[108]Institute for Molecular Medicine, Finland (FIMM), University of Helsinki, Helsinki, Finland
[109]ISGlobal, Institute for Global Health, Barcelona, Spain
[110]CIBER de Epidemiologia y Salud Publica (CIBERESP), Madrid, Spain
[111]Universitat Pompeu Fabra (UPF), Barcelona, Spain
[112]Department of Pediatrics, University of California San Francisco School of Medicine, San Francisco, USA
[113]Department of Child and Adolescent Psychiatry, Psychosomatics and Psychotherapy, University Hospital Essen, University of Duisburg-Essen, Essen, Germany
[114]KG Jebsen Center for Diabetes Research, Department of Clinical Science, University of Bergen, Bergen, Norway
[115]Department of Pediatrics, Haukeland University Hospital, Bergen, Norway
[116]Department of Clinical Medicine - Department of Paediatrics, Aarhus University Hospital, Aarhus N, Denmark
[117]Broad Institute of MIT and Harvard, Cambridge, USA
[118]Diabetes Center, Massachusetts General Hospital, Boston, USA
[119]Department of Medicine, Universite de Sherbrooke, Sherbooke, Canada
[120]Institute of Nutritional Science, University of Potsdam, Nuthetal, Germany
[121]The First Affiliated Hospital of Jinan University, Guangzhou, China
[122]Human Development & Health, Faculty of Medicine, University of Southampton, Southampton, UK
[123]RIKEN, Centre for Integrative Medical Sciences, Laboratory for Genomics of Diabetes and Metabolism, Yokohama, Japan
[124]Department of Biological Psychology, Vrije Universiteit Amsterdam, Amsterdam, the Netherlands
[125]Netherlands Twin Register, Department of Biological Psychology, VU University, Amsterdam, the Netherlands
[126]Amsterdam Public Health, Amsterdam, the Netherlands
[127]Center for Medical Genetics and molecular Medicine, Haukeland University Hospital, Bergen, Norway
[128]Division of Gastroenterology, Hepatology and Nutrition, Cincinnati Children's Hospital Medical Center, Cincinnati, USA
[129]Department of Child Health, School of Medicine, Cardiff University, Cardiff, UK
[130]Institute of Primary Care and Public Health, University of Lausanne, Lausanne, Switzerland
[131]Department of Computational Biology, University of Lausanne, Lausanne, Switzerland
[132]Swiss Institute of Bioinformatics, Lausanne, Switzerland
[133]Division of Endocrinology, Department of Medicine, Creighton University, Omaha, USA
[134]Department of Epidemiology and Biostatistics, MRC PHE Centre for Environment & Health, School of Public Health, Imperial College London, London, UK
[135]Li Ka Shing Centre for Health Information and Discovery, The Big Data Institute, University of Oxford, Oxford, UK
[136]Estonian Genome Center, University of Tartu, Tartu, Estonia
[137]Norwegian Institute of Public Health, Oslo, Norway
[138]Centre for Genetic Origins of Health and Disease (GOHaD), The University of Western Australia, Crawley, Australia

[139]Department of Public Health, Section of Epidemiology, Faculty of Health and Medical Sciences, University of Copenhagen, Copenhagen, Denmark
[140]Center for Prevention of Preterm Birth, Perinatal Institute, Cincinnati Children's Hospital Medical Center, Department of Pediatrics, University of Cincinnati College of Medicine, Cincinnati, USA
[141]March of Dimes Prematurity Research Center Ohio Collaborative, Cincinnati, USA
[142]Human Genetics Division, Cincinnati Children ™s Hospital Medical Center, Cincinnati, USA
[143]President, Burroughs-Wellcome Fund
[144]Department of Genes and Environment, Division of Epidemiology, Norwegian Institute of Public Health, Oslo, Norway
[145]William Harvey Research Institute, Barts and the London School of Medicine and Dentistry, Queen Mary University of London, London, UK
[146]Division of Pediatric Endocrinology, Diabetes, and Metabolism, Department of Pediatrics, Columbia University Medical Center, New York, USA
[147]Research Centre of Applied and Preventive Cardiovascular Medicine, University of Turku, Turku, Finland
[148]Max Planck Institute for Psycholinguistics, Nijmegen, the Netherlands
[149]Medical Research Council Integrative Epidemiology Unit at the University of Bristol, Bristol, UK
[150]Institute of Molecular Medicine, Helsinki, Finland
[151]Centre for Cardiovascular Science, Queen's Medical Research Institute, University of Edinburgh, Edinburgh, UK
[152]Department of Epidemiology and Biostatistics, MRC PHE Centre for Environment & Health, School of Public Health, Imperial College London, London, UK
[153]Department of Psychology, Mid Sweden University, Östersund, Sweden
[154]Division of Endocrinology, Department of Medicine, Boston Children's Hospital, Boston, USA
[155]Department of Genetics, Harvard Medical School, Boston, USA
[156]Center for Basic and Translational Obesity Research, Boston Children's Hospital, Boston, USA
[157]Program in Medical and Population Genetics, Broad Institute of Harvard and MIT, Cambridge, USA
[158]Department of Radiology, University of California San Francisco, San Francisco, USA
[159]Department of Obstetrics and Gynecology, Erasmus MC, University Medical Center, Rotterdam, the Netherlands
[160]IMIM (Hospital del Mar Medical Research Institute), Barcelona, Spain
[161]Institute of Epidemiology, Helmholtz Zentrum Munchen - German Research Center for Environmental Health, Neuherberg, Germany
[162]Division of Metabolic and Nutritional Medicine, Dr. von Hauner Children's Hospital, University of Munich Medical Center, Munich, Germany
[163]European Centre for Environment and Human Health, University of Exeter, Truro, UK
[164]School of Medicine and Public Health, Faculty of Medicine and Health, The University of Newcastle, Callaghan, Australia
[165]Institute of Molecular Biosciences, University of Queensland, Brisbane, Australia
[166]University of Queensland Diamantina Institute, University of Queensland, Brisbane, Australia
[167]K.G. Jebsen Center for Genetic Epidemiology, Department of Public Health and Nursing, Norwegian University of Science and Technology, Norway
[168]Institute of Medical Informatics, Biometry and Epidemiology, Chair of Epidemiology, Ludwig Maximilians University, Munich, Germany
[169]Helmholtz Center Munich, Institute of Epidemiology, Neuherberg, Germany
[170]Institute of Medical Statistics and Epidemiology, Technical University Munich, Munich, Germany
[171]Division of Gastroenterology, Hepatology and Nutrition, The Childrens Hospital of Philadelphia, Philadelphia, USA

[172]Division of Human Genetics, Cincinnati Children's Hospital Medical Center, Center for Prevention of Preterm Birth, Perinatal Institute, Cincinnati Children's Hospital Medical Center, Department of Pediatrics, University of Cincinnati College of Medicine, Cincinnati, USA
[173]University of Exeter Medical School, College of Medicine and Health, University of Exeter, Exeter, UK
[174]MRC integrative Epidemiology Unit, University of Bristol, UK
[175]Population Health Science, Bristol Medical School, Bristol University, UK
[176]Department of Clinical Research, Research Unit for Obstetrics and Gynecology, University of Southern Denmark, Odense, Denmark
[177]Institute of Translational Genomics, Helmholtz Zentrum Munchen â “ German Research Center for Environmental Health, 85764 Neuherberg, Germany
[178]Technical University of Munich (TUM) and Klinikum Rechts der Isar, TUM School of Medicine, Ismaninger Str. 22, 81675 Munich, Germany
[179]Australian Centre for Precision Health, Unit of Clinical and Health Sciences, University of South Australia, Adelaide, Australia
[180]Division of Human Genetics, The Children's Hospital of Philadelphia, Philadelphia, USA
[181]Division of Endocrinology and Diabetes, The Children's Hospital of Philadelphia, Philadelphia, USA
[182]Department of Pediatrics, University of Pennsylvania Perelman School of Medicine, Philadelphia, United States
[183]Department of Genetics, University of Pennsylvania Perelman School of Medicine, Philadelphia, United States
[184]Center for Applied Genomics, Childrens Hospital of Philadelphia, Philadelphia, United States
[185]The Generation R Study Group, Erasmus MC, University Medical Center Rotterdam, Rotterdam, the Netherlands
[186]Department of Pediatrics, Erasmus MC, University Medical Center Rotterdam, Rotterdam, the Netherlands
[187]Dept of Pediatrics, University of Iowa, Iowa City, IA 52240 USA
[188]Centre for Global Health Research, Usher Institute, University of Edinburgh, Teviot Place, Edinburgh, EH8 9AG, Scotland
[189]Nuffield Department of Population Health, Univeristy of Oxford, Oxford, UK
[190]Department of Nutrition, University of North Carolina, Chapel Hill, NC, United States
[191]Department of Genetics, University of North Carolina, Chapel Hill, NC, United States
[192]Department of Medicine and Therapeutics, The Chinese University of Hong Kong
[193]Li Ka Shing Institute of Health Sciences, The Chinese University of Hong Kong, Hong Kong, China
[194]Hong Kong Institute of Diabetes and Obesity, The Chinese University of Hong Kong, Hong Kong, China
[195]Research Unit of Population Health, Faculty of Medicine, University of Oulu, Oulu, Finland
[196]Department of Public and Occupational Health, Amsterdam University Medical Center, University of Amsterdam, Amsterdam
[197]Department of Public Health and Novo Nordisk Foundation Center for Basic Metabolic Research, Faculty of Health and Medical Sciences, University of Copenhagen, Denmark
[198]Novo Nordisk Foundation Center for Basic Metabolic Research, Faculty of Health and Medical Sciences, University of Copenhagen, Denmark
[199]The Childrens Obesity Clinic, accredited European Centre for Obesity Management, Department of Pediatrics, Holbäk Hospital, Denmark
[200]Liggins Institute, University of Auckland, Auckland, NZ
[201]Institute of Biomedical and Clinical Science, University of Exeter Medical School, Exeter, UK
[202]Department of Pediatrics, Children ™s Hospital of Philadelphia, 3401 Civic Center Blvd, Philadelphia, PA 19104, USA.

[203]Department of Pediatrics, Division of Human Genetics, Perelman School of Medicine, 3400 Civic Center Blvd, Philadelphia, PA 19104, USA.
[204]Avera Institute for Human Genetics, Avera McKennan Hospital and University Health Center, Sioux Falls, South Dakota, USA
[205]Department of Psychiatry, University of South Dakota Sanford School of Medicine, South Dakota, USA
[206]Division of Endocrinology, Department of Pediatrics, Boston Children's Hospital
[207]Department of Pediatrics, Harvard Medical School
[208]Department of Psychiatry and Behavioral Sciences, College of Medicine, State University of New York Downstate Health Sciences University, Brooklyn, New York, USA

### Description of participating cohorts

#### 23andMe

Genome-wide summary statistics from a previously published GWAS on gestational duration and preterm delivery were used [1]. Briefly, participants in the research program of 23andMe, a personal genomics and biotechnology company provided written informed consent and answered surveys online according to a human-subjects protocol approved by Ethical and Independent Review Services (www.eandireview.com). Unrelated women of European ancestry self-reported the gestational duration of their first live singleton birth. Women with a medical indication for preterm delivery were excluded. Preterm-birth status was determined on the basis of dichotomization of gestational duration (preterm, <37 weeks; term, ≥37 weeks).
DNA extraction and genotyping were performed by the National Genetics Institute. The analysis was restricted to 43,568 women. Genotype data were imputed against the reference haplotypes of phase 1 of the 1000 Genomes Project [2].

Linear regression was used to estimate genetic associations with gestational duration and logistic regression to test such associations with preterm birth. The maternal age and the top five principal components were included as covariates.

#### Avon Longitudinal Study of Parents and Children

The Avon Longitudinal Study of Parents and Children (ALSPAC) is a longitudinal birth cohort, which has been described in detail elsewhere [3,4]. In short, pregnant women resident in Avon, UK with expected dates of delivery 1st April 1991 to 31st December 1992 were invited to take part in the study. The initial number of pregnancies enrolled is 14,541. Of these initial pregnancies, there were a total of 14,676 foetuses, resulting in 14,062 live births and 13,988 children who were alive at 1 year of age. Ethical approval for the study was obtained from the Avon Longitudinal Study of Parents and Children Law and Ethics Committee (IRB# 00003312) and the Local Research Ethics Committees

(Bristol and Weston, Southmead, and Frenchay Health Authorities). Written informed consent was obtained from all adult participants in the study. Consent for biological samples has been collected in accordance with the Human Tissue Act (2004). Please note that the study website contains details of all the data that is available through a fully searchable data dictionary and variable search tool (http://www.bristol.ac.uk/alspac/researchers/our-data/).

**Born in Bradford**

The Born in Bradford (BiB) cohort study was established in 2007 as detailed elsewhere [5]. All women booked for delivery at the Bradford Royal Infirmary are offered an oral glucose tolerance test (OGTT) at 26–28 weeks gestation. On attendance at the OGTT clinic, full consent was obtained for recruitment to BiB, and the woman was invited to complete an interviewer-administered questionnaire. Between March 2007 and November 2010, >80% of the women who attended the OGTT took up the invitation, and we recruited 12,453 women with 13,776 pregnancies to the cohort. Ethics approval has been obtained for the main platform study and was provided by the Bradford Local NHS Research Ethics Committee (ref 06/ Q1202/48).

**British 1958 Birth Cohort (1958BC-T1DGC and 1958BC-WTCCC2)**

The 1958 British Birth Cohort (1958BC) consists of all born during one week in March 1958 in England, Scotland, and Wales (n=17,638) [6]. Participants have been followed-up from birth and at age 45 years, 11,971 cohort members who had not died or emigrated, were invited to a biomedical assessment. In this survey 9,377 participants took part in clinical assessments and DNA collection. Ethical approval for the 45y survey was obtained from South East Multi-centre Research Ethics Committee (ref. 01/1/44) and the Joint UCL/UCLH Committees on the Ethics of Human Research (Committee A) Ref: 08/H0714/40.

Genome-wide data has been obtained through two sub-studies, both using the 1958BC as a control population. First, 3000 DNA samples were randomly selected and genotyped on the Affymetrix SNP 6.0 platform as part of the Wellcome Trust Case Control Consortium (WTCCC2) [7]. Secondly, 2,592 participants from the 1958BC were used as controls for a Type 1 diabetes case-control study (T1DGC). DNA samples were genotyped through the JDRF/WT Diabetes and Inflammation Laboratory (DIL) using the Illumina Infinium 550K chip [8]. Imputation was done in IMPUTE after quality control. For B58C-WTCCC2 quality control included SNP exclusions (Minor allele frequency (MAF) < 0.01, HWE P-value <1 x 10-20, call rate < 0.98, genotype plate association <1 x 10-5), and sample exclusions (heterozgygosity, call rate, relatedness, non-European ancestry and sex discrepancy). For B58C-T1DGC, criteria for SNP exclusions were MAF < 0.01, HWE

P-value <1 x 10-7, or SNP call rate < 0.95, and sample exclusions were made for heterozygosity, call rate, non-European ancestry and potential sex discrepancy.

Information on gestational duration was collected at the 33-year and 41-year follow-up surveys [9], when cohort members reported whether they had been pregnant and if so, what was the outcome of each pregnancy (miscarriage, abortion, stillbirth, livebirth). For live births, mothers provided information about birth weight. Information on gestational duration was asked by a question whether the child was born at term, or alternatively how many weeks in advance or late.

**Children’s Hospital of Philadelphia (CHOP)**

All subjects were consecutively recruited from the Greater Philadelphia area from 2006 to 2021 at the Children's Hospital of Philadelphia. Our study cohort consisted of children of European ancestry. All these participants had their blood drawn into an 8ml EDTA blood collection tube and were subsequently DNA extracted for genotyping. All subjects were biologically unrelated and were aged less than 18 years old at the time of blood collection. This study was approved by the Institutional Review Board of the Children's Hospital of Philadelphia. Parental informed consent was given for each study participant for both the blood collection and subsequent genotyping.

We performed high throughput genome-wide SNP genotyping, using the Illumina Infinium™ II HumanHap550 and Human610 BeadChip technology (Illumina, San Diego), at the Center for Applied Genomics at CHOP. We used 750ng of genomic DNA to genotype each sample, according to the manufacturer’s guidelines.

Samples were genotyped on a combination of the HumanHap 550 version 1, HumanHap 550 Version 3 and 610 Quad SNP chips. The SNPs in common with the three different chip versions used were the basis for all further analyses. The Sanger Imputation Server was used to impute ~40 million SNPs using the HRC 1.1 reference haplotypes.

**Danish Blood Donor Study**

The Danish Blood Donor Study (DBDS) is a prospective cohort of blood donors of whom the first of 116,000 individuals have been genotyped [10]. This study was performed under the reproductive health protocol (CVK-1805807). The DBDS cohort included 20,577 unique pregnancies identified from the Danish Medical Birth Registry (DMBR, 1,430, 485, 4,794 deliveries <259 days, <238 days, and >294 days, respectively) [11]. The DMBR covers all births since 1973 (both live- and still-births). Multi-fold births and still-births were excluded from the analysis. Furthermore, pregnancies with recorded obstetric complications: placental abruption, placenta previa, pre-eclampsia, eclampsia, polyhydramnios, oligohydramnios, placental insufficiency, cervical insufficiency, isoimmunization, breech presentation or maternal conditions: diabetes (Types I, II, gestational), hypertension, autoimmune diseases (SLE, RA, IBD, scleroderma), immunodeficiency states (HIV), abnormality of pelvic organs were excluded. Complications and maternal conditions were identified by linkage with the Danish National Patient Registry (DNPR). The DNPR covers all hospital admissions starting from 1977. DBDS samples were genotyped at deCODE Genetics using the Illumina Global Screening Array. Imputation was performed using a Northern European reference panel based on whole-genome sequencing. Quality control filtering required sample call rate of ≥0.98 and principal-component analysis was used to exclude ancestral outliers. We also performed SNP quality control (call rate of ≥0.98 in both cases and controls and HWE P of ≥1 × 10−6 in controls). Association testing was performed using BOLT-LMM [12](v2.3) or SAIGE (v0.36.3.3) [13] for the continuous and binary phenotypes, respectively, including year of birth, year of birth squared, and the first ten principal components.

**deCODE**

The study was approved by the National Bioethics Committee, Iceland (approval no. VSN-15-169) following evaluation of the Icelandic Data Protection Authority. We have obtained informed consent for all participants in this study who donated samples.

Information on gestational age comes from the Icelandic birth register, which includes 142,447 Icelanders born between 1982 and 2016 [14]. After excluding multiple births and infant deaths, we had information on gestational duration for 61,116 mothers. Mean birth year of mothers was 1969 (s.d. 25.1) and mean age at the birth of first child was 26.6 (s.d. 6.0).

Genotyping and imputation

Genotyping and imputation of the Icelandic samples were performed as previously described [15,16]. In short, we sequenced the whole genomes of 28,075 Icelanders using Illumina technology to a mean depth of at least 10X (median 32X). Genotypes were called using joint calling with the Genome Analysis Toolkit HaplotypeCaller (GATK version 3.4.07) [17]. Genotype calls were improved by using information about haplotype sharing, taking advantage of the fact that all sequenced individuals had also been chip-typed and long-range phased. About 33 million variants that passed the quality threshold were then imputed into 155,250 Icelanders, who had been genotyped with various Illumina SNP chips and their genotypes phased using long-range phasing [18,19]. Using genealogical information, the sequence variants were also imputed into 285,664 of their first- and second-degree relatives [15]. Out of 61,116 mothers included in the study, 37,397 were chip typed.

Association testing

We used logistic regression assuming an additive model in the case-control analysis to test for association between variants and disease, treating disease status as the response and expected genotype counts from imputation as covariates, and using likelihood ratio test to compute P-values. The model also included adjustment for additional covariates, i.e. county of birth, current age or age at death (first and second order terms included), availability of blood sample for the individual and an indicator function for the overlap of the lifetime of the individual with the time span of phenotype collection. To test the association of sequence variants with gestational duration we used a linear mixed model implemented in BOLT-LMM [12]. Prior to association analysis the measurements were adjusted for year of birth and age and, where appropriate, normalized to a standard normal distribution using rank based inverse normal transformation. All the analysis was done using software developed at deCODE genetics [15]. Only variants with imputation info over 0.8 and MAF > 0.01% were included. To account for inflation in test statistics due to cryptic relatedness and stratification, we applied the method of LD score regression [20], using an LD score estimated from the European ancestry samples in the 1000 Genomes Project, to estimate the inflation in the test statistics and adjusted all P-values accordingly.

**Danish National Birth Cohort - DNBC**

The DNBCGOYACASES, DNBCGOYACONTROLS, and DNBCPTD samples are sub-cohorts nested within the Danish National Birth Cohort (DNBC) [21]. The DNBC is a collection of data on 92,274 pregnant women recruited between 1996 and 2002, from their first antenatal visit to their general practitioner. Women participated in four telephone interviews (16 and 30 weeks gestation and 6 and 18 months after birth). They also provided two blood samples during pregnancy. Information about pregnancy outcomes for the DNBC participants was obtained from the Danish Medical Birth Register [11] and the Danish National Patient Register [22]. Gestational duration in this dataset was determined by a consensus algorithm combining all available information from multiple sources: self-reported date of last menstrual period, self-reported expected delivery date, and gestational age at birth registered in the Medical Birth Register and the National Patient Register.

The participants in the GOYA (Genomics of extremely Overweight Young Adults) study were drawn from the 67,863 women within the DNBC who provided information about prepregnancy BMI, gave birth to a live born infant and provided a blood sample during pregnancy [23]. A case sample of the 3.6% most obese women (DNBCGOYACASES; n=2451) was defined as those with the largest residuals from the regression of BMI on age and parity (all entered as continuous variables). The BMI for these women ranged from 32.6 to 64.4. From the remaining cohort we selected a random sample of similar size (DNBCGOYACONTROLS; n=2450). Genome-wide SNP array data for the GOYA participants were generated using the Illumina Human 610 Quad v1.0 BeadChip. Ethical approval was obtained from the Regional Scientific Ethical Committee of the Region of Mid Jutland and the study was also approved by the Danish Data Protection Agency.

The Danish National Birth Cohort - Preterm Delivery Study (DNBCPTD) is a case-control study [24] using mother-infant pairs to investigate genetic and environmental influences on spontaneous preterm birth. The study is nested within the DNBC; for case pairs delivery occurred before 37 weeks of gestation and for control pairs delivery occurred in gestational week 39, 40, or 41. Exclusion criteria were multiple deliveries, pregnancy complications such as placental abnormalities, preeclampsia/eclampsia, congenital abnormalities or stillbirth. Individuals were further required to be of Northern European ancestry. Genome-wide genotype data were obtained using the Illumina Human 660W Quad array and generated as part of the Gene Environment Association Studies (GENEVA) consortium [25]. Ethical approval was obtained from the Regional Scientific Ethical Committee of Copenhagen and the study was also approved by the Danish Data Protection Agency.

After genotyping, data from the DNBCGOYACASES, DNBCGOYACTRLS, and DNBCPTD samples were aligned to the forward strand and combined, including only overlapping SNPs between the Illumina Human 610-Quad and Human 660W Quad arrays. Data cleaning and QC steps were based on the following requirements for samples and SNPs to be included: sample missingness rate < 4%, heterozygosity within 3 standard deviations from the mean, variant missingness rate < 2%, minor allele frequency > 1%, A/T and C/G variants excluded, Hardy-Weinberg P value > 1e-06. Furthermore, principal components analysis jointly with data from the Human Genome Diversity Project was used to restrict the analysed sample to individuals of European ancestries, and individuals were excluded if they were related to another participant in the sample (corresponding to an IBD proportion >0.1875). The cleaned data containing 502,119 SNPs was phased using EAGLE v2.3 [26] and imputed to the Haplotype Reference Consortium release 1.1 [27] panel using Minimac3 [28]. After imputation, association testing was carried out in the DNBCGOYACASES (n=1610), DNBCGOYACTRLS (n=1912), and DNBCPTD (n=2226) cohorts separately using SNPTEST 2.5.2 [29].

For analyses of parental transmitted and non-transmitted data, data downloaded from the Database of Genotypes and Phenotypes (dbGaP) (phs000103.v1.p1) was used.

**Estonian Genome Center of the University of Tartu**

The Estonian Genome Center of the University of Tartu (EGCUT) is a population-based biobank with 51,515 participants in the first wave of data collection [30]. Data on diagnoses is obtained via periodical linking of the EGCUT dataset to national electronic databases and registries [30]. All EGCUT participants have signed a broad informed consent form and the study was conducted under the approval 234T-12 issued by the Ethics Review Committee of the University of Tartu.

**Exeter Family of Childhood Health (EFSOCH)**

The EFSOCH study [31] is a prospective study of children born between 2000 and 2004, and their parents, from a geographically defined region of Exeter, UK. All women gave informed consent and ethical approval was obtained from the local review committee. Gestational age at delivery was calculated based on the last menstrual period, or when that was unreliable or unavailable, dating by ultrasound scan was used.

Maternal and paternal DNA samples were extracted from parental blood samples obtained at the study visit (when the women were 28 weeks pregnant), and offspring DNA was obtained from cord blood at birth.

Genotyping of 2768 EFSOCH samples (n=969 mothers, 937 fathers and 862 children) was performed using the Illumina Infinium HumanCoreExome-24 array (n=551,839 SNPs/indels). Individual DNA samples with genotype call rate <98% were removed (n=50 individuals [1.8%]). SNPs were removed if they had call rates <95% (n=13,151 SNPs), showed evidence of deviation from Hardy-Weinberg equilibrium ($P<1x10^{-6}$; n=455 further SNPs), or had a minor allele frequency (MAF) <1% (n=257,289 further SNPs). Genotypically-derived sex information was compared with sex information in the phenotype file, and mismatched samples were excluded (n=13 individuals [0.61%]). Kinship was estimated using King [32]. Where evidence of labelling errors was clear, labels were updated, otherwise samples with kinship errors were excluded (n=22 individuals [0.79%]). Principal component analysis was performed to assess ancestry of the sample using flashPCA [33]. Outliers were defined as >4.56 SD from the cluster mean (defined using 1000 Genomes European data as the reference) and excluded (n=21 individuals [0.76%]). Imputation was performed using the Michigan imputation server and samples were imputed to the Haplotype Reference Consortium HRC v1.1 reference panel. SNPs were included in the analysis if imputation quality score was >0.4 and MAF ≥1%. After imputation, and once all exclusions had been made, 7,674,771 SNPs were available for analysis in 2662 individuals (939 mothers, 911 fathers and 812 children). Analyses were adjusted for the genotyping batch.

Ethical approval for the Exeter Family of Childhood Health was given by the North and East Devon (UK) Local Research Ethics Committee (approval number 1104), and informed consent was obtained from the parents of the newborns.

**FIN**

The Finnish dataset (FIN) was collected for a genetic study of spontaneous preterm birth [1,34]. Briefly, whole blood samples were collected from more than 1,600 mother/child pairs from the Helsinki (southern Finland) University Hospitals between 2004 and 2014.

All the studied samples are of Finnish descent. Crown-rump length at the first ultrasound screening between 10+ and 13 weeks was used to determine the gestational age. 2,962 blood samples from mothers and children were genotyped. After genotype quality control (QC) procedure and applying the phenotype-based inclusion/exclusion criteria, 1,322 mothers were selected and used in the analysis. The study was approved by the Ethics Committee of the Helsinki University Central Hospital. Written informed consent was given by all participants.

**GPN**

The Genomic and Proteomic Network for Preterm Birth Research (GPN) study [35]is a multicenter observational genome-wide association study (GWAS) designed to determine the genetic predisposition to idiopathic preterm birth. Phenotype data and genotype data from 743 spontaneous preterm births (20 to less than 34 weeks gestation), and 752 controls (39 to less than 42 weeks gestation) of diverse ethnic background (White, Hispanics, African Americans, and Others) were collected. For this current study, we identified 419 mothers of European descent using PCA from the data downloaded from dbGaP (phs000714.v1.p1).

**HAPO**

All pregnant women at less than 32 weeks of gestation were eligible for enrollment in HAPO unless they met one of several exclusion criteria. All participants gave written informed consent, and an external data monitoring committee provided oversight. Study phenotype collection methods and inclusion and exclusion criteria have been published elsewhere [36,37]. Participants underwent a 75-g oral glucose tolerance test (OGTT) at ~28 weeks' gestation. Maternal DNA was taken from blood collected into an EDTA tube at 2 h during the OGTT, when phenotypes of interest were measured, including glucose, blood pressure, weight, and height. Glucose and C-peptide were measured in a central laboratory [36,37], and DNA was prepared using the automated Autopure LS from Gentra Systems. Submitted for genotyping were 3,152 European ancestry mother and offspring HAPO samples, of which 2,797 survived quality control (QC). DNA samples were genotyped using the Illumina Human 610 Quad v1 B SNP array at the Broad Institute following agreed-upon protocols of the Gene-Environment Association Studies (GENEVA) consortium [38]. Genotype data that passed initial QC at the genotyping centers were released to the GENEVA Coordinating Center (CC), National Center for Biotechnology Information database of Genotypes and Phenotypes (dbGaP), and HAPO study teams, who collectively performed QC using procedures previously described by the GENEVA consortium [38]. Poorly performing samples or SNPs were removed based on misspecified sex, chromosomal anomalies, unintended sample duplicates, sample relatedness, low call rate, high number of Mendelian errors, departures from Hardy-Weinberg equilibrium, duplicate discordance, sex differences in heterozygosity, and low minor allele frequencies as described elsewhere [38,39].

For the analysis of parental transmitted and non-transmitted alleles, we utilized phenotype and genotype data downloaded from dbGaP (phs000096.v2.p1).

**HUNT**

From 2012-2015 the HUNT-Michigan (HUNT-MI) collaboration genotyped approximately 72.000 individuals from the HUNT biobank [40]. The genotyping effort was a research collaboration between researchers at NTNU and the University of Michigan. Every individual with a DNA sample with a suitable DNA concentration was selected for genotyping. Samples were picked at random and genotyped in batches. All genotyping was performed at the Genomics-Core Facility (GCF) at the Norwegian University of Science and Technology, NTNU. The Trøndelag Health Study (HUNT) is a large population-based cohort from the county Trøndelag in Norway. All residents in the county, aged 20 years and older, have been invited to participate. Data was collected through three cross-sectional surveys, HUNT1 (1984-1986), HUNT2 (1995-1997) and HUNT3 (2006- 2008), and has been described in detail previously [41], with the fourth survey recently completed (HUNT4, 2017-2019). DNA from whole blood was collected from HUNT2 and HUNT3, with genotypes available from 71,860 participants. All genotyped participants have signed a written informed consent regarding the use of data from questionnaires, biological samples and linkage to other registries for research purposes.

In total, DNA from 71,860 HUNT samples was genotyped using one of three different Illumina HumanCoreExome arrays (HumanCoreExome12 v1.0, HumanCoreExome12 v1.1 and UM HUNT Biobank v1.0). Samples that failed to reach a 99% call rate, had contamination > 2.5% as estimated with BAF Regress [42], large chromosomal copy number variants, lower call rate of a technical duplicate pair and twins, gonosomal constellations other than XX and XY, or whose inferred sex contradicted the reported gender, were excluded. Samples that passed quality control were analysed in a second round of genotype calling following the Genome Studio quality control protocol described elsewhere [43]. Genomic position, strand orientation and the reference allele of genotyped variants were determined by aligning their probe sequences against the human genome (Genome Reference Consortium Human genome build 37 and revised Cambridge Reference Sequence of the human mitochondrial DNA;http://genome.ucsc.edu) using BLAT [44]. Variants were excluded if (1) their probe sequences could not be perfectly mapped to the reference genome, cluster separation was < 0.3, Gentrain score was < 0.15, showed deviations from Hardy Weinberg equilibrium in unrelated samples of European ancestry with p-value < 0.0001), their call rate was < 99%, or another assay with higher call rate genotyped the same variant.

Ancestry of all samples was inferred by projecting all genotyped samples into the space of the principal components of the Human Genome Diversity Project (HGDP)

reference panel (938 unrelated individuals; downloaded from http://csg.sph.umich.edu/chaolong/LASER/) [45,46], using PLINK v1.90 [47]. Recent European ancestry was defined as samples that fell into an ellipsoid spanning exclusively European populations of the HGDP panel. The different arrays were harmonized by reducing to a set of overlapping variants and excluding variants that showed frequency differences > 15% between data sets, or that were monomorphic in one and had MAF > 1% in another data set. The resulting genotype data were phased using Eagle2 v2.3 [26] . If you need information about ancestry or population structure (principal components) please contact the K.G. Jebsen center for genetic epidemiology.
Imputation was performed on the 69,716 samples of recent European ancestry using Minimac3 (v2.0.1,http://genome.sph.umich.edu/wiki/Minimac3) [28] with default settings (2.5 Mb reference-based chunking with 500kb windows) and a customized Haplotype Reference consortium release 1.1 (HRC v1.1) for autosomal variants and HRC v1.1 for chromosome X variants [27] . The customized reference panel represented the merged panel of two reciprocally imputed reference panels: (1) 2,201 low-coverage whole-genome sequence samples from the HUNT study and (2) HRC v1.1 with 1,023 HUNT WGS samples removed before merging. We excluded imputed variants with Rsq < 0.3 resulting in over 24.9 million well-imputed variants. The study was approved by The Regional Committee for Medical Research Ethics (#2016/551).

**Northern Finland Birth Cohort 1966**

The Northern Finland Birth Cohort 1966 is a prospective follow-up study of children from the two northernmost provinces of Finland born in 1966 [48]. All individuals still living in northern Finland or the Helsinki area (n = 8,463) were contacted and invited for clinical examination. A total of 6007 participants attended the clinical examination at the participants' age of 31 years. DNA was extracted from blood samples given at the clinical examination (5,753 samples available) [49]. The subset with DNA is representative of the original cohort in terms of major environmental and social factors. Informed consent was obtained from all subjects. After performing standard sample QC we included 5,402 NFBC66 participants that were genotyped on an Illumina HumanCNV370DUO Analysis BeadChip. 329,401 variants were included in the imputation scaffold. Variants were imputed to the HRC reference r1.1 2016 [27] on the Michigan Imputation Server. Haplotypes were pre phased using ShapeIT v2 r790 [50] and imputation was performed using MACH. Prior to analysis we excluded variants monomorphic in this dataset. We used rvtests [51] for association testing. To correct for distant genetic relatedness and population stratification within NFBC1966 we calculated a genetic relationship matrix based on all SNPs with MAF > 5%.

NFBC data is available from the University of Oulu, Infrastructure for Population Studies. Permission to use the data can be applied for research purposes via electronic material request portal. In the use of data, we follow the EU general data protection regulation

(679/2016) and Finnish Data Protection Act. The use of personal data is based on cohort participant's written informed consent at his/her latest follow-up study, which may cause limitations to its use. Please, contact NFBC project center and visit the cohort website (www.oulu.fi/nfbc) for more information.

**Project Viva**

Study population: Project Viva is a longitudinal pre-birth cohort established to examine the effects of events during early development on lifetime health outcomes [52]. Between 1999 and 2002, the study recruited women in early pregnancy from eight obstetric offices of Atrius Harvard Vanguard Medical Associates, a multispecialty group practice in eastern Massachusetts, US. Exclusion criteria included multiple gestation, inability to answer questions in English, gestational age ≥22 weeks at recruitment, and plans to move away from the study area before delivery. Of 2312 eligible women, 2100 were still enrolled at delivery and had a live birth. For this analysis, we restricted to 604 white participants with genetic data and excluded 19 with pre-eclampsia, 107 who received any fertility treatment, 14 with low blood pressure, 19 with gestational diabetes, 6 who smoked ≥10 cigarettes/day and 8 who drank >1 servings/day of alcohol, leaving 431 participants in the analysis sample.

Women reported the date of last menstrual period (LMP) at study enrollment (median 9.9 weeks gestation). We obtained the date of delivery from the hospital medical record. We calculated the length of gestation in days by subtracting the date of the LMP from the date of delivery. If gestational age according to the second trimester ultrasound differed from that according to the LMP by more than 10 days, we used the ultrasound result to determine gestational duration.

We did not include principal components as covariates because none of the first 10 principal components were associated with gestational age.

Genotyping, phasing, and imputation: Samples were genotyped using the Illumina Infinium Core Exome-24 array. For quality control, we removed duplicates and low-quality samples, samples with SNP call rate <95% and samples with sex mismatch. We filtered out SNPs with call rate <98% (~5K), monomorphic SNPs (~800K) and SNPs with HWE p <1e-8 (~10K). Phasing and imputation was completed on the Michigan Imputation Server using ShapeIT v2.r790 and reference panel 1000G Phase 3 v5.

All mothers provided written informed consent. Institutional review boards at all participating institutions gave approval for this study.

Funding: Project Viva was supported by grants from the US National Institutes of Health (UH3 OD23286 and R01 HD034568).

**STORK**

The STORK study is a prospective cohort of 1031 healthy pregnant women of Scandinavian heritage who registered for obstetric care at the Oslo University Hospital Rikshospitalet from 2001 to 2008 [53]. Exclusion criteria were multiple pregnancies, known history of type 1 or type 2 diabetes mellitus, and severe chronic diseases (pulmonary, cardiac, gastrointestinal or renal).

DNA samples were genotyped on the Illumina Infinium CoreExome chip using Illumina iScan by the Department of Clinical Sciences, Clinical Research Centre, Lund University, Malmö, Sweden. Of the 1031 individuals who provided blood samples, 529 participants had usable SNP data after genotyping and quality control described elsewhere [54]. All 529 women had data on gestational length.

The study was approved by the Regional Committee for Medical Research Ethics, Southern Norway, Oslo, Norway (reference number S-2014/224-0119a and S-07392a) and performed according to the Declaration of Helsinki. All participating women provided written informed consent.

**STORK Groruddalen**

The STORK Groruddalen study (STORK-G) is a population-based cohort which included 823 healthy women attending three public mother–child health clinics for antenatal care in the multi-ethnic area of Groruddalen, Oslo, Norway [55]. Women were eligible if they: (i) lived in the study districts; (ii) planned to give birth at one of two study hospitals; (iii) were <20 weeks of pregnant; (iv) could communicate in Norwegian or any of the eight translated languages; (v) were able to give an informed consent. Women with pregestational diabetes or in need of intensive hospital follow-up during pregnancy were excluded. The participation rate was 74%, varying from 63.9% to 82.6% across ethnic groups.

DNA samples were genotyped on the Illumina Infinium CoreExome chip using Illumina iScan by the Department of Clinical Sciences, Clinical Research Centre, Lund University, Malmö, Sweden. Of the 664 genotyped samples those with low call rate (i.e. < 95%, n=0), extreme heterozygosity (> |mean± (3xSD)|, n=1), mismatched gender (n=24) or cryptic relatedness (i.e. one individual (chosen at random) from each related pair, defined as genome-wide IBD > 0.185 (n=6) were excluded from analyses. Genetic ethnic origin was defined by ancestry informative principal component analysis based on the variance-standardized relationship matrix generated in PLINK 1.9 software package [47] were used. In STORK Groruddalen a total of 310 women with European ancestry are present after quality control has been performed. 310 women of European ancestry had data on gestational length.

The study was approved by the Regional Committee for Medical Research Ethics, Southern Norway, Oslo, Norway (ref.number 2015/1035) and performed according to the Declaration of Helsinki. We obtained written informed consent from all participants before any study-related procedure.

**The Genetics of Glucose regulation in Gestation and Growth (Gen3G)**
Gen3G is a prospective population-based pre-birth cohort that recruited pregnant women receiving prenatal care at the Centre Hospitalier Universitaire de Sherbrooke (CHUS) in QC, Canada between January 2010 to June 2013 [56]. Participating women were enrolled in the first trimester of pregnancy. Exclusion criteria included history of overt diabetes or laboratory evidence of overt diabetes at the first trimester study visit (hemoglobin A1C ≥6.5% or glucose ≥185 mg/dl after a 50-gram glucose load), multiple pregnancy, and use of medications that affect glucose metabolism. Of 1024 eligible women, 873 were still enrolled at delivery and had a live birth. For this analysis, we restricted to 582 participants with genetic data and excluded 37 with pre-eclampsia or GDM or hypertension, 46 with a repeat c-section, 6 with breech presentation, 19 who smoked ≥10 cigarettes/day and 15 who were not of european ethnicity, leaving 459 participants in the analysis sample.

Women reported the date of last menstrual period (LMP) at study enrollment (median 9.6 weeks gestation). We obtained the date of delivery from the hospital medical record. We calculated the length of gestation in days by subtracting the date of the LMP from the date of delivery. If gestational age according to the second trimester ultrasound differed from that according to the LMP by more than 5 days, we used the ultrasound result to determine gestational duration.

Three principle components were included as covariates because they were associated with gestational age ($p<0.1$).

We isolated DNA from maternal blood buffy coats using the Gentra Puregene Blood Kit (Qiagen, Mississauga, ON, Canada). 598 women samples were genotyped using Illumina MEGAex arrays. For quality control, we removed duplicates and samples with SNP call rate <98%. We also filtered SNPs with call rate <95% (~141K), monomorphic SNPs and SNPs with HWE p <1e-8 (~757K) and SNPs with concordance between duplicates <90% (37). Before running the imputation, we also removed insertions/deletions (738), SNPs on chromosome XY and 0 (~9K), SNPs with minor allele frequency <0.01 (~269K) and duplicate SNPs with the lower call rate (~19K), After QC, we had data available on 582 women over 838,884 SNPs. We performed phasing and imputation on the Michigan Imputation Server using ShapeIT v2.r790 and reference panel 1000G Phase 3 v5.

Ethical approval was obtained from the CHUS ethic committee board and every participant gave written informed consent before enrolment in the study.

Funding: The Gen3G prospective cohort was supported by the Fonds de Recherche du Québec – Santé (FRQS – subvention Fonctionnement – Recherche Clinique - grant #20697) and by a Canadian Institute of Health Research (CIHR) Operating grant (Institute of Nutrition, Metabolism and Diabetes; MOP- 115071).

**The Norwegian Mother, Father and Child Cohort Study**

Study population

The Norwegian Mother, Father and Child Cohort Study (MoBa) is an open-ended cohort study that recruited pregnant women in Norway from 1999 to 2008. Approximately 114,500 children, 95,200 mothers, and 75,000 fathers of predominantly Norwegian ancestry were enrolled in the study from 50 hospitals all across Norway [57,58]. The project Better Health by Harvesting Biobanks (HARVEST) randomly selected 11,490 umbilical cord blood DNA samples from the biobank of this study for family triad genotyping, excluding samples matching any of the following criteria: (1) stillborn, (2) deceased, (3) twins, (4) non-existing data at the Norwegian Medical Birth Registry, (5) missing anthropometric measurements at birth in Medical Birth Registry, (6) pregnancies where the mother did not answer the first questionnaire (as a proxy for higher dropout rate), and (7) missing parental DNA samples. In 2016, HARVEST randomly selected a second set of 8,900 triads using the same criteria. The same year NORMENT selected 5,910 triads with the same selection criteria as HARVEST, and extended this with 3,209 triads in 2018.

Genotyping, phasing and imputation

Genotyping of the samples was performed in seven different batches on different Illumina platforms (HumanCoreExome-12 v.1.1 and HumanCoreExome-24 v.1.0, Illumina's Global Screening Array v.1.0, InfiniumOmniExpress-24v1.2 and HumanOmniExpress-24-v1.0). The Genome Reference Consortium Human Build 37 (GRCh37) reference genome was used for all annotations.

Genotypes were called in Illumina GenomeStudio (v.2011.1 and v.2.0.3). Cluster positions were identified from samples with call rate ≥ 0.98 and GenCall score ≥ 0.15. We excluded variants with low call rates, signal intensity, quality scores, and deviation from Hardy-Weinberg equilibrium (HWE) based on the following QC parameters: call rate < 98 %, cluster separation < 0.4, 10% GC-score < 0.3, AA T Dev > 0.025, HWE *p*-value $< 1 \times 10^{-6}$. Samples were excluded based on call rate < 98 % and heterozygosity excess > 4 SD. Study participants with recent white Nordic ancestry were included after merging with ancestry reference samples from the HapMap project (ver. 3).

Pre-phasing was conducted locally using Shapeit v2.790[50]. Imputation was performed at the Sanger Imputation Server with positional Burrows-Wheeler transform and HRC version 1.1 as reference panel [27].

Phenotype

The MoBa cohort is linked to the Medical Birth Registry of Norway, from where we extracted information on gestational duration, and medically initiated delivery (inductions or c-sections).Maternal health information prior to and during pregnancy, including gestational duration and parity, as well as complications of pregnancy and birth were recovered from the Medical Birth Registry of Norway. We defined spontaneous onset of delivery as a delivery initiated by spontaneous contractions or rupture of membranes. Deliveries initiated by induction methods, including the use of prostaglandins, oxytocin, amniotomy or any other induction procedure, or planned cesarean section were excluded for gestational duration, and whenever these occurred in preterm or post-term cases.

Association analysis

Prior to analysis, we used a greedy algorithm to exclude one individual from each set of related subjects, where sets were defined as two individuals with a kinship coefficient >0.125. All analyses were performed using BOLT-LMM [12] (gestational duration) or REGENIE [59] (preterm and post-term deliveries), using genotype dosage as input and including parity and the 10 first principal components as covariates.

Informed consent was obtained from all study participants. The administrative board of the Norwegian Mother, Father and Child Cohort Study led by the Norwegian Institute of Public Health approved the study protocol. The MoBa cohort is currently regulated by the Norwegian Health Registry Act. The study was approved by The Regional Ethics Committee for Medical Research Ethics South East Norway (#2015/2425).

**The Preterm birth Genome Project**

The Preterm birth Genome Project (PGP) is a consortium initiated by the Preterm Birth International Collaborative (PREBIC), the World Health Organisation (WHO), and the March of Dimes [60]. Participants in the PGPII and PGPIII cohorts were recruited between the years 2007 and 2010, and between years 2013 and 2016, respectively. For both cohorts, DNA samples and detailed phenotypic data were collected (based on the

Optimal PREBIC dataset [61]) from women who delivered in Perth, Western Australia. Informed written consent was obtained from the study participants. Genetic samples were collected from peripheral blood or saliva on days one to three post-delivery. DNA were extracted and genotyped separately for each cohort. Cleaned genotyped data were phased using Eagle2 [26] and imputed using Minimac3 and Minimac4 [28] against the Haplotype Reference Consortium (version r1.1 2016) reference panel [27] on the Michigan Imputation Server. Ethics approvals for PGPII and PGPIII were obtained from the Women's and Newborn Health Service (HREC approvals 1572EW and 2013030EW).

### Cohort acknowledgements


**23andMe**

We would like to thank the research participants and employees of 23andMe for making this work possible.

**ALSPAC**

Core funding for ALSPAC is provided by the UK Medical Research Council and Wellcome (217065/Z/19/Z) and the University of Bristol. Genotyping of the ALSPAC maternal samples was funded by Wellcome (WT088806) and the offspring samples were genotyped by Sample Logistics and Genotyping Facilities at the Wellcome Sanger Institute and LabCorp (Laboratory Corporation of America) using support from 23andMe. A comprehensive list of grants funding is available on the ALSPAC website (http://www.bristol.ac.uk/alspac/external/documents/grant-acknowledgements.pdf). We are extremely grateful to all the families who took part in ALSPAC, the midwives for their help in recruiting them, and the whole ALSPAC team, which includes interviewers, computer and laboratory technicians, clerical workers, research scientists, volunteers, managers, receptionists and nurses.

**BiB**

Born in Bradford (BiB) data used in this research was funded by Wellcome (WT101597MA), a joint grant from the UK Medical Research Council (MRC) and UK Economic and Social Science Research Council (ESRC) (MR/N024397/1), the British Heart Foundation (CS/16/4/32482) and the National Institute for Health Research (NIHR) under its Collaboration for Applied Health Research and Care (CLAHRC) for Yorkshire and Humber and the Clinical Research Network (CRN). Born in Bradford is only possible because of the enthusiasm and commitment of the Children and Parents in BiB. We are grateful to all the participants, teachers, school staff, health professionals and researchers who have made Born in Bradford happen.

**CHOP**
The authors thank the network of primary care clinicians and the patients and families for their contribution to this project and to clinical research facilitated by the Pediatric Research Consortium (PeRC) at The Children's Hospital of Philadelphia. R. Chiavacci, E. Dabaghyan, A. (Hope) Thomas, K. Harden, A. Hill, C. Johnson-Honesty, C. Drummond, S. Harrison, F. Salley, C. Gibbons, K. Lilliston, C. Kim, E. Frackelton, G. Otieno, K. Thomas, C. Hou, K. Thomas and M.L. Garris provided expert assistance with genotyping and/or data collection and management. The authors would also like to thank S. Kristinsson, L.A. Hermannsson and A. Krisbjšrnsson of Rafšrninn ehf for extensive software design and contributions.

**DBDS**
We thank deCODE genetics/Amgen, Bio- and Genomebank Denmark and Department of Clinical Immunology, Copenhagen University Hospital for financial support of this study.

**DNBC**
The Danish National Birth Cohort (DNBC) was established with a significant grant from the Danish National Research Foundation. Additional support was obtained from the Danish Regional Committees, the Pharmacy Foundation, the Egmont Foundation, the March of Dimes Birth Defects Foundation, the Health Foundation and other minor grants. The DNBC biobank is a part of the Danish National Biobank resource, which was established with the support of major grants from the Novo Nordisk Foundation, the Danish Medical Research Council and the Lundbeck Foundation.

The DNBC preterm delivery study (DNBC-PTD) is a nested study within the DNBC. The generation of GWAS genotype data for the DNBC-PTD sample was carried out within the Gene Environment Association Studies (GENEVA) consortium with funding provided through the National Institutes of Health's Genes, Environment, and Health Initiative (U01HG004423; U01HG004446; U01HG004438).

The Genomics of overweight in Young Adults (GOYA) study is a nested study within the DNBC, and conducted in collaboration with the MRC Integrative Epidemiology Unit at the University of Bristol (MC_UU_12013/1-9). The genotyping for DNBC-GOYA study was funded by the Wellcome Trust (WT 084762).

We thank dbGAP for depositing and hosting the phenotype and genotype data of the DNBC data set.

**EFSOCH**

The Exeter Family Study of Childhood Health (EFSOCH) was supported by South West NHS Research and Development, Exeter NHS Research and Development, the Darlington Trust and the Peninsula National Institute of Health Research (NIHR) Clinical Research Facility at the University of Exeter. The opinions given in this paper do not necessarily represent those of NIHR, the NHS or the Department of Health. Genotyping of the EFSOCH study samples was funded by the Wellcome Trust and Royal Society grant 104150/Z/14/Z.

**EGCUT**

This study was funded by EU H2020 grant 692145, Estonian Research Council Grant IUT20-60, IUT24-6, and European Union through the European Regional Development Fund Project No. 2014-2020.4.01.15-0012 GENTRANSMED and 2014-2020.4.01.16-0125. Data analyzes were carried out in part in the High-Performance Computing Center of University of Tartu.

**FIN**

We thank the participants in the Finnish birth cohort as well as the research group who collected the data. We thank the infants and their parents who agreed to take part in the GPN study and the medical and nursing colleagues who collected that data. NIH (1R01HD101669-01A1), Burroughs Wellcome Fund (10172896), the March of Dimes Prematurity Research Center Ohio Collaborative, a grant from the Cincinnati Children's Hospital Medical Center (GAP/RIP).

**FINNGEN**

We want to acknowledge the participants and investigators of the FinnGen study.

**Gen3G**

Gen3G investigators would like to acknowledge all Gen3G participants, the research staff from the endocrinology group at the Centre de Recherche of the Centre Hospitalier Universitaire de Sherbrooke (CRCHUS), the clinical and research staff of the Clinique de Prélèvement en Grossesse du CHUS, and the clinical staff from the CHUS Obstetric Department. The CRCHUS is a FRQ-Sante affiliated research centre; recruitment and follow-up of Gen3G participants was possible based on the on-going support from the CRCHUS.

**GPN**

The GPN datasets used for the analyses described in this manuscript were obtained from dbGaP at https://www.ncbi.nlm.nih.gov/gap/ through dbGaP accession number phs000714.v1.p1. We thank dbGAP for depositing and hosting the phenotype and genotype data of the GPN data set.

**HAPO**

This study was supported by National Institutes of Health (NIH) grants HD-34242, HD-34243, HG-004415, and CA-141688, and by the American Diabetes Association. The authors are indebted to the participants of the HAPO Study at the following centers: Newcastle and Brisbane, Australia; Toronto, Canada; and Belfast, U.K. We thank dbGAP for depositing and hosting the phenotype and genotype data of the HAPO data set.

**HUNT**

The Trøndelag Health Study (HUNT) is a collaboration between HUNT Research Centre (Faculty of Medicine and Health Sciences, Norwegian University of Science and Technology NTNU), Trøndelag County Council, Central Norway Regional Health Authority, and the Norwegian Institute of Public Health. The genetic investigations of the HUNT Study, is a collaboration between researchers from the K.G. Jebsen center for genetic epidemiology and University of Michigan Medical School and the University of Michigan School of Public Health. The K.G. Jebsen Center for Genetic Epidemiology is financed by Stiftelsen Kristian Gerhard Jebsen; Faculty of Medicine and Health Sciences, NTNU, Norway.

**MoBa**

This work was supported by grants (B.J.) from The Swedish Research Council, Stockholm, Sweden (2015-02559), The Research Council of Norway, Oslo, Norway (FRIMEDBIO #547711, #273291), March of Dimes (#21-FY16-121) and (to P.R.N.) from the European Research Council (AdG SELECTionPREDISPOSED #293574), the Bergen Research Foundation ("Utilizing the Mother and Child Cohort and the Medical Birth Registry for Better Health"), Stiftelsen Kristian Gerhard Jebsen (Translational Medical Center), the University of Bergen, the Research Council of Norway (FRIPRO grant #240413), the Western Norway Regional Health Authority (Strategic Fund "Personalized Medicine for Children and Adults"), the Novo Nordisk Foundation (grant #54741), and the Norwegian Diabetes Association. This work was partly supported by the Research Council of Norway through its Centres of Excellence funding scheme (#262700, #223273), Better Health by Harvesting Biobanks (#229624) and The Norwegian Mother, Father and Child Cohort Study is supported by the Norwegian Ministry of Health and Care Services and the Ministry of Education and Research, NIH/NIEHS (contract no N01-ES-75558), NIH/NINDS (grant no.1 UO1 NS 047537-01 and grant no.2 UO1 NS 047537-06A1). We are grateful to all the families in Norway who are taking part in this ongoing cohort study. All analyses were performed using digital labs in HUNT Cloud at the Norwegian University of Science and Technology, Trondheim, Norway.

The Norwegian Mother, Father and Child Cohort Study is supported by the Norwegian Ministry of Health and Care Services and the Ministry of Education and Research. We are grateful to all the participating families in Norway who take part in this on-going cohort study. We thank the Norwegian Institute of Public Health (NIPH) for generating high-quality genomic data. We thank the Norwegian Institute of Public Health (NIPH) for generating high-quality genomic data. This research is part of the HARVEST collaboration, supported by the Research Council of Norway (#229624). We also thank the NORMENT Centre for providing genotype data, funded by the Research Council of Norway (#223273), South East Norway Health Authority and KG Jebsen Stiftelsen. We further thank the Center for Diabetes Research, the University of Bergen for providing genotype data and performing quality control and imputation of the data funded by the ERC AdG project SELECTionPREDISPOSED, Stiftelsen Kristian Gerhard Jebsen, Trond Mohn Foundation, the Research Council of Norway, the Novo Nordisk Foundation, the University of Bergen, and the Western Norway health Authorities (Helse Vest). All analyses were performed using digital labs in HUNT Cloud at the Norwegian University of Science and Technology, Trondheim, Norway.

**NFBC**

NFBC1966 received financial support from the Academy of Finland (project grants 104781, 120315, 129269, 1114194, 24300796, Center of Excellence in Complex Disease Genetics and SALVE), University Hospital Oulu, Biocenter, University of Oulu, Finland (75617), NHLBI grant 5R01HL087679-02 through the STAMPEDE program (1RL1MH083268-01), NIH/NIMH (5R01MH63706:02), ENGAGE project and grant agreement HEALTH-F4-2007-201413, EU FP7 EurHEALTHAgeing -277849, the Medical Research Council, UK (G0500539, G0600705, G1002319, PrevMetSyn/SALVE) and the MRC, Centenary Early Career Award. The program is currently being funded by the H2020-633595 DynaHEALTH action, academy of Finland EGEA-project (285547) and EU H2020 ALEC project (Grant Agreement 633212).

**PGP**

This study was funded by the World Health Organisation, March of Dimes, National Health & Medical Research Council (APP1042267).

**Project Viva**

We thank the participants and staff of Project Viva.

**STORK-G**

The authors thank study staff and general staff at the Child Health Clinics in Stovner, Grorud and Bjerke districts in Oslo and at the delivery- and post-natal wards at

Akershus University Hospital and Oslo University Hospital, for help with collecting the data, and the Hormone Laboratory, Oslo University Hospital for DNA-extraction.

**STORK**

The STORK study received additional funding from the Norwegian Diabetes Association, the Norwegian Odd Fellow Research Fund and Johan Selmer Kvanes' Endowment for Research in Diabetes.

**UK-Biobank**

We want to thank the participants of the UK Biobank and the investigators of the Pan-UK Biobank team.

**WTCCC58BC**

The management of the 1958 Birth Cohort is funded by the Economic and Social Research Council (grant number ES/M001660/1). Access to these resources was enabled via the 58READIE Project funded by Wellcome Trust and Medical Research Council (grant numbers WT095219MA and G1001799). DNA collection was funded by MRC grant G0000934 and cell-line creation by Wellcome Trust grant 068545/Z/02. This study makes use of data generated by the Wellcome Trust Case-Control Consortium. A full list of investigators who contributed to generation of the data is available from the Wellcome Trust Case-Control Consortium website. Funding for the project was provided by the Wellcome Trust under the award 076113. This research used resources provided by the Type 1 Diabetes Genetics Consortium, a collaborative clinical study sponsored by the National Institute of Diabetes and Digestive and Kidney Diseases (NIDDK), National Institute of Allergy and Infectious Diseases, National Human Genome Research Institute, National Institute of Child Health and Human Development, and Juvenile Diabetes Research Foundation International (JDRF) and supported by U01 DK062418.

### Author funding statements

B.J. received funding from The Swedish Research Council, Stockholm, Sweden (2015-02559 and 2019-01004), The Research Council of Norway, Oslo, Norway (FRIMEDBIO #547711, #273291), March of Dimes (#21-FY16-121). Research reported in this publication (B.J., G.Z. and R.M.F.) was supported by the Eunice Kennedy Shriver National Institute Of Child Health & Human Development of the National Institutes of Health under Award Number R01HD101669. The content is solely the responsibility of the authors and does not necessarily represent the official views of the National Institutes of Health. G.H.M has received funding from the Norwegian Diabetes

Association and Nils Normans minnegave. G.H.M. is supported by the Norwegian Research Council (Postdoctoral mobility research grant 287198). M.C.B's contribution to this work was supported by a UK Medical Research Council (MRC) Skills Development Fellowship (MR/P014054/1) and a University of Bristol Vice-Chancellor's fellowship. M.C.B and D.A.L are supported by the British Heart Foundation (AA/18/7/34219) and work in a Unit that receives funding from the University of Bristol and UK Medical Research Council (MRC) (MC_UU_00011/6). M.V. is supported by the Research Council of Norway (project #301178). D.A.L is supported by a British Heart Foundation Chair (CH/F/20/90003). S.F.A Grant is supported by Daniel B. Burke Chair for Diabetes Research and NIH Grant R01 HD056465. IDF to CAG center from CHOP; CHOP's Endowed Chair in Genomic Research. Funding for T.L was provided by the European Regional Development Fund and the programme Mobilitas Pluss (MOBTP155). B.S is a core member of the NIHR Exeter Clinical Research Facility. R.M.F and R.N.B were funded by a Wellcome Trust and Royal Society Sir Henry Dale Fellowship (WT104150). R.M.F. is funded by a Wellcome Trust Senior Research Fellowship (WT220390). B.F was supported by the Oak Foundation. L.B is a senior research scholar from the Fonds de la recherche du Québec en santé (FRQS) and member of the CR-CHUS, a FRQS-funded Research Center. E.O. has received funding from the US National Institutes of Health. D.W. is funded by the Novo Nordisk Foundation (NNF18SA0034956, NNF14CC0001, NNF17OC0027594).
