## Extended Data Figures for "Genetic effects on the timing of parturition and links to fetal birth weight"

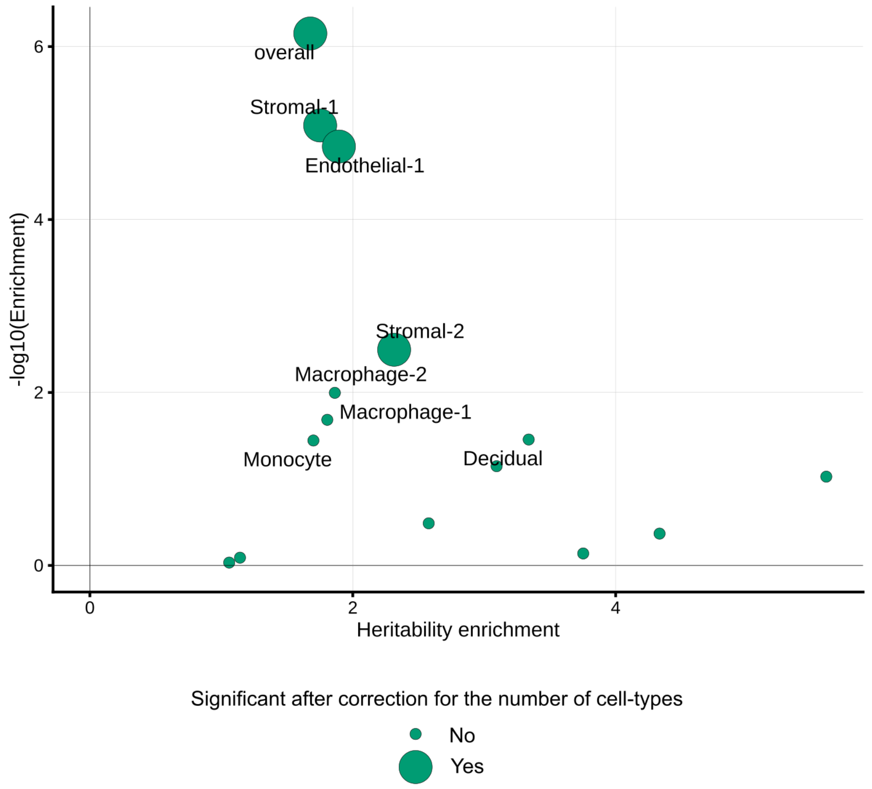


**Extended Data Fig. 1. SNP-heritability enrichment of gestational duration for genes differentially expressed during labor in different cell types of the myometrium and overall.** LD-score regression was used to partition heritability, and estimate the heritability enrichment for each cell type and overall. We calculated LD scores (European individuals from phase 3 of the 1000 Genomes project) for sets of genes differentially expressed at labor (± 100 kb) for each cell type separately and for the overall set of genes differentially expressed in the myometrium. Each dot represents a cell type, with larger showing significant heritability enrichment after correcting for the number of cell types. See Online Methods for a cautionary note regarding the comparison of different cell-type enrichment p-values.

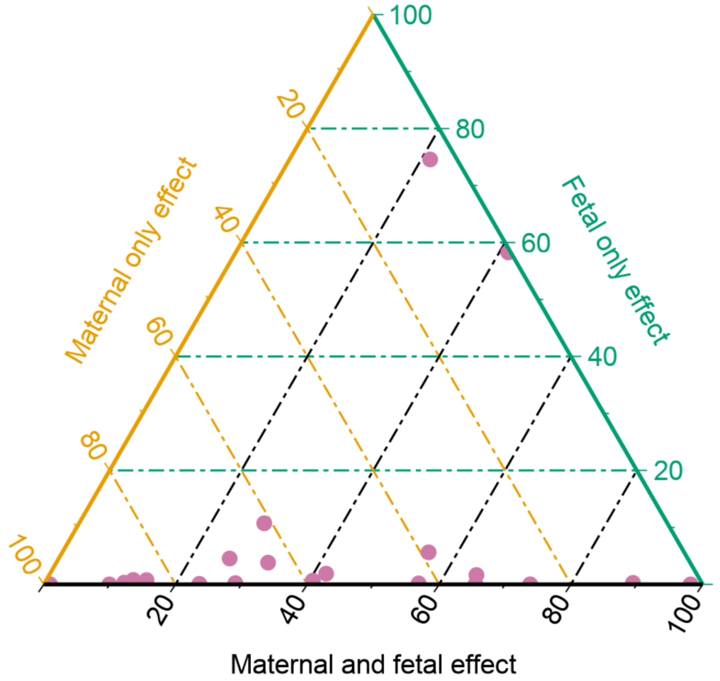


**Extended Data Fig. 2. Ternary plot representing the probabilities of having maternal, fetal or maternal and fetal effect for each index SNP.** The sum of all probabilities for each index SNP is 1. Lines are coloured according to the axis they belong to. All points in a horizontal line (green) have the same probability of "fetal only effect", points on a line (yellow) parallel to the right side of the triangle have the same probability of a "Maternal only effect", and lines (black) parallel to the left side of the triangle have the same probability of a "Maternal and fetal effect". Probabilities were obtained using Gaussian Mixture models clustering using the effect size and standard error estimates of the parental transmitted and non-transmitted alleles (n = 136,833 parent-offsprings). While five different clusters were identified, the fetal effect was broken down into two groups (parent-of-origin and independent of parent-of-origin) and the maternal and fetal effects also into two groups (same or opposite maternal and fetal direction). For this figure, probability of a "Fetal only effect" is the sum of the two groups with fetal effect, and "Maternal and fetal effect" is the sum of the probabilities of the two clusters with maternal and fetal effects.

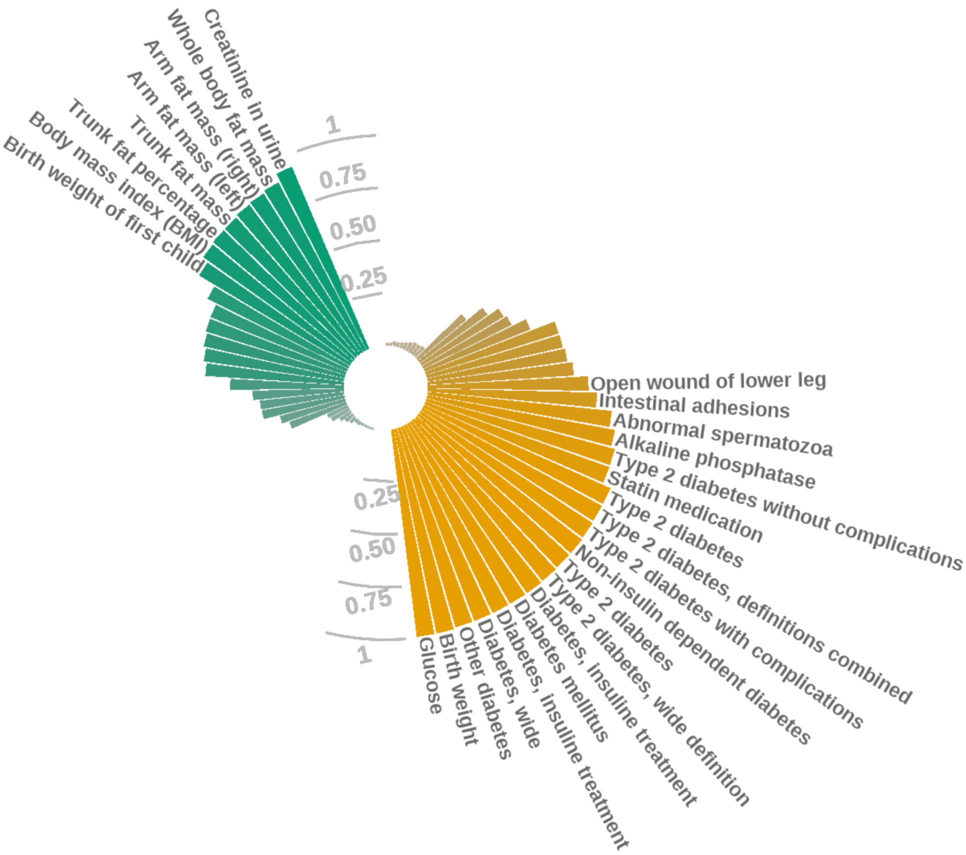


**Extended Data Fig. 3. Colocalization between the maternal effects on gestational duration (green) and fetal effects on birth weight (yellow) and other phenotypes from UK Biobank and FINNGEN at the *ADCY5* locus.** Posterior probability of colocalization between the maternal effect on gestational duration (rs28654158) and the fetal only effect on birth weight (rs11708067) with traits from UK Biobank and FINNGEN. Only traits with a posterior probability of colocalization >= 0.01 are plotted, and names are only shown if the posterior probability is > 0.5. Maternal locus on gestational duration was centered around rs28654158 (± 1.5 Mb) and the fetal locus on birth weight around rs11708067 (± 1.5 Mb).

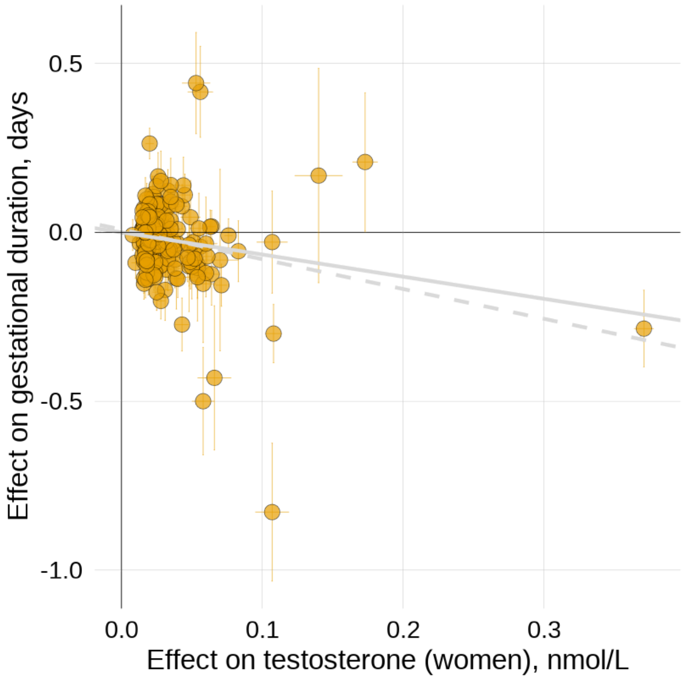


**Extended Data Fig. 4. Association between testosterone levels (women) and maternal effect on gestational duration.** Scatterplot for two-sample Mendelian randomization analysis for the effect of testosterone (independent of SHBG) on gestational duration (maternal effect). Each dot represents one of the testosterone associated SNPs. Horizontal and vertical error bars represent the 95% CI. The gray line depicts the inverse-variance weighted method estimate, and the gray-dashed line the MR-Egger estimate.
