## Supplementary Figures for "Genetic effects on the timing of parturition and links to fetal birth weight"

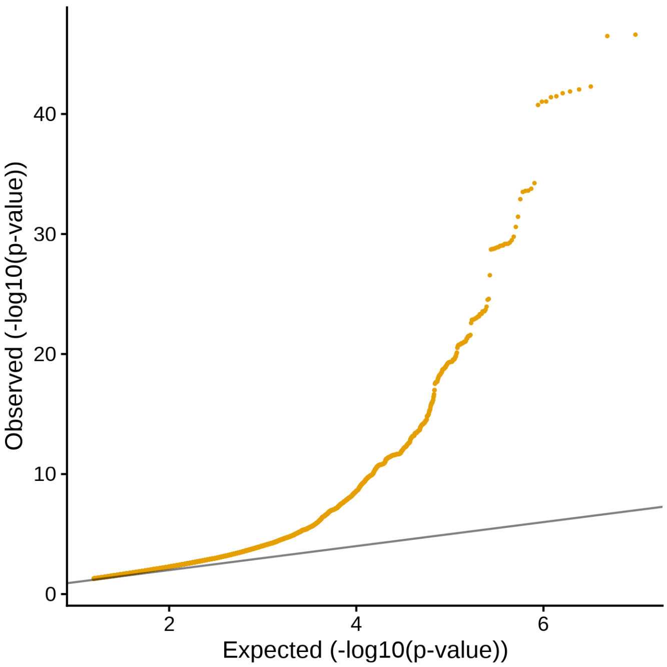


**Supplementary Fig. 1. Quantile-quantile plot of gestational duration GWAS meta-analysis.**

A

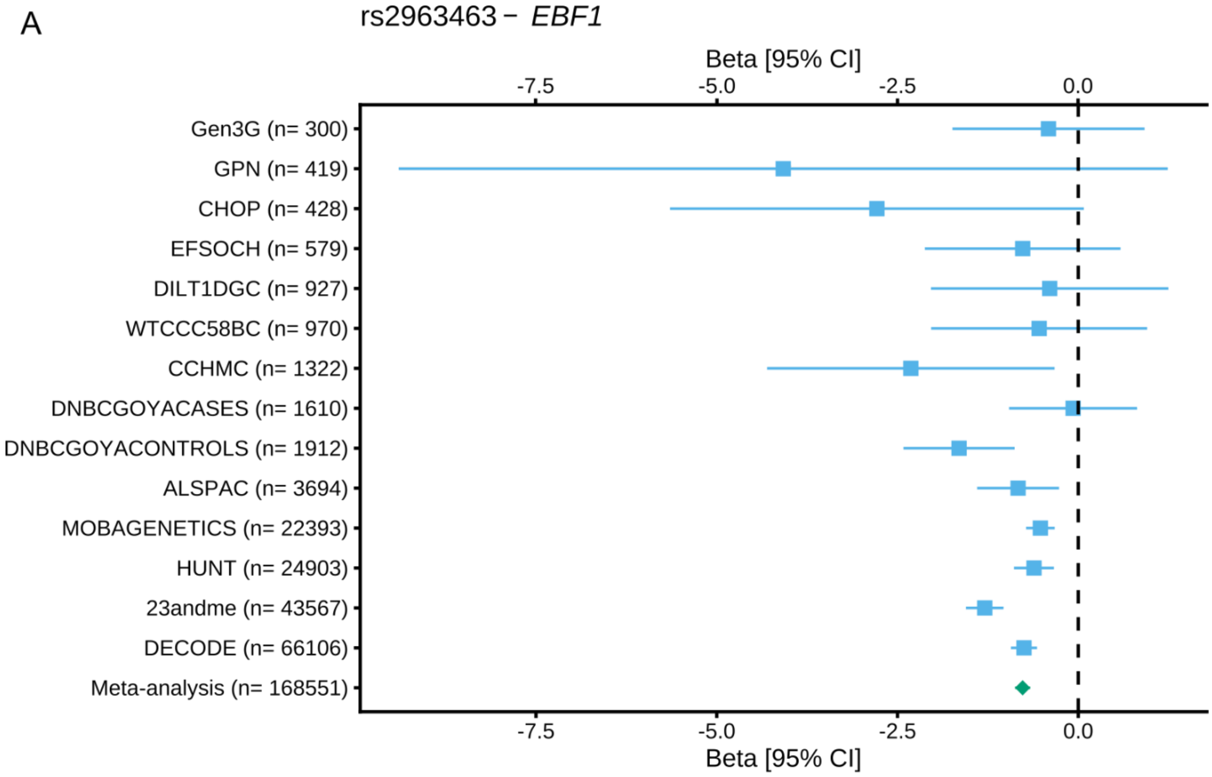
rs2963463 − EBF1
Beta [95% CI]
-7.5
-5.0
-2.5
0.0
Gen3G (n= 300)
GPN (n= 419)
CHOP (n= 428)
EFSOCH (n= 579)
DILT1DGC (n= 927)
WTCCC58BC (n= 970)
CCHMC (n= 1322)
DNBCGOYACASES (n= 1610)
DNBCGOYACONTROLS (n= 1912)
ALSPAC (n= 3694)
MOBAGENETICS (n= 22393)
HUNT (n= 24903)
23andme (n= 43567)
DECODE (n= 66106)
Meta-analysis (n= 168551)
-7.5
-5.0
-2.5
0.0
Beta [95% CI]

B

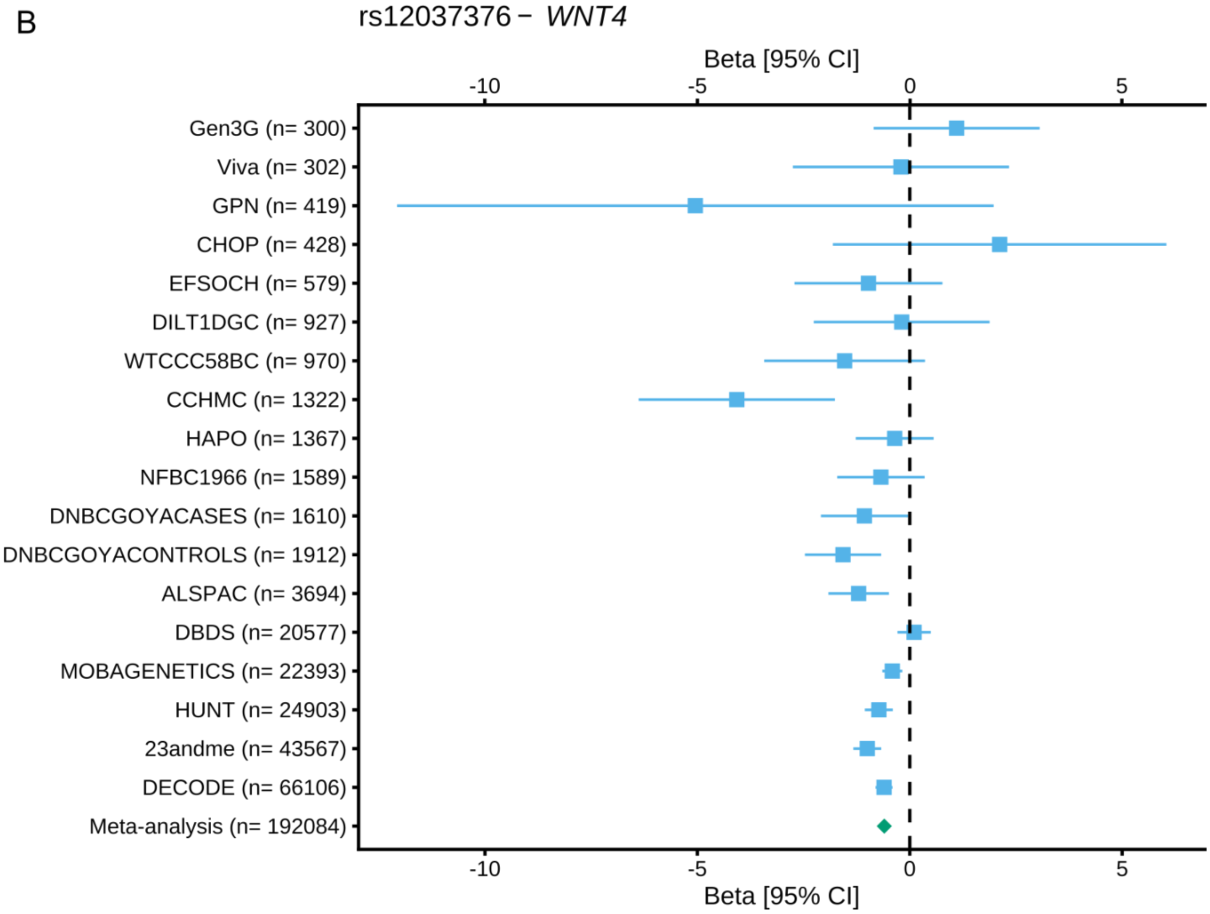
rs12037376 − WNT4
Beta [95% CI]
-10
-5
0
5
Gen3G (n= 300)
Viva (n= 302)
GPN (n= 419)
CHOP (n= 428)
EFSOCH (n= 579)
DILT1DGC (n= 927)
WTCCC58BC (n= 970)
CCHMC (n= 1322)
HAPO (n= 1367)
NFBC1966 (n= 1589)
DNBCGOYACASES (n= 1610)
DNBCGOYACONTROLS (n= 1912)
ALSPAC (n= 3694)
DBDS (n= 20577)
MOBAGENETICS (n= 22393)
HUNT (n= 24903)
23andme (n= 43567)
DECODE (n= 66106)
Meta-analysis (n= 192084)
-10
-5
0
5
Beta [95% CI]

C

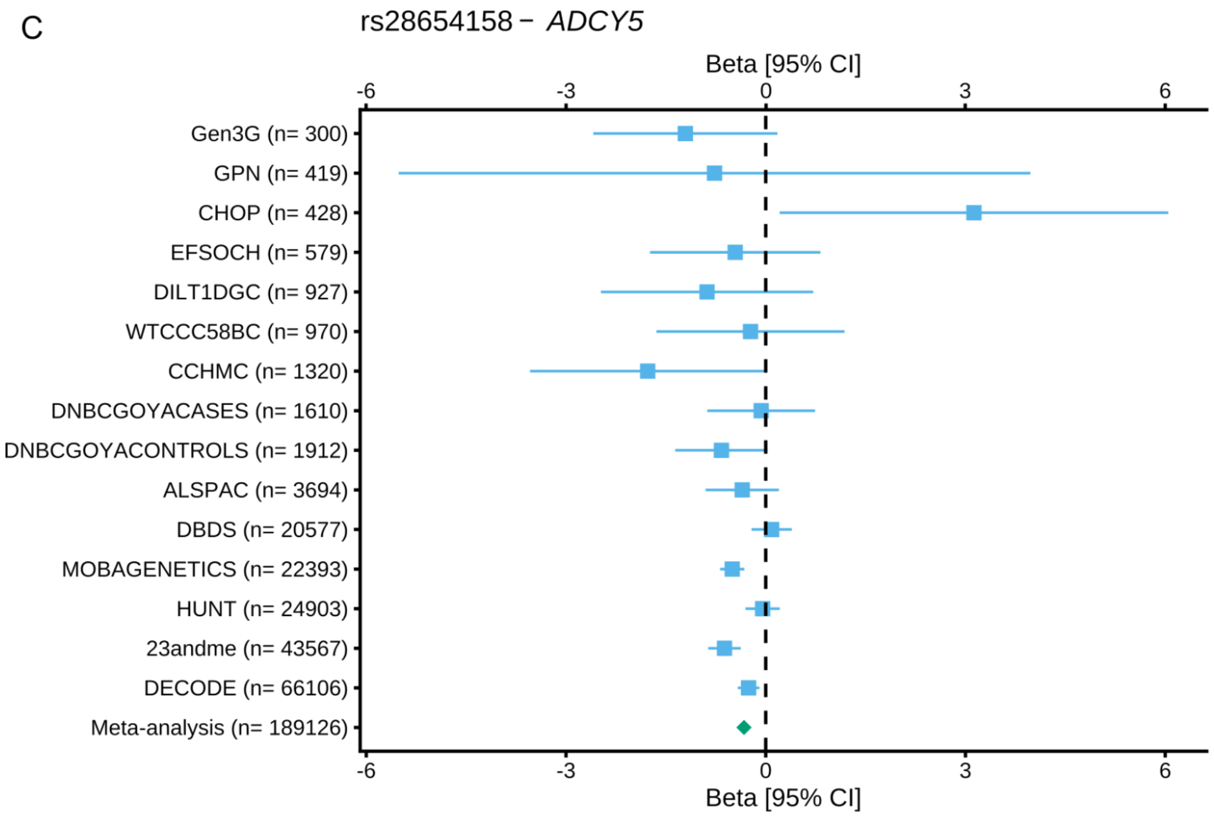
rs28654158 − ADCY5
Beta [95% CI]
-6
-3
0
3
6
Gen3G (n= 300)
GPN (n= 419)
CHOP (n= 428)
EFSOCH (n= 579)
DILT1DGC (n= 927)
WTCCC58BC (n= 970)
CCHMC (n= 1320)
DNBCGOYACASES (n= 1610)
DNBCGOYACONTROLS (n= 1912)
ALSPAC (n= 3694)
DBDS (n= 20577)
MOBAGENETICS (n= 22393)
HUNT (n= 24903)
23andme (n= 43567)
DECODE (n= 66106)
Meta-analysis (n= 189126)

D

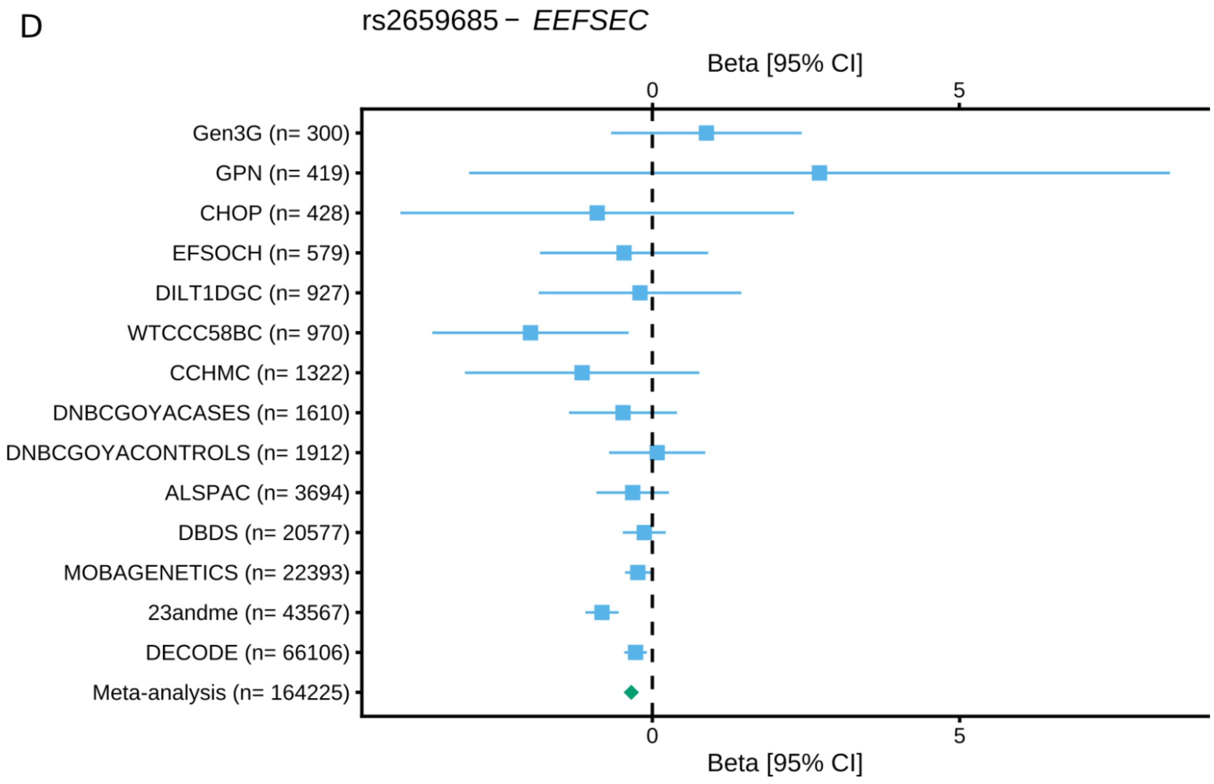
rs2659685 − EEFSEC
Beta [95% CI]
0
5
Gen3G (n= 300)
GPN (n= 419)
CHOP (n= 428)
EFSOCH (n= 579)
DILT1DGC (n= 927)
WTCCC58BC (n= 970)
CCHMC (n= 1322)
DNBCGOYACASES (n= 1610)
DNBCGOYACONTROLS (n= 1912)
ALSPAC (n= 3694)
DBDS (n= 20577)
MOBAGENETICS (n= 22393)
23andme (n= 43567)
DECODE (n= 66106)
Meta-analysis (n= 164225)

E

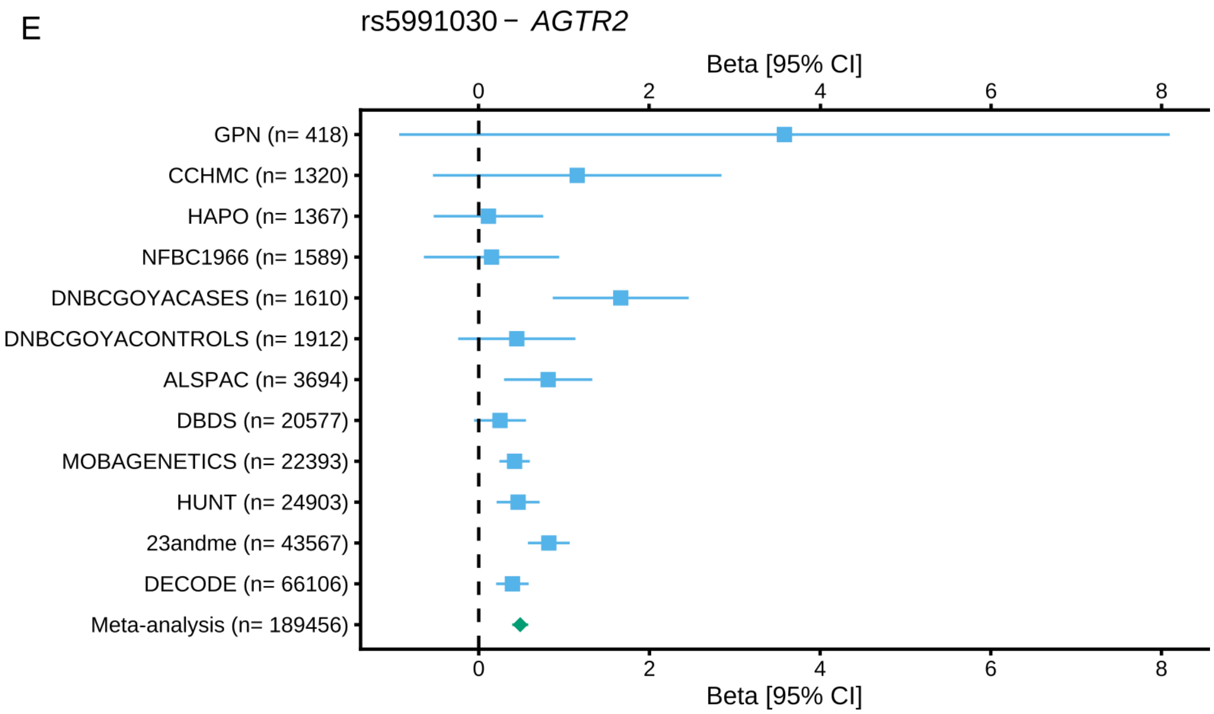


**Supplementary Fig. 2. Forest plots of the five top variants associated with gestational duration that had significant heterogeneity after fixed effects inverse-variance weighted meta-analysis.** Each square represents the effect size for a particular cohort and error bars are the 95% CI. Diamond represents the estimate after meta-analysis. The index variant for the following loci is shown: A) *EBF1*, B) *WNT4*, C) *EEFSEC*, D) *ADCY5* and E) *AGTR2*. Sample size for each study cohort and after meta-analysis is detailed in parentheses after the cohort name.

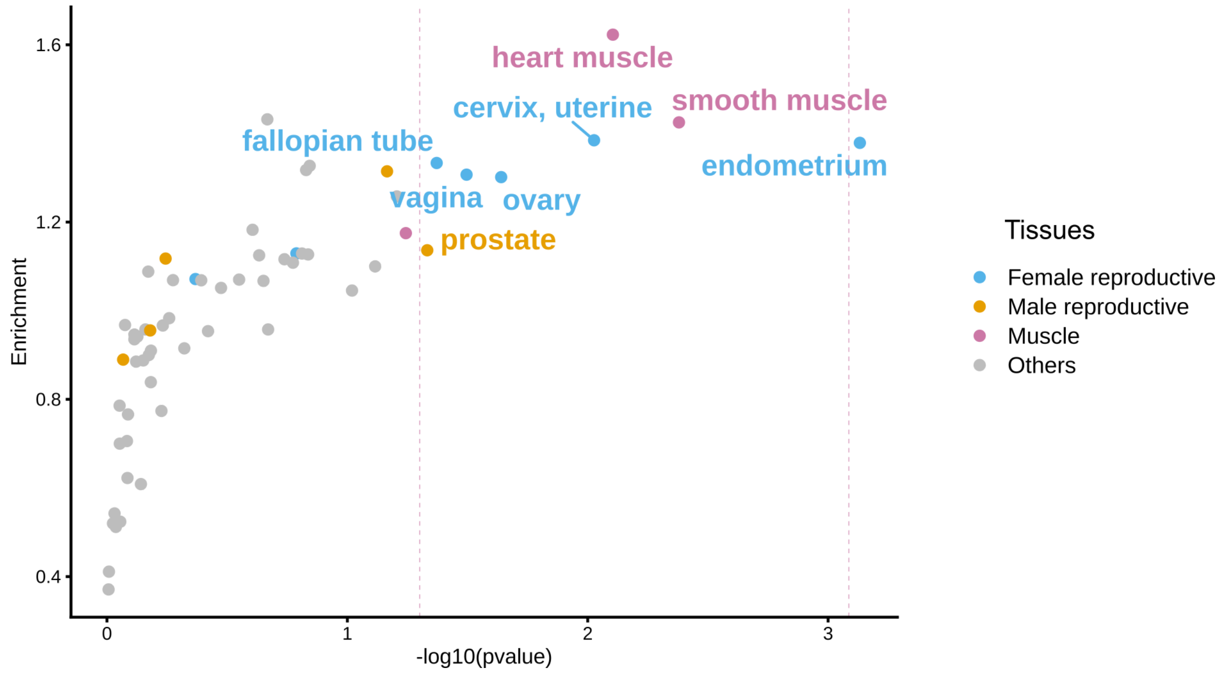


**Supplementary Fig. 3. RNA tissue-specific enrichment of nearest protein coding genes to gestational duration index SNPs.** Tissue-specific RNA was obtained from the Human Protein Atlas. The x-axis shows the -log10(p-value) for enrichment using a Wilcoxon test, and the y-axis the enrichment (median in nearest protein coding genes / median all other genes).

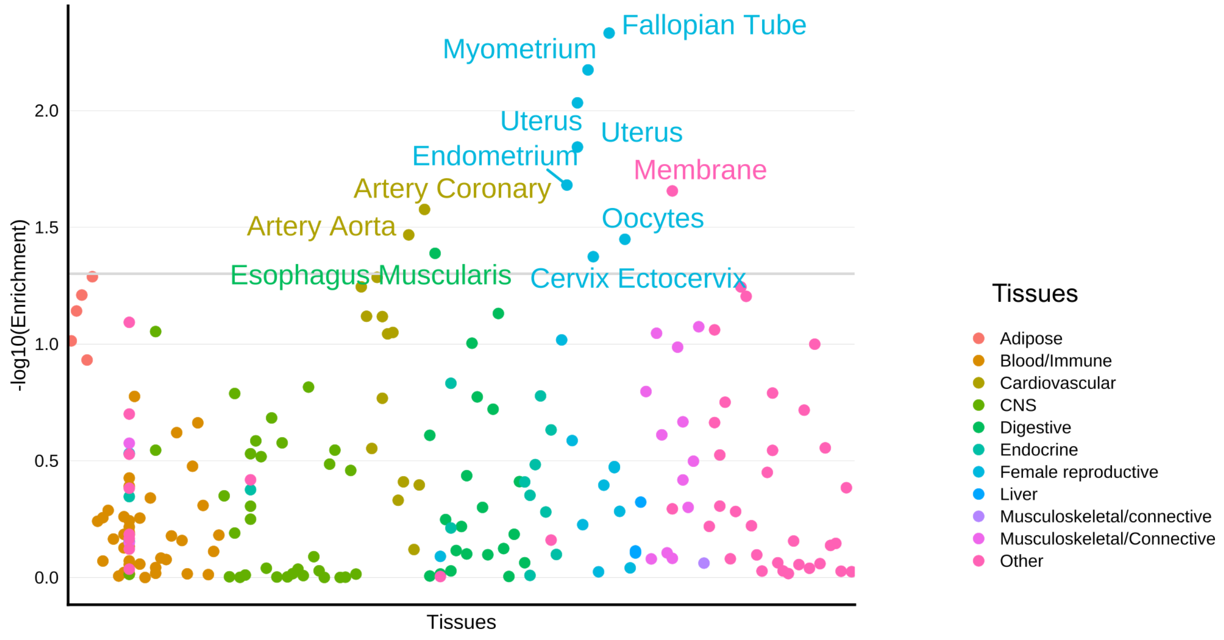


**Supplementary Fig. 4. Enrichment of SNP-heritability of gestational duration for tissue-specific gene expression.** Partitioned LD-score regression was used to estimate enrichment of SNP-heritability in 205 cell-types/ tissues, with pre-calculated LD-scores. Each dot represents a cell-type/ tissue; the ones that are labeled have a significant enrichment (p-value < 0.05, marked with a dark gray horizontal line).

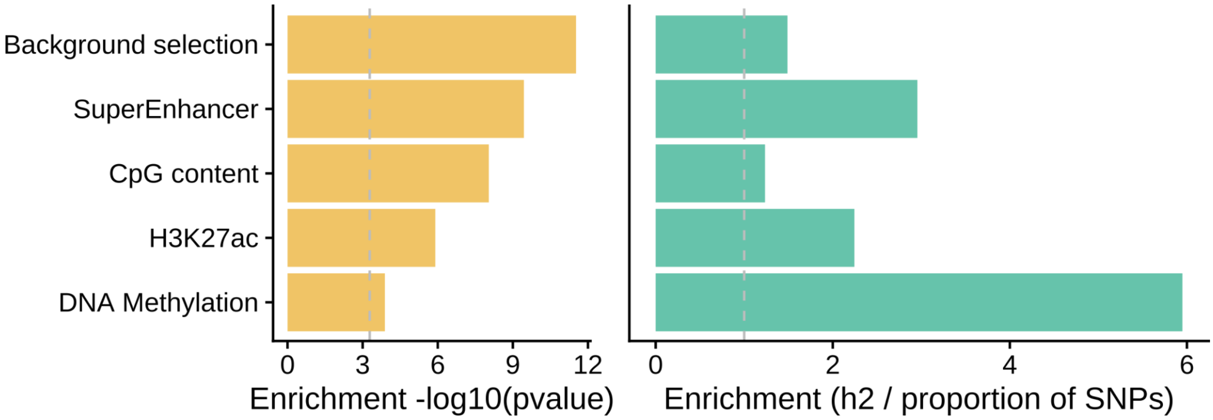


**Supplementary Fig. 5. Enrichment p-values for partitioned LD-score regression analysis of gestational duration SNP heritability.** The left plot shows the LD-score regression enrichment p-value for the five functional annotation categories that passed a Bonferroni correction (p-value< 0.05 / 97 categories).

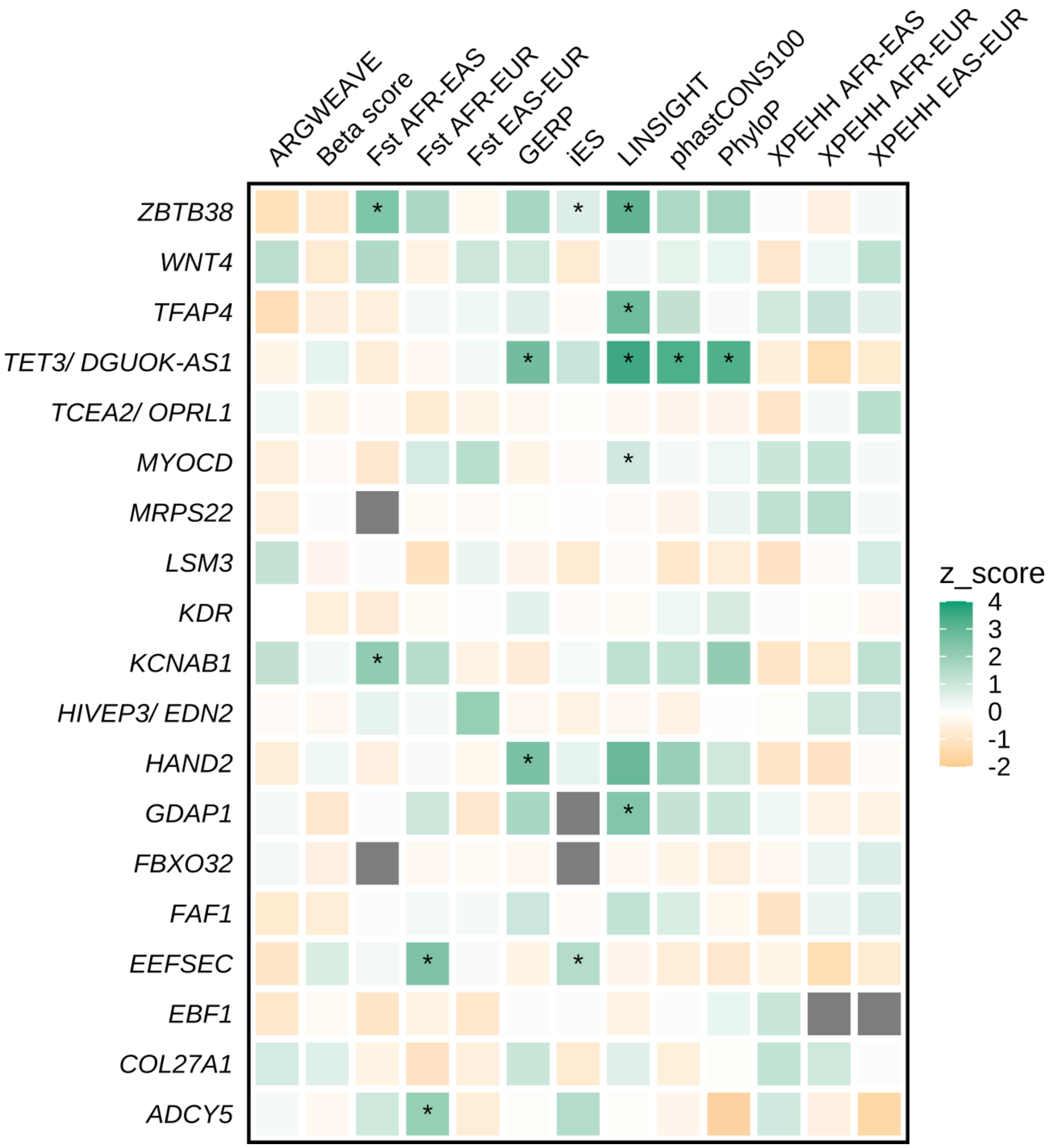


**Supplementary Fig. 6. Evolutionary metrics enrichment for gestational duration loci.** Enrichment analysis was performed using the MOSAIc pipeline for all regions with genome-wide significant associations with gestational duration, except the *HLA* gene region and the two in the X chromosome (*AGTR2* and *RAP2C*). The z-score for enrichment was obtained from a distribution that corresponds to the metric of the evolutionary force for 5,000 matched controls. The asterisk shows the regions significantly enriched for a specific evolutionary metric.

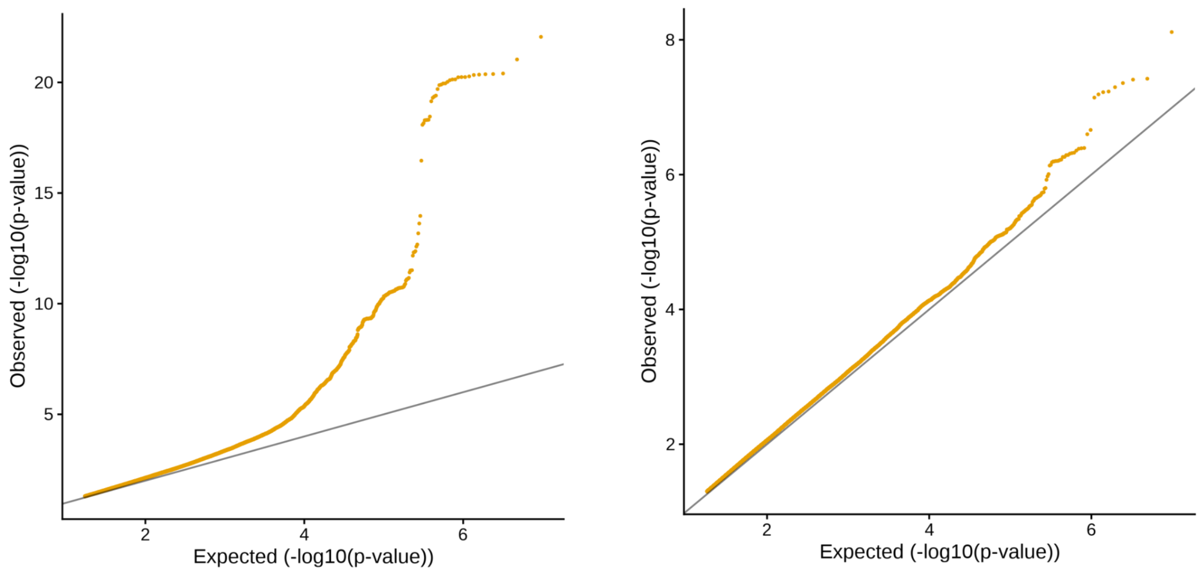


**Supplementary Fig. 7. Quantile-quantile plot of preterm and post-term delivery GWAS meta-analysis.**

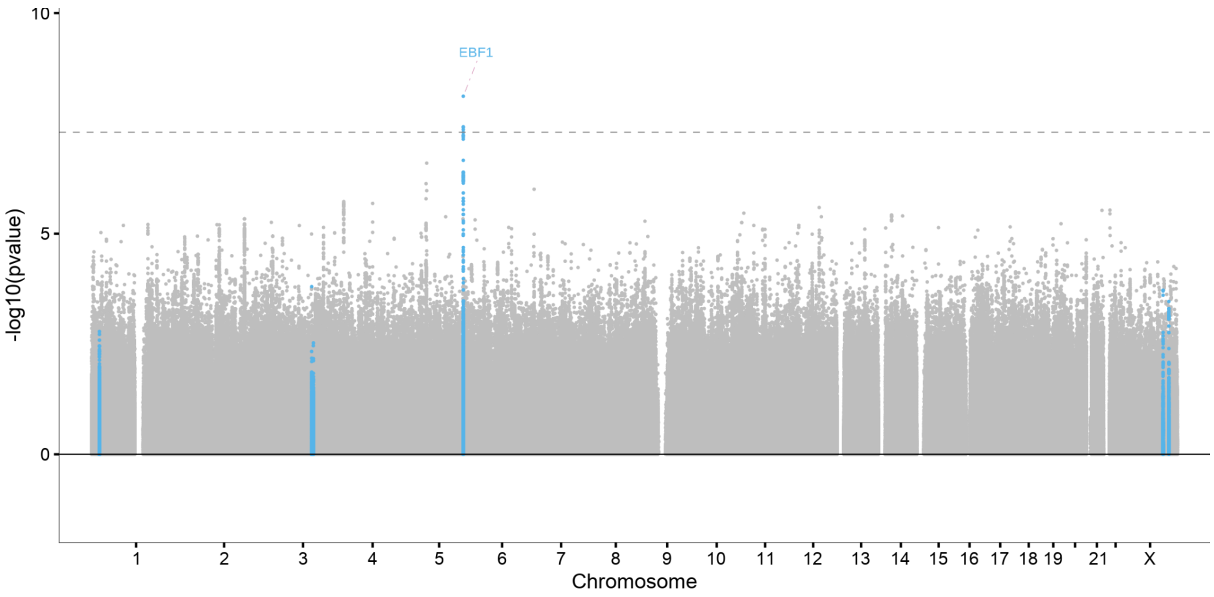


**Supplementary Fig. 8. Manhattan plot of the meta-analysis of GWAS of post-term delivery.** In blue, loci previously associated with gestational duration. n = 131,279, n cases = 15,972.

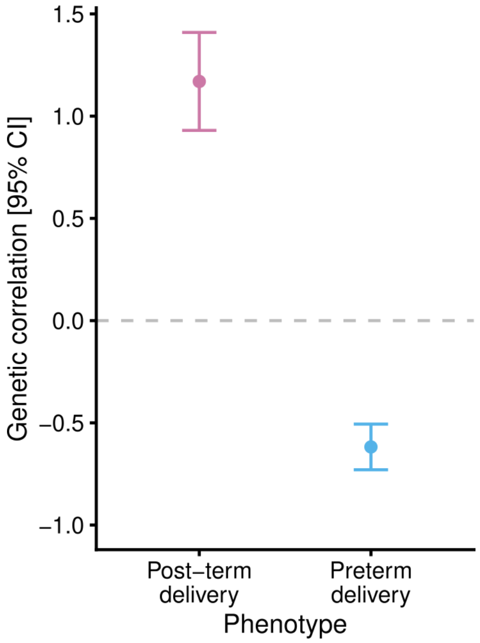


**Supplementary Fig. 9. Genetic correlations between gestational duration and preterm and post-term deliveries.** LD-score regression was used to estimate the genetic correlation with pre-computed LD-scores derived from samples with recent European ancestry from the 1000 Genomes Project.

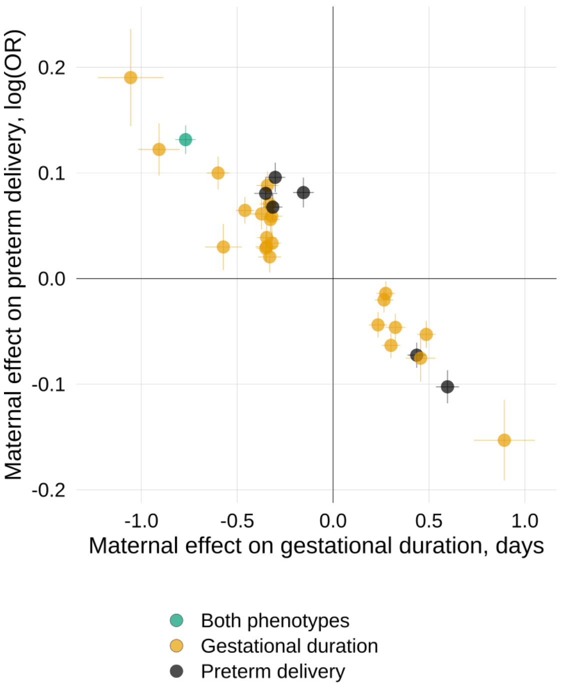


**Supplementary Fig. 10. Maternal effects on gestational duration and preterm delivery for the index SNPs on the two phenotypes.** Yellow, index SNPs of GWAS of gestational duration. Black, index SNPs of GWAS of preterm delivery. Green, index SNPs shared between gestational duration and preterm delivery at the *EBF1* locus. Horizontal and vertical bars represent the standard errors.

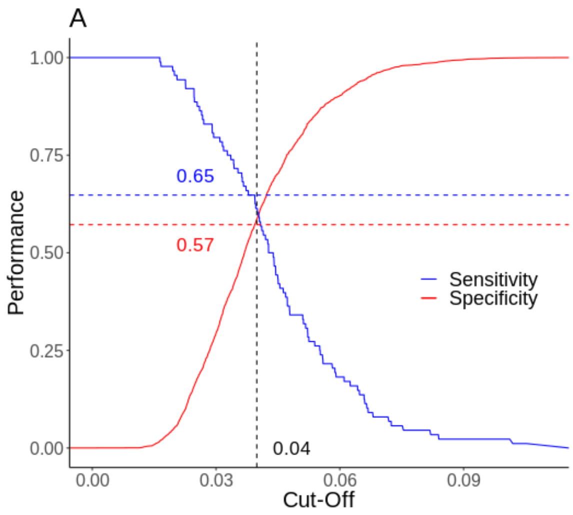


**Supplementary Fig. 11. Polygenic score optimal sensitivity and specificity for preterm delivery.** Optimal probability threshold cut-off that maximizes sensitivity and specificity of the gestational duration (A) and preterm delivery (B) polygenic scores predicting preterm delivery.

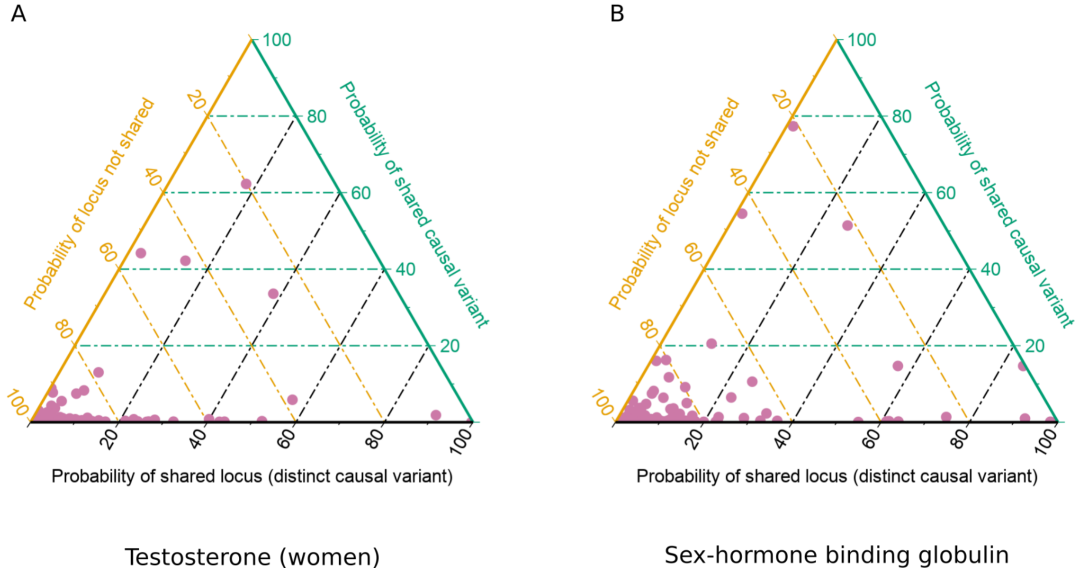


**Supplementary Fig. 12. Ternary plot for the genome-wide colocalization probabilities of gestational duration and testosterone and sex-hormone binding globulin in women.** The sum of all probabilities for each approximately independent region (n regions = 1,703) is 1, with each pink dot representing one of such regions. Lines are coloured according to the axis they belong to. All points in a horizontal green line have the same probability of “sharing the causal variant” between gestational duration and the hormone, points on a yellow line parallel to the right side of the triangle have the same probability of a “not sharing the locus” (i.e., locus has no association with either or both traits), and black lines parallel to the left side of the triangle have the same probability of a “sharing the locus, but with distinct causal variant”. Probabilities were obtained using coloc on previously published summary statistics for testosterone and sex-hormone binding globulin.

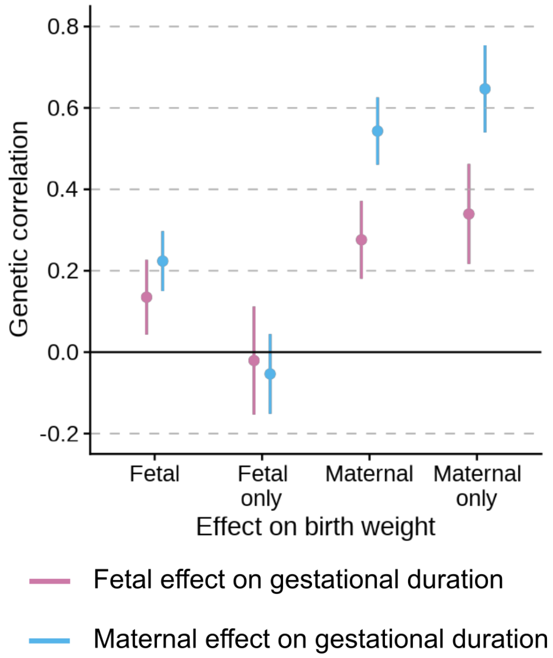


**Supplementary Fig. 13. Genetic correlations (95% CI) between the maternal (blue) and fetal (pink) effects on gestational duration and the maternal and fetal effects on birth weight.** The maternal only and fetal only effects on birth weight refer to the effects of the maternal genome after adjusting for the fetal effects and the effect of the fetal genome after adjusting for the maternal effects, respectively.

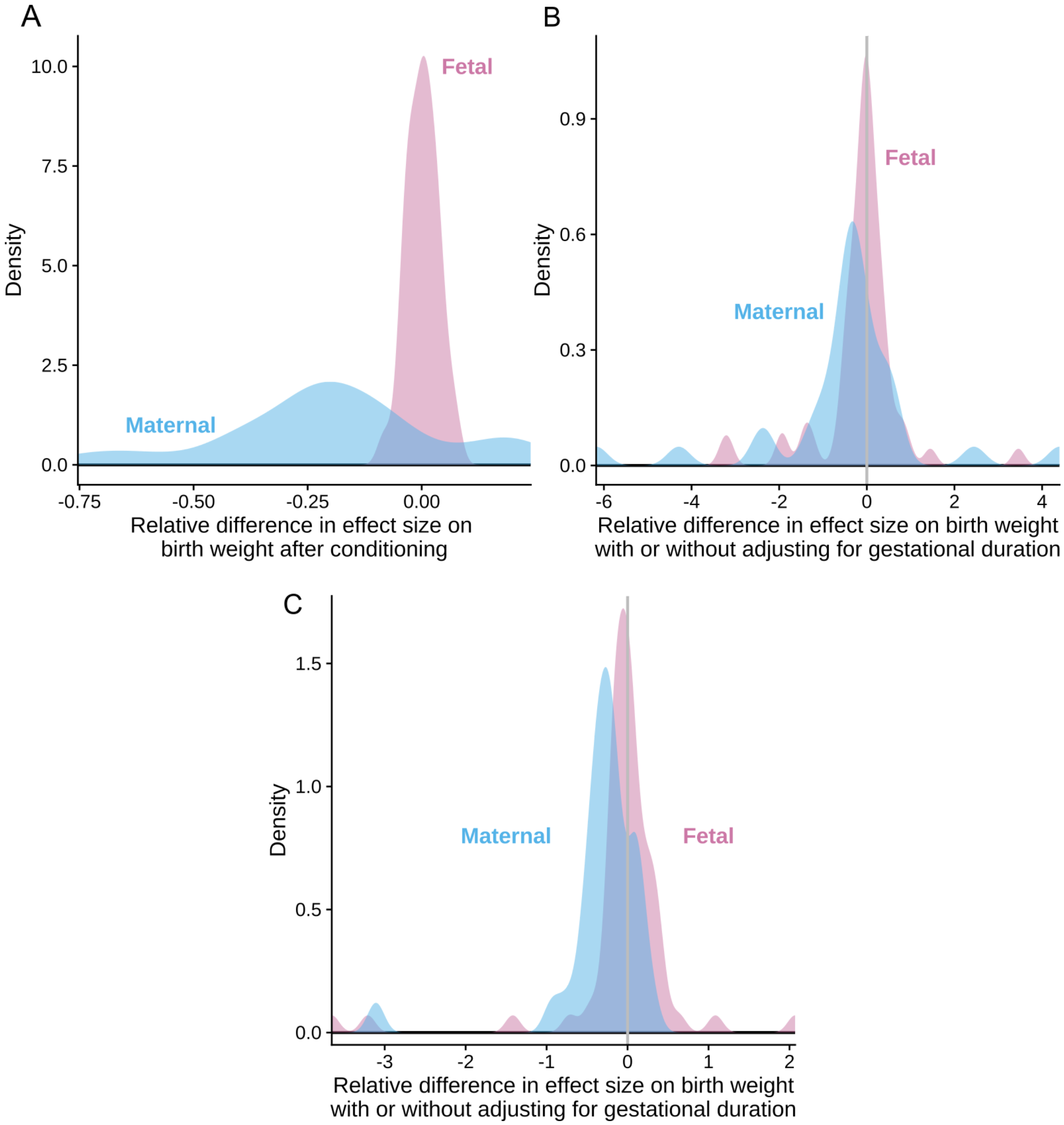


**Supplementary Fig. 14. Relative difference in effect size on birth weight before and after adjusting by gestational duration.** All analyses were performed on genetic variants classified as having a "Maternal Only" (n= 32) or a "Fetal Only" (n = 86) effect on birth weight in Warrington, et al, 2019, Nature Genetics [9]. In blue, relative difference in effect sizes for the maternal only effects on birth weight before and after conditioning; in pink, relative difference in effect sizes for the fetal only effects on birth weight after conditioning. A, Conditional analysis was performed using approximate multi-trait conditional and joint analysis in summary statistics. B, Relative difference in effect size on birth weight with and without adjusting for gestational duration using the maternal non-transmitted and paternal transmitted alleles (n = 21,060 parent-offsprings from MoBa) for the maternal and fetal effects on birth weight, respectively. C, Relative difference in effect size on birth weight with and without adjusting for gestational duration using genotype dosage in 32,511 mothers and 16,387 fetuses from Iceland.

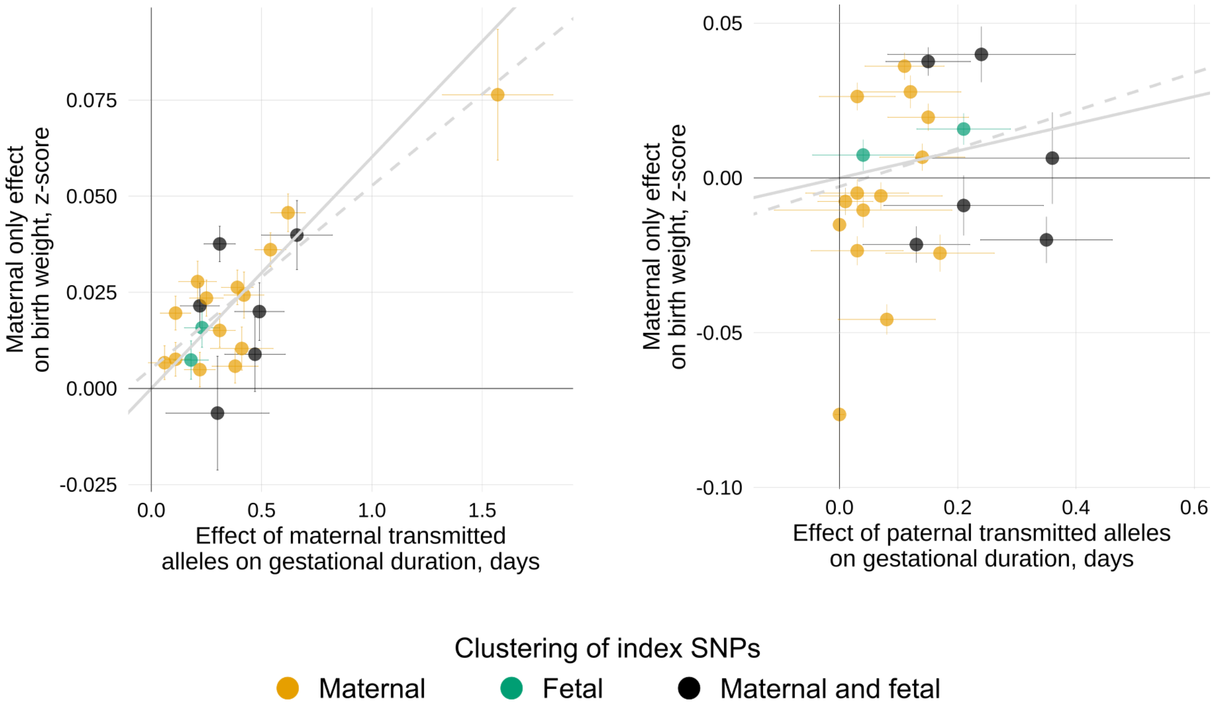


**Supplementary Fig. 15. Association between maternal and paternal transmitted gestational duration-increasing alleles and maternal effect on birth weight.** Scatterplot for two-sample Mendelian randomization analysis for the effect of gestational duration on birth weight (maternal effect). Each dot represents one of the gestational duration index SNPs. The x-axis shows the SNP effect of the maternal or paternal transmitted alleles on gestational duration (meta-analysis of multiple data sets, including data from Iceland, MoBa, HUNT, ALSPAC, DNBC, FIN, GPN, and HAPO; n = 136,833), and the y-axis the maternal only effect on birth weight (weights were obtained from a previously published GWAS; n = 210,248). Horizontal and vertical error bars represent the standard error. The gray line depicts the inverse-variance weighted method estimate, and the gray-dashed line the MR-Egger estimate. Colors represent the clustering of the index SNP.

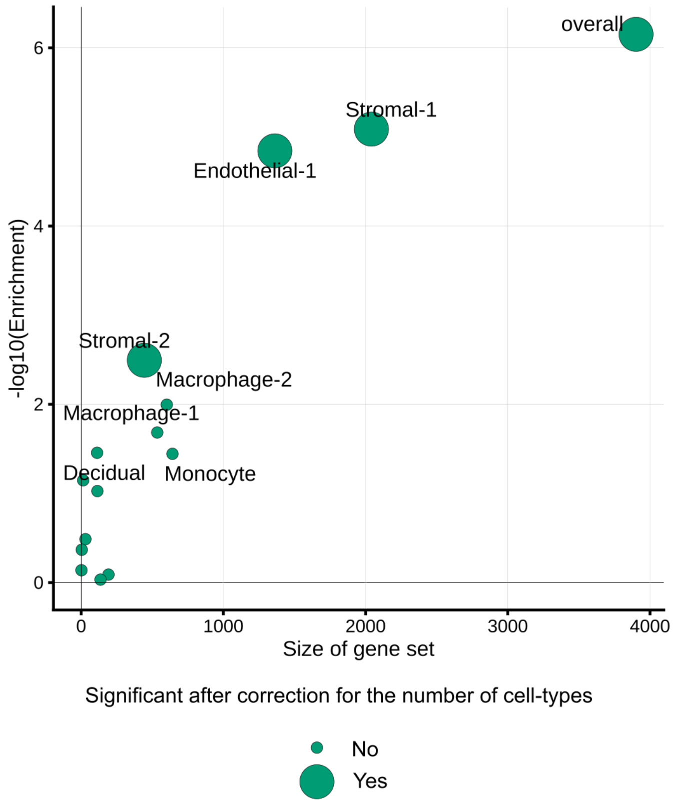


**Supplementary Fig. 16. Gene set size and SNP-heritability enrichment of gestational duration for genes differentially expressed during labor in different cell types of the myometrium and overall.** LD-score regression was used to partition heritability, and estimate the heritability enrichment p-value for each cell type and overall. The x-axis shows the number of genes included in each cell-type and overall; the y-axis shows the -log10(p-value) for enrichment. We calculated LD scores (European individuals from phase 3 of the 1000 Genomes project) for each set of genes differentially expressed at labor (± 100 kb), separately for each cell type and for the overall set of genes differentially expressed in the myometrium. Each dot represents a cell type, with larger showing significant heritability enrichment after correcting for the number of cell types.
